# Investigating the potential causal relationship between parity and long-term maternal cardiometabolic health outcomes using Mendelian randomization

**DOI:** 10.64898/2026.08.09.26360053

**Authors:** Caroline Brito Nunes, Abigail Fraser, Gunn-Helen Moen, Alesha A Hatton, David M. Evans

## Abstract

**Background:** Multiple observational studies have reported associations between greater parity and increased CVD risk. Whether these associations reflect causal effects or are confounded by socioeconomic factors remains unclear.

**Methods:** We investigated associations between number of children ever born (NEB) and 16 cardiometabolic traits in up to 172,122 females and 138,390 males in the UK Biobank, and an independent sample of 53,237 UK Biobank spousal pairs. We additionally conducted sex-stratified two-sample Mendelian randomization (MR) and applied a novel spousal MR framework, in which an individual’s spouse’s genotype was used as the instrumental variable to estimate the causal effect of NEB on cardiometabolic health outcomes, as an approach to minimize bias from horizontal pleiotropy.

**Results:** NEB was associated with multiple cardiometabolic traits in the multivariable regression, even after adjustment for socioeconomic status, with differences in the strength of association observed between males and females. Traditional MR provided evidence that higher NEB causally increases type 2 diabetes risk in females, body mass index (BMI) in both sexes, female basal metabolic rate (BMR) and male body fat percentage but decreases female blood pressure. Spousal MR corroborated positive effects on female BMI and BMR and additionally suggested inverse causal effects on female HDL cholesterol and ApoA1 and male blood glucose.

**Conclusion:** These findings indicate a possible causal relationship between NEB and long term cardiometabolic health, although causal effects are likely to be small.

## Background

Pregnancy triggers a wide range of maternal physiological adaptations - including changes in glucose and lipid metabolism, blood pressure (BP) regulation, and cardiovascular function - to meet the needs of both the mother and the fetus[1,2]. While many of these changes return to a pre-pregnancy baseline postpartum[3,4], higher parity has been linked to long-term adverse maternal cardiovascular health.

Observational studies have suggested that the physiological stress of multiple pregnancies may increase the risk of diabetes and cardiovascular disease (CVD), including heart disease, stroke, and hypertensive disorders. However, findings have been inconsistent, with some studies reporting the inverse[5] or no relationship[6–10], while others describe positive[11–14] or J-shaped non-linear associations[15–22]. In comparison, evidence linking parity with body mass index (BMI) and BP appears more consistent, with higher parity generally associated with a higher maternal BMI[23–34], but lower maternal BP[35,36].

These associations likely reflect a complex interplay of physiological, socioeconomic, and lifestyle factors that are difficult to disentangle. However, comparing associations by sex can provide additional insight as males serve as negative controls - any association observed in males cannot be driven by pregnancy, and likely reflects shared behavioural and lifestyle changes associated with childrearing. Supporting this, one large-scale study reported similar parity-CVD associations in both sexes, suggesting that effects in females may be largely driven by these shared factors rather than pregnancy itself[37].

Observational studies are also often limited in their ability to control for confounding, and Mendelian Randomization (MR) provides a complementary framework for estimating causal effects. By leveraging genetic variants as instrumental variables (IVs), MR is less susceptible to confounding and reverse causation than conventional observational epidemiological approaches[38]. To date, only one MR study has investigated parity and maternal CVD risk, reporting positive causal effects on atrial fibrillation, heart failure, and stroke[39]. A large study (N = 444,611) conducted in India further used firstborn sex as an IV - exploiting male offspring preference - and reported modest but sustained reductions in systolic and diastolic BP with each additional child, persisting for over a decade postpartum[40].

In this study, we triangulated evidence from multivariable regression and two complementary MR approaches to investigate the effects of number of children ever born (NEB) on adult cardiometabolic traits in both sexes (**Figure 1**). First, we performed regression analyses of cardiometabolic traits on NEB in the UK Biobank (UKBB) adjusting for key confounders. We tested for sex heterogeneity; stronger effects in females would support pregnancy-specific mechanisms, whereas similar effects across sexes would suggest shared behavioural and lifestyle factors associated with childrearing. We then performed MR using two approaches: i) sex-stratified two-sample MR, comparing heterogeneity in causal effect estimates between sexes and ii) a novel spousal MR method. Unlike traditional MR, which uses an individual’s own genetic variants and phenotype, spousal MR estimates the causal effect of NEB on an individual’s outcome (e.g. female cardiometabolic traits), using their partner’s genotype as an IV (e.g. male genotype for female outcomes). Because the IV is derived from the partner rather than the individual, it is theoretically less susceptible to bias arising from the individual’s own genotype affecting the outcome through pathways independent of NEB (see **Figure 2** for a more nuanced discussion).

**Figure 1.**
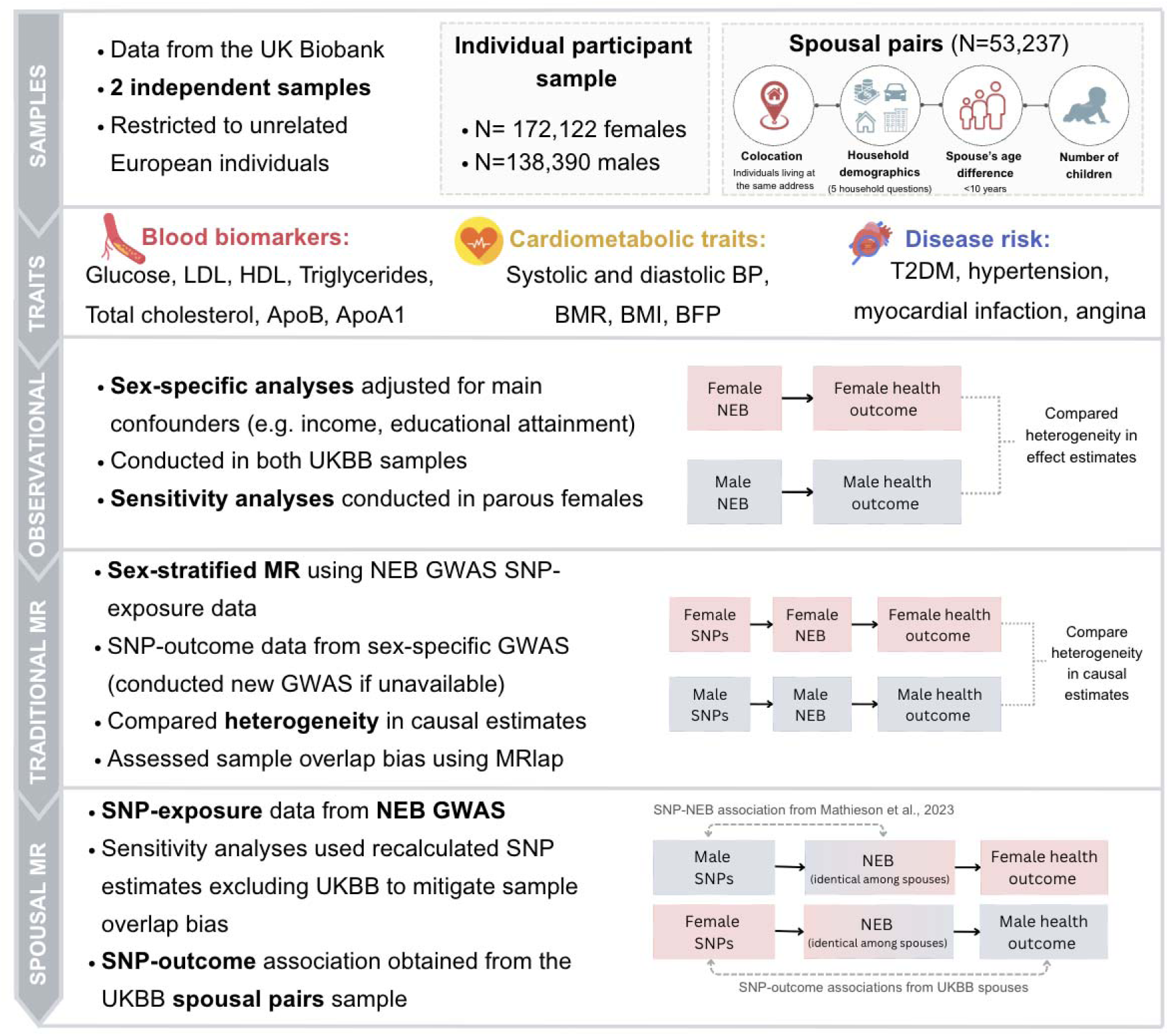
Summary of the data resources, study sample and analyses conducted in this study. MR; Mendelian randomization. MR; Mendelian randomization. GWAS; genome-wide association study. LDL; low-density lipoprotein cholesterol. HDL; high-density lipoprotein cholesterol. ApoB; apolipoprotein B. ApoA1; apolipoprotein A1. BP; blood pressure. BMR; basal metabolic rate. BMI; body mass index. BFP; body fat percentage. NEB; number of children ever born. SNP; single-nucleotide polymorphism. EA; Educational attainment.

**Figure 2.**
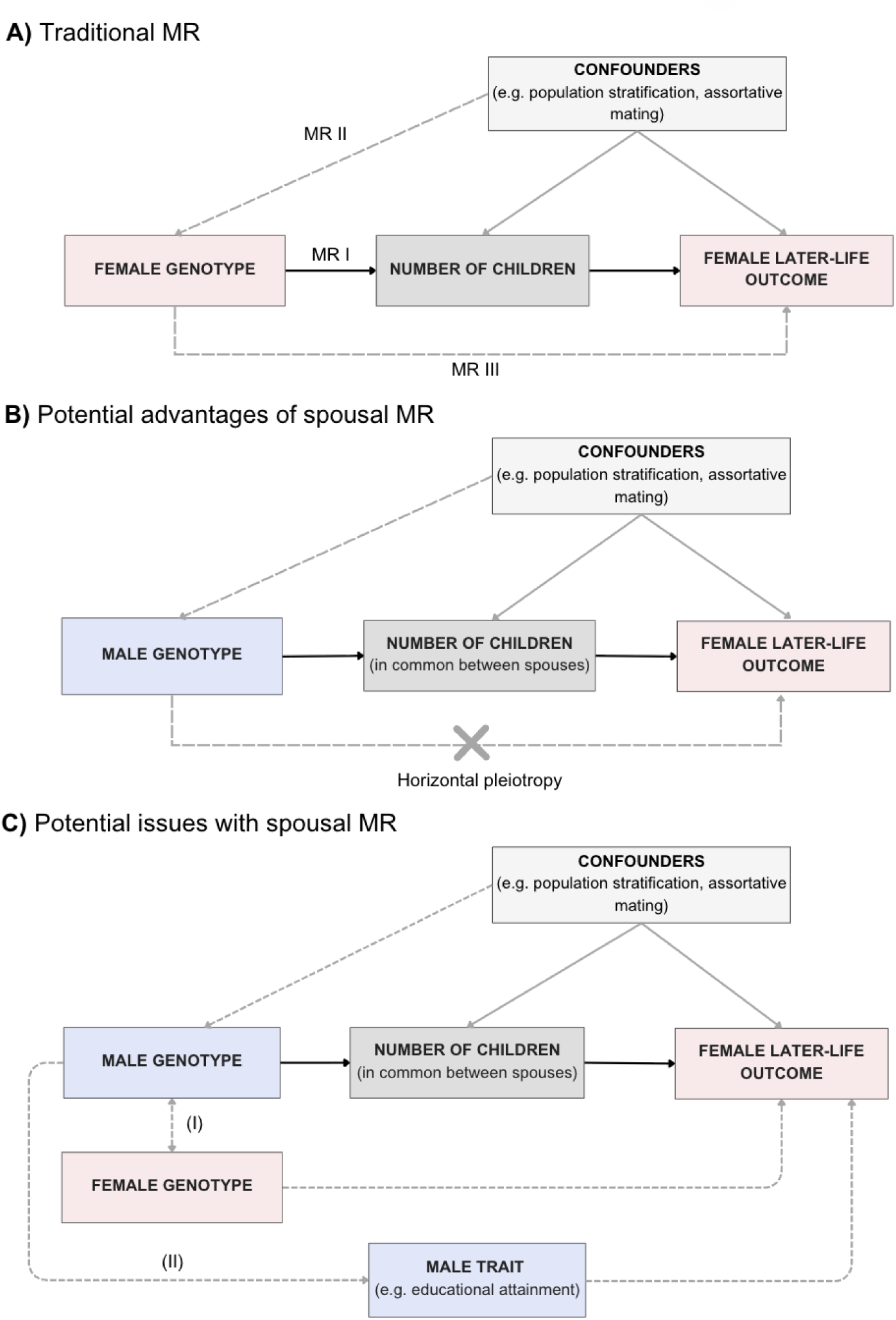
Illustration of the spousal MR study design to investigate the relationship between number of children ever born (NEB) and female later-life outcomes. Diagram **A** shows the traditional design for MR, where female genotype is assumed not to pleiotropically influence later life outcomes via pathways that do not go through the exposure variable. Diagram illustrates the three core instrumental variable assumptions: relevance (MR I), independence (MR II), and the exclusion restriction (MR III). Diagram **B** shows that male genotype can be used instead of female genotype to proxy NEB if spouses are matched on the number of children, potentially reducing the likelihood of violations of the exclusion restriction assumption (i.e. no horizontal pleiotropy). This same framework can be used to assess whether NEB is causally associated with male later-life outcomes but with female variants proxying the exposure. Diagram **C** shows potential violations of the exclusion restriction assumption using spousal MR. Path (I) shows that if male and female spousal genotypes are correlated (e.g. as under assortative mating), this may create a path from the genetic instruments to the outcome if those variants pleiotropically affect the outcome. However, under random mating, maternal and paternal genotypes should be uncorrelated (i.e. no assortative mating on exposure or traits related to the exposure). Path (II) shows that if the male genotype influences a male phenotype other than number of ever births (e.g. educational attainment or socioeconomic status), it may have subsequent effects on the partner’s cardiometabolic trait. MR; Mendelian Randomization. NEB; Number of children ever born.

## Methods

### The UK Biobank cohort

The UKBB is a large, prospective population-based cohort of over 500,000 participants aged 37-73 years at baseline, with extensive health-related and genome-wide genetic data. Participants provided informed consent at recruitment. Genotype pre-imputation quality control, phasing, and imputation are described in Bycroft et al., 2018[41]. In addition to UKBB’s central quality control, we excluded individuals who withdrew consent or failed standard quality checks (e.g., due to sex mismatch, high heterozygosity, missingness, sex chromosome aneuploidy; N=1,932).

### UKBB spousal pair sample

We identified opposite-sex spousal pairs using a returned dataset from Tenesa et al. (2016)[42], which reports spouses identified primarily through colocation information along with other matching variables. To further reduce misclassification of non-spouses residing at the same address, we applied additional household-based matching criteria. Analyses were restricted to unrelated spouses of European ancestry, showing concordant NEB and with available phenotype data (N = 53,325). Full details of the spousal identification, validation, and filtering procedures are provided in **Supplementary Note 1.0, Supplementary Tables 1-2,** and **Supplementary Figures 1-3.**

### UKBB individual participant sample

The initial UKBB sample included 217,772 females and 184,424 males reporting relevant phenotypes. After excluding those in the spousal pair sample and restricting to unrelated individuals of European ancestry (IBD<0.08), the final sample included 172,122 females and 138,390 males.

### Phenotype selection in the UKBB

In the multivariable analyses, NEB was defined as number of live births (females) or children fathered (males). We focused on live-born offspring to capture completed pregnancies encompassing gestation, postpartum adaptation, and lifestyle and behavioural changes associated with childrearing.

Outcomes of interest included:

*i)* Disease outcomes: angina, myocardial infarction, hypertension and type-2 diabetes (T2DM), with sufficient sample sizes for analyses within spousal pair. T2DM was ascertained via self-reported doctor diagnosis ("Has a doctor ever told you that you have diabetes?") and likely captures predominantly type 2 diabetes, though some type 1 cases may also be included.
*ii)* Cardiometabolic traits: systolic and diastolic BP, body fat percentage (BFP), BMI, and basal metabolic rate (BMR). BMI was included as a positive control, given its consistent association with higher parity in observational studies.
*iii)* Lipids and blood biomarkers: apolipoprotein A-1 (ApoA1), Apolipoprotein B (ApoB), HDL, low-density lipoprotein (LDL), triglycerides, total cholesterol and blood glucose levels.

Covariates included birth year and sociodemographic characteristics (household income and educational attainment (EA)), though we also tested associations between NEB and both income and EA.

Quantitative traits with values >4 standard deviations from the mean were excluded. Phenotypes were standardized (mean 0, variance 1) to facilitate comparability across traits, with untransformed results presented in the main text for interpretability in original units; inverse normal-transformed analyses are reported in Supplementary Tables. **Supplementary Tables 3-4** provide phenotype definitions, measurement details, frequency tables, and sample sizes.

### Multivariable regression analyses in the UKBB

We regressed outcomes on NEB in both the spousal pair and individual participant samples. The first regression model adjusted for the individual’s year of birth and the first ten genome-wide genetic principal components (PCs), and a second model additionally adjusted for income and EA. Logistic, ordinal logistic, or linear regression were used for binary, ordered categorical, and continuous outcomes, respectively. Analyses were sex-stratified, with men conceptualised as a negative control, and sex differences in the magnitude of regression coefficients assessed using heterogeneity Z-tests.

Sensitivity analyses excluding nulliparous women were performed to account for unmeasured confounding (e.g. health conditions affecting both fertility and cardiometabolic outcomes), and socioeconomic differences between nulliparous and parous females that may disproportionately drive observed associations (up to 135,620 females in the individual sample and 47,791 in the spousal subset).

### MR analyses

To investigate whether NEB causally influences cardiometabolic outcomes and disease risk, we conducted two-sample MR analyses using two complementary approaches: sex-stratified traditional MR and spousal MR.

#### Exposure GWAS

NEB-associated variants were obtained from a previous GWAS meta-analysis of up to 785,604 European-ancestry individuals (including the UKBB)[43]. The sex-combined GWAS identified 28 independent genome-wide significant SNPs, with an additional four and two SNPs identified in female- and male-specific analyses, respectively (**Supplementary Table 5** and **7)**. We define female IVs as the set of sex-combined plus female-specific SNPs, and male IVs as the sex-combined plus male-specific SNPs; these definitions are used consistently throughout all MR analyses. Instrument strength was assessed using F-statistics.

#### SNP-outcome associations in sex-stratified traditional MR

Sex-specific European GWAS summary statistics were used for the traditional MR analyses for the same outcomes of interest listed above (**Supplementary Table 6**)[44–50]. For traits lacking publicly available sex-specific data, we performed de novo GWAS in the UKBB stratified by sex (**Supplementary Material 2.0; Supplementary Figures 4-8**). Proxies (r² > 0.7) were identified using LDmatrix[51] and used when lead NEB variants were unavailable (**Supplementary Table 5**).

#### SNP-outcome associations in spousal MR

SNP dosages for all 34 NEB GWAS SNPs[43] were extracted from the UKBB imputed data using Plink (v2.0). SNP-outcome associations were derived by regressing spouses’ outcomes on their partner’s IVs (e.g. female outcomes on male IVs and vice versa), using the same phenotypes and trait definitions as the multivariable analyses, and adjusting for ten male and ten female genome-wide PCs. Sample sizes for each outcome are provided in **Supplementary Table 4**.

#### Sample overlap bias in MR

Because both exposure and outcome GWASs included UKBB participants, we applied MRlap to complement traditional MR analyses and correct for sample overlap, winner’s curse, and weak instrument bias[52]. As MRlap allows only a single exposure GWAS, we used sex-combined NEB summary statistics, in which most instruments were identified. Outcome associations were obtained from sex-specific GWASs (**Supplementary Table 6**). Instruments included all lead SNPs from the sex-combined GWAS, and additional loci identified in sex-specific analyses; however, effect estimates for all instruments were derived from the sex-combined NEB GWAS, as required by MRlap. Proxy SNPs were used when exposure variants were unavailable in the outcome GWAS (**Supplementary Table 5**).

For spousal MR analyses, NEB-associated SNP effect estimates were recalculated excluding UKBB participants to address sample overlap bias (**Supplementary Table 7**; methods in **Supplementary Materials 3.0**). Sensitivity analyses were performed using these recalculated SNP-exposure associations.

#### Validating assumptions in traditional MR

NEB SNPs were cross-referenced against the GWAS Atlas[53] and GWAS Catalog[54] to identify potential pleiotropic variants that would violate the MR exclusion restriction assumption (**Supplementary Table 5**). We additionally examined associations between NEB SNPs and EA in the entire UK Biobank sample (**Supplementary Table 8**). Heterogeneity was assessed using Cochran’s Q statistic, and directional pleiotropy with the MR-Egger intercept.

#### Validating assumptions in spousal MR

To assess potential violations of MR assumptions due to correlation between spouses’ genotypes (**Figure 2c, Path I**), we constructed sex-specific NEB genetic risk scores (GRS) using effect sizes from the NEB GWAS, with dosages reflecting alleles associated with increased NEB[43]. Within spousal pairs, each GRS was regressed on ten male and ten female genome-wide PCs, and Pearson correlations between spouses’ GRS were calculated using the residuals.

To assess potential violations due to horizontal pleiotropy via dynastic effects (**Figure 2c, path II**), we tested NEB SNPs for pleiotropic associations with socioeconomic and lifestyle variables (smoking, and EA) both within individuals and across spouses (using an individual’s own genotype and their partner’s phenotype) (**Supplementary Table 8**).

#### Statistical analyses

The inverse-variance weighted (IVW) method was used as the primary estimator, complemented by MR-Egger[55], Weighted Median[56], Simple Mode and Weighted Mode[57] methods. Analyses were conducted using the TwoSampleMR package in R[58]. To explore potential sex differences in IVW causal effect estimates in traditional MR, we performed Z-tests.

Given the strong association between EA and NEB, we performed additional analyses excluding SNPs associated with EA in the UKBB sample (P<0.05) in the corresponding sex; most of which were also genome-wide significant for EA in look-ups in GWAS Atlas[53] and GWAS Catalog[54]. Seven SNPs were excluded from both sexes, with additional sex-specific exclusions in females and males (**Supplementary Table 8**).

### Multiple Testing Correction

We performed principal component analyses, using the prcomp function in R, across 95,197 females and 78,711 males with complete outcome data to estimate the number of independent principal components that explain the majority of variation in our outcomes. In both sexes, eight PCs accounted for 80% of the shared variance across outcomes. We therefore evaluated our statistical tests against a modified significance threshold of P<0.05/8 = 6.25 10^-3^ (**Supplementary Figures 9-10**).

### Power Calculations

We used the MR power calculator (https://shiny.cnsgenomics.com/mRnd/) to estimate the causal effect size detectable with 80% power in both traditional and spousal MR analyses, assuming a (two-sided) type 1 error rate of α = 0.05. The variance of NEB (i.e. the exposure variable) was assumed to be 2.4 (similar to NEB in the UKBB individual participant sample, as the variance/standard error of NEB was not reported in the NEB GWAS) whereas the variance for continuous outcomes was assumed to be one (outcomes were standardised). We used the approximate sample size for BMI (400,000 in traditional MR and 53,000 in the spousal MR), an R^2^_xz_ value of 0.19% (see **Supplementary Table 9** for R^2^ calculations) and an observational effect estimate of 0.0195 (from our female BMI multivariable analysis correcting for birth year, PCs, income and EA). Power was also calculated for a binary outcome with a prevalence (K) of 6%, using N = 200,000 (traditional MR) and 53,000 (spousal MR), and the same parameters.

## Results

### Multivariable regression analyses in the UKBB

#### Overview of findings and adjustment for socioeconomic status

In the UKBB individual participant sample (N up to 172,122 females and 138,390 males), NEB was associated with all cardiometabolic outcomes before adjustment for income and EA (**Supplementary Table 10**). After adjustment, most associations attenuated and sex heterogeneity was reduced, though all traits remained robustly associated with NEB as discussed below (P<6.25×10^-3^, **Figures 3-5**; **Supplementary Table 10**). Associations in the spouse subset (N up to 53,237) were generally directionally consistent but attenuated with wider confidence intervals (CIs), likely reflecting reduced power (**Supplementary Table 10; Supplementary Figure 11**). Sociodemographic comparisons between the two samples are shown in **Supplementary Table 11** and **Supplementary Figures 12-17**.

**Figure 3.**
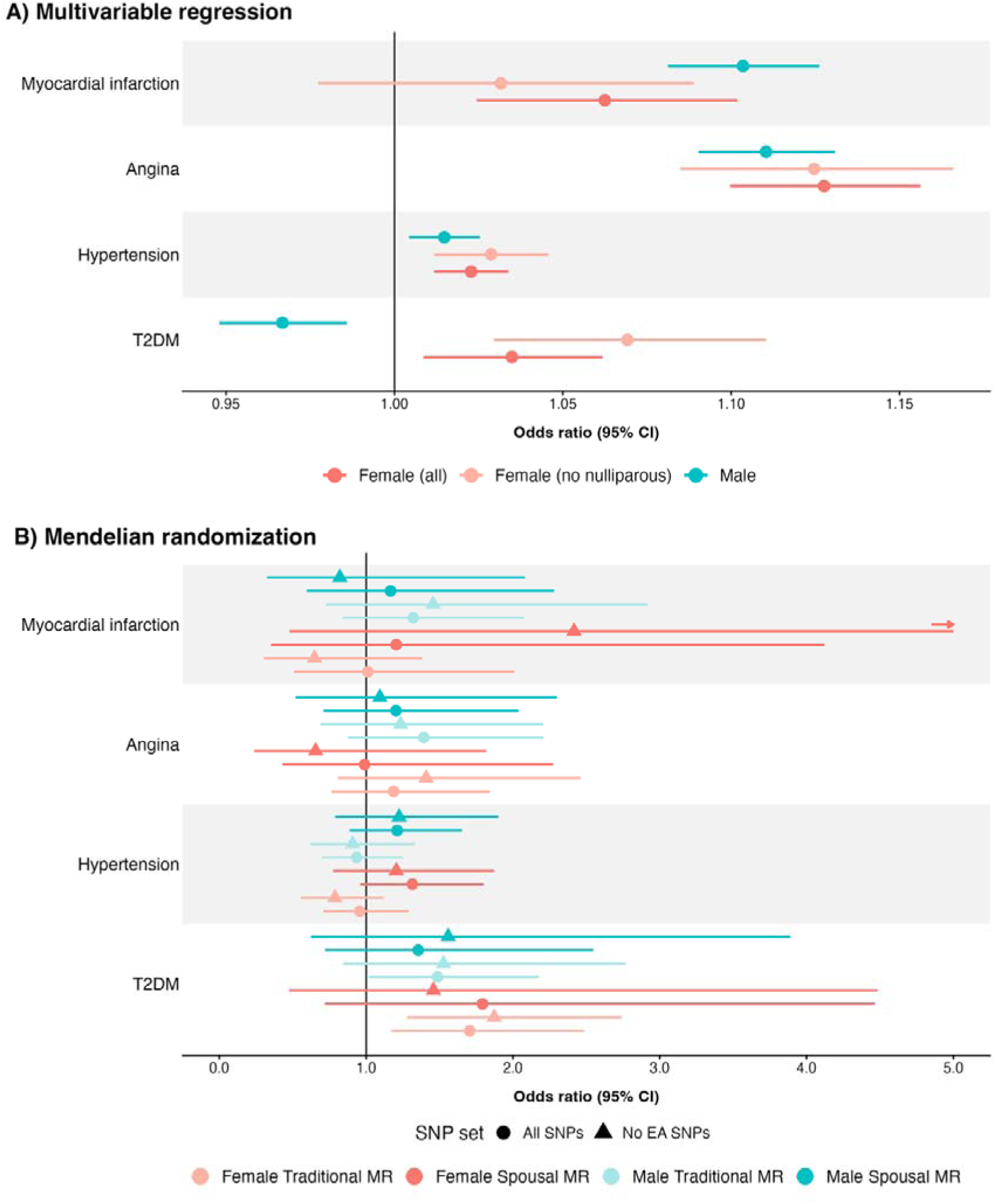
Associations between number of children ever born (NEB) and disease outcomes. Panel A shows the results from the multivariable regression analyses including all females (N=172,122), all males (N=138,390) and the sensitivity analysis including only parous females (N=135,620). Effect estimates are expressed as odds ratios (OR) representing the change in disease odds per additional child. Analyses were corrected for year of birth, ten genome-wide genetic principal components (PCs), income and educational attainment (EA). Estimates are presented for males (blue), all females (red) and only parous females (orange). Panel B shows the Mendelian randomization (MR) estimates using the inverse variance weighted (IVW) method for both the traditional and spousal MR frameworks. Analyses used genetic instruments and effect estimates for NEB from Mathieson et al. (2023). Round markers indicate estimates using all NEB SNPs; triangular markers show results after excluding SNPs associated with EA. SNP-outcome associations were derived from UK Biobank spouse pairs, testing (i) male SNPs against female outcomes and (ii) female SNPs against male outcomes, both adjusted for ten male and ten female genetic PCs. Points indicate effect estimates; horizontal bars represent 95% confidence intervals (CI). CI; Confidence intervals. EA; Educational attainment. IVW; inverse variance weighted. MR; Mendelian Randomization. NEB; number of children ever born. PC; Principal component. SD; Standard deviation. T2DM; Type-2 diabetes mellitus.

#### Associations with diabetes and cardiovascular outcomes

Higher NEB was associated with increased odds of angina, myocardial infarction, and hypertension in both sexes, with no evidence of sex heterogeneity (**Supplementary Table 10**; **Figure 3**). NEB was inversely associated with T2DM in males but positively associated in females, though the latter did not meet the multiple-test corrected threshold (P<6.25×10^-3^) (**Supplementary Table 10**; **Figure 3**).

#### Associations with cardiometabolic traits and blood biomarkers

In females, NEB was inversely associated with systolic BP, diastolic BP, and total cholesterol, whereas in males it was positively associated with ApoB and LDL cholesterol and inversely with blood glucose (**Figures 4** and **5; Supplementary Table 10**). These associations were sex-specific, and sex heterogeneity was evident for all traits except blood glucose (P<0.05; **Supplementary Table 10**). For the remaining traits, associations were present in both sexes: NEB was positively associated with triglycerides, BMI, BFP, and BMR, and inversely associated with HDL and ApoA1, with sex heterogeneity reflecting stronger associations for HDL and ApoA1 in females and for BMI, BFP, and BMR in males (**Figures 4** and **5; Supplementary Table 10**).

**Figure 4.**
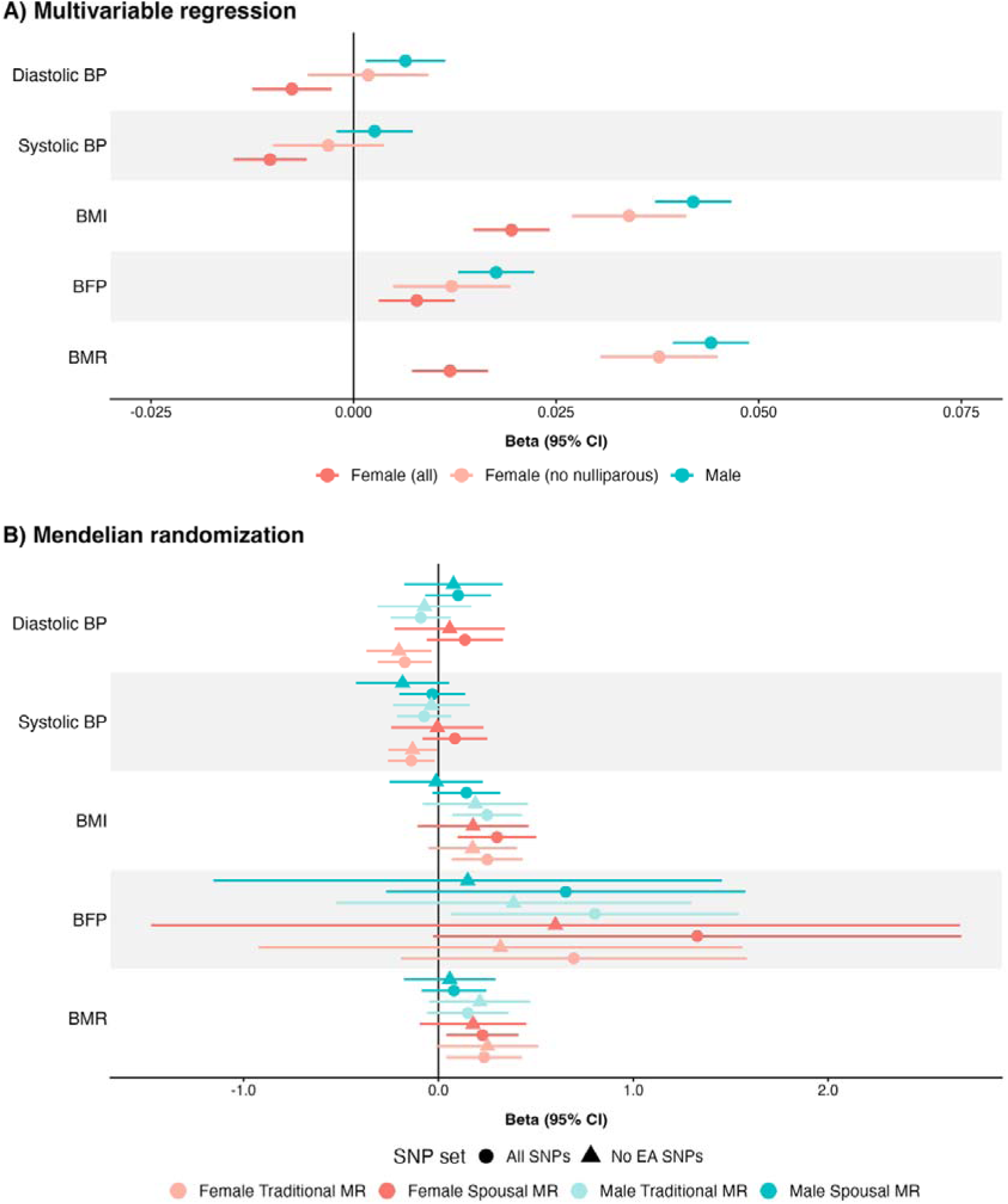
Associations between number of children ever born (NEB) and cardiometabolic traits. Panel A shows the results from the multivariable regression analyses including all females (N=172,122), all males (N=138,390) and sensitivity analysis including only parous females (N=135,620). Estimates represent linear regression coefficients corresponding to the change in the outcome per additional child, expressed in standard deviation (SD) units. Analyses were corrected for year of birth, ten genome-wide genetic principal components (PCs), income and educational attainment (EA). Associations plotted are for the untransformed but normalized continuous outcomes. Estimates are presented for males (blue), all females (red) and only parous females (orange). Panel B shows the Mendelian randomization (MR) estimates using the inverse variance weighted (IVW) method for both the traditional and spousal MR frameworks. Analyses used genetic instruments and effect estimates for NEB from Mathieson et al. (2023). Round markers indicate estimates using all NEB SNPs; triangular markers show results after excluding SNPs associated with EA. SNP-outcome associations were derived from UK Biobank spouse pairs, testing (i) male SNPs against female outcomes and (ii) female SNPs against male outcomes, both adjusted for ten male and ten female genetic PCs. Effect estimates represent the change in outcome per additional child. Outcomes that were inverse-normal transformed (including BMI, BMR, and BP) are presented in standard deviation (SD) units; while body fat percentage (BFP) are expressed in their original measurement units (percentage points). For all panels points indicate effect estimates and horizontal bars represent 95% confidence intervals (95% CI). BP; blood pressure. BFP; body fat percentage. BMR; basal metabolic rate. BMI; body mass index. CI; Confidence intervals. EA; Educational attainment. IVW; inverse variance weighted. MR; Mendelian Randomization. NEB; number of children ever born. PC; Principal component. SD; Standard deviation.

**Figure 5.**
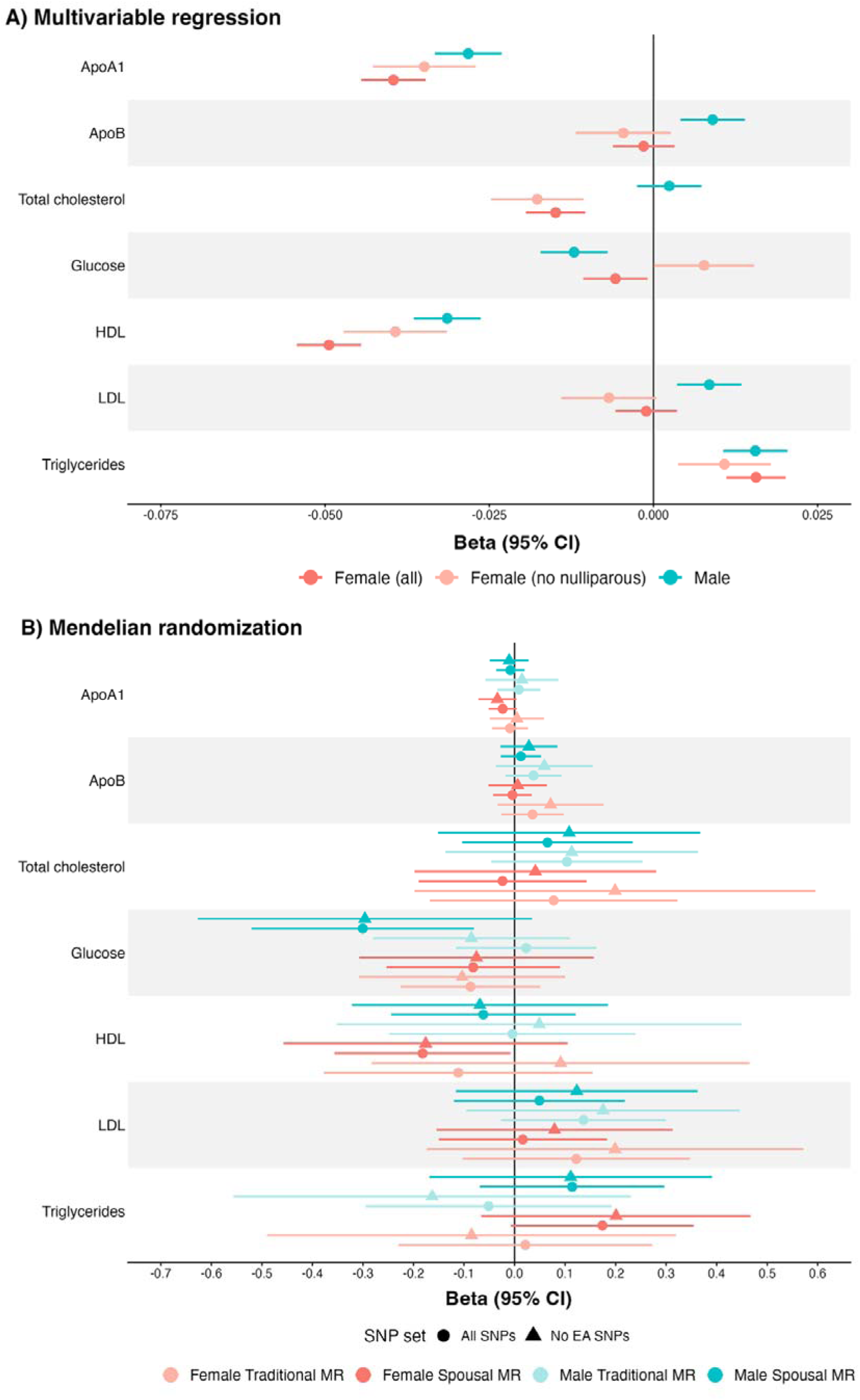
Associations between number of children ever born (NEB) and blood biomarkers. Panel A shows the results from the multivariable regression analyses including all females (N=172,122), all males (N=138,390) and the sensitivity analysis including only parous females (N=135,620). Estimates represent linear regression coefficients corresponding to the change in the outcome per additional child, expressed in standard deviation (SD) units. Analyses were corrected for year of birth, ten genome-wide genetic principal components (PCs), income and educational attainment (EA). Associations plotted are for the untransformed but normalized continuous outcomes. Estimates are presented for males (blue), all females (red) and only parous females (orange). Panel B shows the Mendelian randomization (MR) estimates using the inverse variance weighted (IVW) method for both the traditional and spousal MR frameworks. Analyses used genetic instruments and effect estimates for NEB from Mathieson et al. (2023). Round markers indicate estimates using all NEB SNPs; triangular markers show results after excluding SNPs associated with EA. SNP-outcome associations were derived from UK Biobank spouse pairs, testing (i) male SNPs against female outcomes and (ii) female SNPs against male outcomes, both adjusted for ten male and ten female genetic PCs. Effect estimates represent the change in outcome per additional child. Inverse-normal transformed outcomes (HDL, LDL, triglycerides, total cholesterol) are expressed in SD units; blood glucose is in mmol/L. ApoA1 and ApoB were natural log-transformed. Points indicate effect estimates; horizontal bars represent 95% confidence intervals (CI). BP; blood pressure. BFP; body fat percentage. BMR; basal metabolic rate. BMI; body mass index. CI; Confidence intervals. EA; Educational attainment. IVW; inverse variance weighted. MR; Mendelian Randomization. NEB; number of children ever born. PC; Principal component. SD; Standard deviation.

#### Sensitivity analyses restricted to parous females

Restricting analyses to parous females did not materially change most associations, with CIs overlapping between the main and sensitivity analyses for all traits except glucose, BMI, and BMR, for which associations strengthened (**Figures 3-5; Supplementary Table 10).** Sex heterogeneity emerged for blood glucose (P<0.05), whereas it was no longer evident for BMI and BMR (**Supplementary Table 10**, **Figures 4** and **5**). For socioeconomic traits, the inverse association with EA and the positive association with household income were both attenuated in magnitude (**Supplementary Table 10**).

### Causal inference analyses using MR

#### Instrument strength and IV assumptions

Instrument strength was adequate across all MR analyses (F-statistics >10), however the phenotypic variance in NEB jointly explained by all the IVs was low (R² = 0.19%; **Supplementary Table 9**). NEB-associated SNPs showed widespread pleiotropy in GWAS Atlas[53] and GWAS Catalog[54], including links to lipid levels, BP, and EA (**Supplementary Table 5**). In traditional MR, Cochran’s Q tests indicated substantial heterogeneity across SNPs, while MR-Egger intercepts suggested little evidence of directional pleiotropy (**Supplementary Table 13**). Heterogeneity was generally lower in the spousal MR analyses, possibly reflecting reduced power, with only weak evidence of heterogeneity across SNPs remaining for some traits (i.e. female BMI and BFP) (**Supplementary Table 16**). Spousal NEB GRS were uncorrelated (r=0.01, 95% CI: -0.00 to 0.02), although an individual’s NEB GRS was negatively associated with both their own and their partner’s EA (**Supplementary Tables 8** and **17**); these associations were no longer evident for partner EA after excluding EA-associated SNPs.

#### Causal evidence from traditional MR analyses

We observed evidence that higher NEB causally increased the odds of T2DM in females (P<6.25×10^-3^; OR = 1.71, 95% CI: 1.17-2.49), and males (OR = 1.28, 95% CI: 1.02-2.18; P=0.041); excluding EA-associated SNPs strengthened the effect in females while male estimates remained similar but with wider CIs (**Supplementary Table 12; Figure 3**). At nominal significance (P<0.05), higher NEB causally increased BMI in both sexes (β_females_ = 0.25, 95% CI: 0.07-0.43; β_males_ = 0.25, 95% CI: 0.07-0.43), female BMR (β = 0.24, 95% CI: 0.04-0.43), and male BFP (β = 0.80, 95% CI: 0.06-1.54), and decreased female systolic (β = - 0.14, 95% CI: -0.26 to -0.02) and diastolic BP (β = -0.17, 95% CI: -0.31 to -0.03) (**Supplementary Table 12; Figure 4**). Excluding EA SNPs attenuated effects for BMI, BMR, and BFP but not for BP (**Figures 4** and **5**).

No strong evidence of causal effects was found for other traits and heterogeneity tests showed no significant sex differences, though these tests may have limited power to detect modest differences in causal effects (**Supplementary Table 12**). Forest plots of SNP-specific causal effect estimates are presented in **Supplementary Figures 18-49**. MRlap-corrected estimates were broadly consistent with IVW results, though systolic BP, HDL, T2DM, and BMI showed larger effect magnitudes, suggesting downward bias due to winner’s curse (**Supplementary Table 14**).

#### Causal evidence from spousal MR

We observed evidence that higher NEB causally increased female BMI (P<6.25×10^-3^; β = 0.30, 95% CI: 0.10-0.50). At nominal significance (P<0.05), higher NEB increased female BMR (β = 0.23, 95% CI: 0.04-0.41), reduced female HDL (β = -0.18, 95% CI: -0.36 to -0.01) and lowered male blood glucose (β = -0.30, 95% CI: -0.52 to -0.08) (**Figures 4** and **5; Supplementary Table 15**). No clear causal effects were observed for other traits in either males or females.

In sensitivity analyses using recalculated SNP-exposure associations excluding UK Biobank to address sample overlap, we identified additional nominally significant effects (P<0.05) not observed in the primary analysis, including higher female triglycerides (β = 0.18, 95% CI: 0.01-0.34) and lower female ApoA1 (after EA SNP exclusion; β = -0.04, 95% CI: -0.076 to - 0.003) (**Supplementary Table 15**).

Exclusion of EA-associated SNPs generally attenuated causal effects (**Figures 3-5; Supplementary Tables 12** and **15**). Overall, traditional and spousal MR were largely directionally consistent, and CIs overlapped across all IVW estimates (**Figures 3-5**).

### Power calculations

At 80% power and assuming a (two-sided) type 1 error rate of α = 0.05, traditional MR could detect causal effects of β=0.07 (continuous traits; reflecting the expected change in standardised outcome per additional child) and OR=1.62 (binary traits; reflecting the relative change in odds of the outcome per additional child), with higher thresholds required in spousal MR (β=0.19; OR=2.21) (**Supplementary Figures 50-53**). As IVs explained only a small proportion of NEB variance (R²=0.19%), power was limited overall and particularly so in the spouse sample.

## Discussion

Observational associations between parity and adverse maternal cardiometabolic heath have been reported previously in the literature. However, these results have been inconsistent across studies and prone to confounding. By leveraging genetic and phenotypic data from the UK Biobank, we aimed to triangulate evidence from multivariable regression analyses and two complementary causal inference approaches to evaluate the potential causal effects of NEB on later-life cardiometabolic health.

Observationally, higher NEB was associated with cardiometabolic traits in both sexes after adjustment for income and EA. However, observational effect sizes were small, implying that any causal effects of NEB on cardiometabolic health are likely modest. Associations were largely consistent when restricting to parous females only, except for glucose, BMI, and BMR, where sex heterogeneity emerged for glucose but was no longer evident for BMI and BMR.

Traditional MR suggested potential causal effects of NEB on several outcomes but was limited by substantial SNP-level heterogeneity and pleiotropy. We therefore applied a spousal MR framework, which potentially minimises horizontal pleiotropy by instrumenting an individual’s NEB using their partner’s genotype - differing from prior spousal MR applications that have typically examined within-couple behavioural influences (e.g. whether an individual’s alcohol consumption influences their partner’s drinking behaviour)[59,60]. While other quasi-experimental approaches have studied parity effects on cardiometabolic outcomes (e.g. using firstborn child sex as an IV for maternal BP[40]), spousal MR offers a complementary strategy by leveraging large-scale genetic data.

Across methods, NEB was consistently associated with anthropometric and adiposity-related trait. Higher NEB was observationally and causally associated with increased female BMI and BMR across both MR approaches, with the increase in BMR potentially reflecting the energetic cost of maintaining higher body mass[61]. Although effects attenuated after removing EA-associated SNPs - suggesting partial mediation by EA - directionally consistent findings across methods support a causal effect. Interestingly, similar causal effect magnitudes for BMI and BMR were detected in males, suggesting that shared lifestyle or childrearing related behaviours (e.g., changes in physical activity, diet, or time constraints) may contribute to these effects.

As BMI is a major determinant of T2DM, CVD, dyslipidaemia, and metabolic syndrome[62–66], NEB-associated increases in BMI may partly explain the adverse lipid and glycaemic profile also observed in females. Multivariable regression and spousal MR analyses provided evidence of lower HDL cholesterol and ApoA1 and higher triglycerides with increasing NEB - indicative of an elevated triglyceride-to-HDL ratio, a recognised marker of insulin resistance [67–70]. Multivariable regression and traditional MR analyses further suggested increased odds of T2DM with consistent causal effect estimates in the spousal MR (though CIs were wider and crossed the null likely due to reduced power). Although the effect estimate for glucose was negative across all approaches in both sexes, this only survived multiple-testing correction in the spousal MR analysis of male glucose. This apparent inconsistency with the T2DM findings might be further explained by differences in phenotype definition: the fasting glucose GWAS used for traditional MR excluded diabetics, while the random glucose measure used in multivariable and spousal MR analyses included individuals with diabetes, whose glucose may be pharmacologically normalised.

We also observed evidence that higher NEB reduces female BP, a pattern consistent in both the multivariable regression and traditional MR analyses, with causal effects remaining robust even after excluding EA SNPs. This aligns with a large study (N= 444,611) using firstborn sex as an IV, which reported modest but sustained reductions in maternal BP (∼1 mmHg systolic, 0.35 mmHg diastolic per child) that attenuated over time[40]. Although a causal effect was not supported by spousal MR, given the older age of UKBB participants compared to the previous study (37-73 years vs 15-49), any incremental BP-lowering effects of higher NEB may have further waned, making subtle effects difficult to detect, particularly given the reduced statistical power of the spousal MR framework. Therefore, causal effects of NEB on BP cannot be ruled out, especially in younger females.

This study also has limitations. First, our analyses focused solely on NEB and did not encompass other aspects of reproductive history (e.g., stillbirths, miscarriages, pregnancy complications) that may have distinct long-term cardiometabolic effects. Additionally, for nulliparous females, it was not possible to determine whether childlessness was voluntary or due to underlying fertility issues, which may itself influence cardiometabolic health. Second, although we attempted to identify and account for pleiotropy, evidence of pleiotropic effects remained, indicating that both traditional and spousal MR estimates may still be biased, albeit to a lesser extent in spousal MR. Third, null findings in spousal MR may reflect limited statistical power, with small-to-modest effects likely gone undetected. Fourth, all analyses assumed linear relationships between NEB and outcomes; while evidence suggests some effects may be non-linear[15–22], current MR methods for detecting non-linear effects are prone to bias, precluding their assessment in this study[71–74]. Fifth, findings are limited to European individuals, and the spousal subsample differed from the broader UKBB sample in reproductive, demographic, and lifestyle characteristics, potentially limiting generalisability. Sixth, we were unable to disentangle the effects of childbearing from childrearing, which may differ between sexes given that females typically provide more direct care.

In conclusion, our study provides evidence of potential causal effects of NEB on later-life cardiometabolic health outcomes in women and men, with effects likely modest for most outcomes. We also illustrate a novel analytical approach, spousal MR, which leverages partner genotype to minimize horizontal pleiotropy. This approach is particularly well suited to exposures shared between partners, such as NEB, and requires genotyped spousal-pair datasets with outcome data available for both partners. As genotyped family datasets continue to grow, spousal MR offers a promising avenue for strengthening causal inference.

## Statements and Declarations

### Authors’ contributions

All authors contributed to the study conception and design. Data analysis was performed by Caroline Brito Nunes. Caroline Brito Nunes, Abigail Fraser, Gunn-Helen Moen, Alesha A. Hatton, and David M. Evans all contributed to the interpretation of the data. The project was conceptualized by David M. Evans. The first draft of the manuscript was written by Caroline Brito Nunes, and Abigail Fraser, Gunn-Helen Moen, Alesha A. Hatton, and David M. Evans critically revised previous versions of the manuscript. All authors read and approved the final manuscript.

### Ethics approval

The UKBB has ethical approval from the North West Multi-Centre Research Ethics Committee (MREC), which covers the UK, and all participants provided written informed consent (REC reference: 21/NW/0157). This research has been conducted using the UK Biobank resource under application number 53641. This project received ethical approval from the Institutional Human Research Ethics committee, University of Queensland (Approval Number 2019002705).

### Consent to participate

Informed consent was obtained from all individual participants included in the study.

### Data availability

UK Biobank (https://www.ukbiobank.ac.uk/) data is available to researchers upon application to the individual cohorts via their websites. All other data used are publicly available and referenced in the main text and/or Supplementary Table 6. Sex-specific GWAS summary statistics generated in this study have been deposited in the University of Queensland eSpace repository and are publicly available under unrestricted access: angina (https://rdm.uq.edu.au/files/a7b2ae66-624e-4090-8e6d-9baf5bfbdb77), myocardial infarction (https://rdm.uq.edu.au/files/63629e1a-1f1c-470d-93e7-c273548f581d), hypertension (https://rdm.uq.edu.au/files/de1df6d5-78f2-4512-a42f-4015d9837a6e) and basal metabolic rate (https://rdm.uq.edu.au/files/e33ec501-7069-4d51-b3bf-ca7f56211250).

## References

1. Sanghavi M, Rutherford JD. Cardiovascular Physiology of Pregnancy. Circulation. American Heart Association; 2014;130:1003–8. 10.1161/CIRCULATIONAHA.114.009029

2. Soma-Pillay P, Catherine N-P, Tolppanen H, Mebazaa A, Tolppanen H, Mebazaa A. Physiological changes in pregnancy. Cardiovasc J Afr. 2016;27:89–94. 10.5830/CVJA-2016-021

3. Biever C. Pregnancy’s true toll on the body: huge birth study paints most detailed picture yet. Nature. 2025;640:16–7. 10.1038/d41586-025-00959-7

4. Bar A, Moran R, Mendelsohn-Cohen N, Korem Kohanim Y, Mayo A, Toledano Y, et al. Pregnancy and postpartum dynamics revealed by millions of lab tests. Sci Adv. 2025;11:eadr7922. 10.1126/sciadv.adr7922

5. Jacobs MB, Kritz-Silverstein D, Wingard DL, Barrett-Connor E. The association of reproductive history with all-cause and cardiovascular mortality in older women: the Rancho Bernardo Study. Fertil Steril. 2012;97:118–24. 10.1016/j.fertnstert.2011.10.028

6. Cooper GS, Ephross SA, Weinberg CR, Baird DD, Whelan EA, Sandler DP. Menstrual and reproductive risk factors for ischemic heart disease. Epidemiology. 1999;10:255–9.

7. Gallagher LG, Davis LB, Ray RM, Psaty BM, Gao DL, Checkoway H, et al. Reproductive history and mortality from cardiovascular disease among women textile workers in Shanghai, China. Int J Epidemiol. 2011;40:1510–8. 10.1093/ije/dyr134

8. Jacobsen BK, Knutsen SF, Oda K, Fraser GE. Parity and total, ischemic heart disease and stroke mortality. The Adventist Health Study, 1976-1988. Eur J Epidemiol. 2011;26:711–8. 10.1007/s10654-011-9598-x

9. Koski-Rahikkala H, Pouta A, Pietiläinen K, Hartikainen A-L. Does parity affect mortality among parous women? J Epidemiol Community Health. 2006;60:968–73. 10.1136/jech.2005.044735

10. Steenland K, Lally C, Thun M. Parity and coronary heart disease among women in the American Cancer Society CPS II population. Epidemiology. 1996;7:641–3. 10.1097/00001648-199611000-00014

11. Naver KV, Lundbye-Christensen S, Gorst-Rasmussen A, Nilas L, Secher NJ, Rasmussen S, et al. Parity and risk of diabetes in a Danish nationwide birth cohort. Diabetic Medicine. 2011;28:43–7. 10.1111/j.1464-5491.2010.03169.x

12. Su H, Jiang C, Zhang W, Zhu F, Jin Y, Cheng K, et al. Parity and incident type 2 diabetes in older Chinese women: Guangzhou Biobank Cohort Study. Sci Rep. Nature Publishing Group; 2023;13:9504. 10.1038/s41598-023-36786-x

13. Musa IR, Osman OE, Adam I. The association between parity and type 2 diabetes mellitus: a cross-sectional, community-based study. BMC Endocrine Disorders. 2024;24:233. 10.1186/s12902-024-01767-2

14. Li P, Shan Z, Zhou L, Xie M, Bao W, Zhang Y, et al. MECHANISMS IN ENDOCRINOLOGY: Parity and risk of type 2 diabetes: a systematic review and dose-response meta-analysis. Eur J Endocrinol. 2016;175:R231–245. 10.1530/EJE-16-0321

15. Dior UP, Hochner H, Friedlander Y, Calderon-Margalit R, Jaffe D, Burger A, et al. Association between number of children and mortality of mothers: results of a 37-year follow-up study. Ann Epidemiol. 2013;23:13–8. 10.1016/j.annepidem.2012.10.005

16. Jaffe DH, Eisenbach Z, Manor O. The effect of parity on cause-specific mortality among married men and women. Matern Child Health J. 2011;15:376–85. 10.1007/s10995-010-0591-x

17. Lawlor DA, Emberson JR, Ebrahim S, Whincup PH, Wannamethee SG, Walker M, et al. Is the association between parity and coronary heart disease due to biological effects of pregnancy or adverse lifestyle risk factors associated with child-rearing? Findings from the British Women’s Heart and Health Study and the British Regional Heart Study. Circulation. 2003;107:1260–4. 10.1161/01.cir.0000053441.43495.1a

18. Parikh NI, Cnattingius S, Dickman PW, Mittleman MA, Ludvigsson JF, Ingelsson E. Parity and risk of later-life maternal cardiovascular disease. Am Heart J. 2010;159:215–221.e6. 10.1016/j.ahj.2009.11.017

19. Peters SA, Yang L, Guo Y, Chen Y, Bian Z, Millwood IY, et al. Parenthood and the risk of cardiovascular diseases among 0.5 million men and women: findings from the China Kadoorie Biobank. Int J Epidemiol. 2017;46:180–9. 10.1093/ije/dyw144

20. Khalid MEM. The effect of age, obesity and parity on blood pressure and hypertension in non-pregnant married women. J Family Community Med. 2006;13:103–7.

21. Li W, Ruan W, Lu Z, Wang D. Parity and risk of maternal cardiovascular disease: A dose– response meta-analysis of cohort studies. European Journal of Preventive Cardiology. 2019;26:592–602. 10.1177/2047487318818265

22. d’Errico A, Fontana D, Sacerdote C, Ardito C. Child rearing or childbearing? Risk of cardiovascular diseases associated to parity and number of children. BMC Public Health. 2024;24:272. 10.1186/s12889-023-17119-z

23. Li W, Wang Y, Shen L, Song L, Li H, Liu B, et al. Association between parity and obesity patterns in a middle-aged and older Chinese population: a cross-sectional analysis in the Tongji-Dongfeng cohort study. Nutr Metab (Lond). 2016;13:72. 10.1186/s12986-016-0133-7

24. Nehring I, Schmoll S, Beyerlein A, Hauner H, von Kries R. Gestational weight gain and long-term postpartum weight retention: a meta-analysis. Am J Clin Nutr. 2011;94:1225–31. 10.3945/ajcn.111.015289

25. Weng HH, Bastian LA, Taylor DH, Moser BK, Ostbye T. Number of children associated with obesity in middle-aged women and men: results from the health and retirement study. J Womens Health (Larchmt). 2004;13:85–91. 10.1089/154099904322836492

26. Hutchins F, Abrams B, Brooks M, Colvin A, Moore Simas T, Rosal M, et al. The Effect of Gestational Weight Gain Across Reproductive History on Maternal Body Mass Index in Midlife: The Study of Women’s Health Across the Nation. J Womens Health (Larchmt). 2020;29:148–57. 10.1089/jwh.2019.7839

27. Iversen DS, Kesmodel US, Ovesen PG. Associations between parity and maternal BMI in a population-based cohort study. Acta Obstetricia et Gynecologica Scandinavica. 2018;97:694–700. 10.1111/aogs.13321

28. Luoto R, Männistö S, Raitanen J. Ten-year change in the association between obesity and parity: results from the National FINRISK Population Study. Gend Med. 2011;8:399–406. 10.1016/j.genm.2011.11.003

29. Taghdir M, Alimohamadi Y, Sepandi M, Rezaianzadeh A, Abbaszadeh S, Mahmud FM. Association between parity and obesity: a cross sectional study on 6,447 Iranian females. J Prev Med Hyg. 2020;61:E476–81. 10.15167/2421-4248/jpmh2020.61.3.1430

30. Lee S-K, Sobal J, Frongillo EA, Olson CM, Wolfe WS. Parity and body weight in the United States: differences by race and size of place of residence. Obes Res. 2005;13:1263–9. 10.1038/oby.2005.150

31. Abrams B, Heggeseth B, Rehkopf D, Davis E. Parity and body mass index in US women: a prospective 25-year study. Obesity (Silver Spring). 2013;21:1514–8. 10.1002/oby.20503

32. Kim SA, Yount KM, Ramakrishnan U, Martorell R. The relationship between parity and overweight varies with household wealth and national development. Int J Epidemiol. 2007;36:93–101. 10.1093/ije/dyl252

33. He S, McArdle PF, Ryan KA, Daue M, Xu H, Barry KH, et al. Association of parity with body mass index and cardiometabolic risk in high-parous women. Menopause. 2023;30:703–8. 10.1097/GME.0000000000002194

34. Zoet GA, Paauw ND, Groenhof K, Franx A, Gansevoort RT, Groen H, et al. Association between parity and persistent weight gain at age 40–60 years: a longitudinal prospective cohort study. British Medical Journal Publishing Group; 2019 [cited 2025 Mar 6]; 10.1136/bmjopen-2018-024279

35. Lupton SJ, Chiu CL, Lujic S, Hennessy A, Lind JM. Association between parity and breastfeeding with maternal high blood pressure. American Journal of Obstetrics and Gynecology. 2013;208:454.e1–454.e7. 10.1016/j.ajog.2013.02.014

36. Haug EB, Horn J, Markovitz AR, Fraser A, Macdonald-Wallis C, Tilling K, et al. The impact of parity on life course blood pressure trajectories: the HUNT study in Norway. Eur J Epidemiol. 2018;33:751–61. 10.1007/s10654-018-0358-z

37. Magnus MC, Iliodromiti S, Lawlor DA, Catov JM, Nelson SM, Fraser A. Number of Offspring and Cardiovascular Disease Risk in Men and Women. Epidemiology. 2017;28:880–8. 10.1097/EDE.0000000000000712

38. Smith GD, Ebrahim S. “Mendelian randomization”: can genetic epidemiology contribute to understanding environmental determinants of disease? Int J Epidemiol. 2003;32:1–22. 10.1093/ije/dyg070

39. Ardissino M, Slob EAW, Carter P, Rogne T, Girling J, Burgess S, et al. Sex Specific Reproductive Factors Augment Cardiovascular Disease Risk in Women: A Mendelian Randomization Study. Journal of the American Heart Association. Wiley; 2023;12:e027933. 10.1161/JAHA.122.027933

40. Teufel F, Geldsetzer P, Sudharsanan N, Subramanyam M, Yapa HM, De Neve J-W, et al. The effect of bearing and rearing a child on blood pressure: a nationally representative instrumental variable analysis of 444 611 mothers in India. International Journal of Epidemiology. 2021;50:1671–83. 10.1093/ije/dyab058

41. Bycroft C, Freeman C, Petkova D, Band G, Elliott LT, Sharp K, et al. The UK Biobank resource with deep phenotyping and genomic data. Nature. Nature Publishing Group; 2018;562:203–9. 10.1038/s41586-018-0579-z

42. Tenesa A, Rawlik K, Navarro P, Canela-Xandri O. Genetic determination of height-mediated mate choice. Genome Biology. 2016;16:269. 10.1186/s13059-015-0833-8

43. Mathieson I, Day FR, Barban N, Tropf FC, Brazel DM, van Heemst D, et al. Genome-wide analysis identifies genetic effects on reproductive success and ongoing natural selection at the FADS locus. Nat Hum Behav. Nature Publishing Group; 2023;7:790–801. 10.1038/s41562-023-01528-6

44. Mahajan A, Taliun D, Thurner M, Robertson NR, Torres JM, Rayner NW, et al. Fine-mapping type 2 diabetes loci to single-variant resolution using high-density imputation and islet-specific epigenome maps. Nat Genet. 2018;50:1505–13. 10.1038/s41588-018-0241-6

45. Yang M-L, Xu C, Gupte T, Hoffmann TJ, Iribarren C, Zhou X, et al. Sex-specific genetic architecture of blood pressure. Nat Med. 2024;30:818–28. 10.1038/s41591-024-02858-2

46. Roshandel D, Lu T, Paterson AD, Dash S. Beyond apples and pears: sex-specific genetics of body fat percentage. Front Endocrinol (Lausanne). 2023;14:1274791. 10.3389/fendo.2023.1274791

47. Pulit SL, Stoneman C, Morris AP, Wood AR, Glastonbury CA, Tyrrell J, et al. Meta-analysis of genome-wide association studies for body fat distribution in 694 649 individuals of European ancestry. Hum Mol Genet. 2019;28:166–74. 10.1093/hmg/ddy327

48. Lagou V, Mägi R, Hottenga J-J, Grallert H, Perry JRB, Bouatia-Naji N, et al. Sex-dimorphic genetic effects and novel loci for fasting glucose and insulin variability. Nat Commun. 2021;12:24. 10.1038/s41467-020-19366-9

49. Graham SE, Clarke SL, Wu K-HH, Kanoni S, Zajac GJM, Ramdas S, et al. The power of genetic diversity in genome-wide association studies of lipids. Nature. 2021;600:675–9. 10.1038/s41586-021-04064-3

50. Kiewa J, Meltzer-Brody S, Milgrom J, Guintivano J, Hickie IB, Whiteman DC, et al. Comprehensive Sex-Stratified Genetic Analysis of 28 Blood Biomarkers and Depression Reveals a Significant Association between Depression and Low Levels of Total Protein in Females. Complex Psychiatry. 2024;10:19–34. 10.1159/000538058

51. Machiela MJ, Chanock SJ. LDlink: a web-based application for exploring population-specific haplotype structure and linking correlated alleles of possible functional variants. Bioinformatics. 2015;31:3555–7. 10.1093/bioinformatics/btv402

52. Mounier N, Kutalik Z. Bias correction for inverse variance weighting Mendelian randomization. Genet Epidemiol. 2023;47:314–31. 10.1002/gepi.22522

53. Tian D, Wang P, Tang B, Teng X, Li C, Liu X, et al. GWAS Atlas: a curated resource of genome-wide variant-trait associations in plants and animals. Nucleic Acids Research. 2020;48:D927–32. 10.1093/nar/gkz828

54. Sollis E, Mosaku A, Abid A, Buniello A, Cerezo M, Gil L, et al. The NHGRI-EBI GWAS Catalog: knowledgebase and deposition resource. Nucleic Acids Research. 2023;51:D977–85. 10.1093/nar/gkac1010

55. Bowden J, Davey Smith G, Burgess S. Mendelian randomization with invalid instruments: effect estimation and bias detection through Egger regression. Int J Epidemiol. 2015;44:512–25. 10.1093/ije/dyv080

56. Bowden J, Davey Smith G, Haycock PC, Burgess S. Consistent Estimation in Mendelian Randomization with Some Invalid Instruments Using a Weighted Median Estimator. Genet Epidemiol. 2016;40:304–14. 10.1002/gepi.21965

57. Hartwig FP, Davey Smith G, Bowden J. Robust inference in summary data Mendelian randomization via the zero modal pleiotropy assumption. Int J Epidemiol. 2017;46:1985–98. 10.1093/ije/dyx102

58. Hemani G, Zheng J, Elsworth B, Wade K, Baird D, Haberland V. The MR-Base platform supports systematic causal inference across the human phenome.

59. Richmond RC, Howe LJ, Heilbron K, Jones S, Liu J, Wang X, et al. Correlations in sleeping patterns and circadian preference between spouses. Commun Biol. Nature Publishing Group; 2023;6:1–14. 10.1038/s42003-023-05521-7

60. Howe LJ, Lawson DJ, Davies NM, St. Pourcain B, Lewis SJ, Davey Smith G, et al. Genetic evidence for assortative mating on alcohol consumption in the UK Biobank. Nat Commun. 2019;10:5039. 10.1038/s41467-019-12424-x

61. Henry CJK. Basal metabolic rate studies in humans: measurement and development of new equations. Public Health Nutr. 2005;8:1133–52. 10.1079/phn2005801

62. Chandrasekaran P, Weiskirchen R. The Role of Obesity in Type 2 Diabetes Mellitus-An Overview. Int J Mol Sci. 2024;25:1882. 10.3390/ijms25031882

63. Powell-Wiley TM, Poirier P, Burke LE, Després J-P, Gordon-Larsen P, Lavie CJ, et al. Obesity and Cardiovascular Disease: A Scientific Statement From the American Heart Association. Circulation. American Heart Association; 2021;143:e984–1010. 10.1161/CIR.0000000000000973

64. Khan SS, Ning H, Wilkins JT, Allen N, Carnethon M, Berry JD, et al. Association of Body Mass Index With Lifetime Risk of Cardiovascular Disease and Compression of Morbidity. JAMA Cardiol. 2018;3:280–7. 10.1001/jamacardio.2018.0022

65. Vekic J, Zeljkovic A, Stefanovic A, Jelic-Ivanovic Z, Spasojevic-Kalimanovska V. Obesity and dyslipidemia. Metabolism - Clinical and Experimental. Elsevier; 2019;92:71–81. 10.1016/j.metabol.2018.11.005

66. Han TS, Lean ME. A clinical perspective of obesity, metabolic syndrome and cardiovascular disease. JRSM Cardiovasc Dis. 2016;5:2048004016633371. 10.1177/2048004016633371

67. McLaughlin T, Abbasi F, Cheal K, Chu J, Lamendola C, Reaven G. Use of metabolic markers to identify overweight individuals who are insulin resistant. Ann Intern Med. 2003;139:802–9. 10.7326/0003-4819-139-10-200311180-00007

68. Pantoja-Torres B, Toro-Huamanchumo CJ, Urrunaga-Pastor D, Guarnizo-Poma M, Lazaro-Alcantara H, Paico-Palacios S, et al. High triglycerides to HDL-cholesterol ratio is associated with insulin resistance in normal-weight healthy adults. Diabetes Metab Syndr. 2019;13:382–8. 10.1016/j.dsx.2018.10.006

69. Gong R, Luo G, Wang M, Ma L, Sun S, Wei X. Associations between TG/HDL ratio and insulin resistance in the US population: a cross-sectional study. Endocr Connect. 2021;10:1502–12. 10.1530/EC-21-0414

70. Oliveri A, Rebernick RJ, Kuppa A, Pant A, Chen Y, Du X, et al. Comprehensive genetic study of the insulin resistance marker TG:HDL-C in the UK Biobank. Nature Genetics. Nature Publishing Group; 2024;56:212–21. 10.1038/s41588-023-01625-2

71. Burgess S, Wood AM, Butterworth AS. Mendelian randomisation and vitamin D: the importance of model assumptions - Authors’ reply. Lancet Diabetes Endocrinol. 2023;11:15–6. 10.1016/S2213-8587(22)00344-8

72. Hamilton FW, Hughes DA, Lu T, Kutalik Z, Gkatzionis A, Tilling K, et al. Non-linear Mendelian randomization: evaluation of effect modification in the residual and doubly-ranked methods with simulated and empirical examples. Eur J Epidemiol. 2025;40:631–47. 10.1007/s10654-025-01208-x

73. Burgess S. Towards more reliable non-linear Mendelian randomization investigations. Eur J Epidemiol. 2024;39:447–9. 10.1007/s10654-024-01121-9

74. Butler-Laporte G, Richards JB. Mendelian randomisation and vitamin D: the importance of model assumptions. The Lancet Diabetes & Endocrinology. Elsevier; 2023;11:14–5. 10.1016/S2213-8587(22)00342-4

