## Supplementary Figures for "Investigating the potential causal relationship between parity and long-term maternal cardiometabolic health outcomes using Mendelian randomization": Supplementary Figures.docx

**Supplementary Materials**

### **1.0 Spousal pair identification in the UKBB**

Firstly, we identified potential spousal pairs in the UKBB using the returned dataset by Tenesa et al. (2016)^42^, which included 90,232 pairs matched on co-location, sex, birth dates, household relationships, and parental ages. These variables were used to identify opposite-sex couples from other cohabiting individuals: age differences <10 years increase the likelihood of a spousal relationship, household relationship codes help confirm the relationship within the household (i.e. individuals reported the household composition to be either 'Husband, wife or partner' or 'Husband, wife or partner' and 'Son and/or daughter (include step-children)’), and parental ages (which either equalled the age or the age at death if the parent was not alive) of either mother or father differed help rule out parent-child pairs. After excluding pairs reported as spouses who were genetically related (IBD > 0.08), suggesting that they were family members, 90,117 pairs remained.

Because co-location information groups individuals by street address, this approach may incorrectly classify unrelated opposite-sex individuals living in shared buildings (e.g., apartment complexes or retirement homes) as spouses. To address this limitation, spouses were further matched using five additional household variables. The ability of these and other candidate household variables to distinguish true spousal pairs was evaluated in 1,027 genetically confirmed parent-offspring trios within UK Biobank, in which the two parents are highly likely to represent a true spousal pair at least at some point in time. **Supplementary Table 1** provides a comparison of spousal pairs identified in the Tenesa et al. dataset with couples present in the parent-offspring trios file. **Supplementary Figure 1** shows the distribution of matches and mismatches across all investigated household variables in these confirmed couples and indicates which variables were used in Tenesa et al. (2016) and which were selected in the present study.

Based on this validation, we selected five household variables (number of vehicles, own/rent accommodation, accommodation type, household income, and Townsend Deprivation Index) which showed low mismatch rates when identifying the parents of 694 out of the 1,027 genetically confirmed parent-offspring trios. **Supplementary Table 2** summarises mismatch patterns across these five household variables in the Tenesa et al. spousal pairs file. **Supplementary Figure 2** presents the distribution of matches and mismatches for these five variables among couples present in both the Tenesa et al. dataset and the trios file, including a comparison of strict versus relaxed matching criteria for household income. **Supplementary Figure 3** shows the distribution of matches and mismatches for spouses identified in genetically confirmed parent–offspring trios who were not present in the Tenesa et al. returned dataset, illustrating reasons for non-identification such as changes in household composition or missing household relationship information.

Identical responses for all five household variables were required for a pair of individuals to be classified as a spousal pair, with the exception of household income, for which one-category differences or missing data were permitted. This refinement yielded 76,883 spousal pairs. We then excluded non-European individuals and pairs in which either spouse was genetically related to an individual in another spousal pair (IBD > 0.08), resulting in 68,073 spousal pairs. Finally, analyses were restricted to 53,325 spousal pairs who reported the same number of children and had available phenotype data.

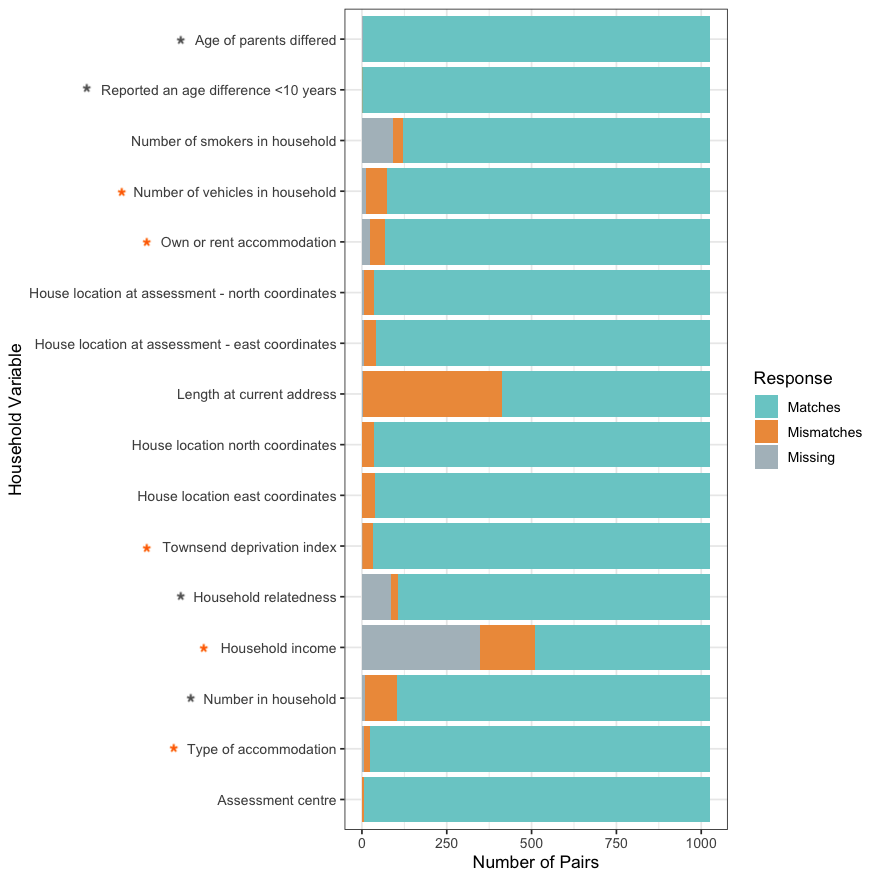

**Supplementary Figure 1.** *Distribution of matches and mismatches for variables reported by couples in genetically confirmed parent-offspring trios in the UK Biobank (N=1,027).* Values reported had to be identical to be considered a ‘match’. In the returned dataset from Tenesa et al., 2015, criteria used to identify spouses included household relatedness, age difference <10 years, number of individuals in household, parents age differed (including age at death). Prior to further matching spouses, we investigated several household variables to determine the most optimal criteria to match potential spouses in the UK Biobank. Variables highlighted with the grey asterisks were used to select spousal pairs in Tenesa et al., 2015. Orange asterisks indicate the household variables used to further select spousal pairs in our study.

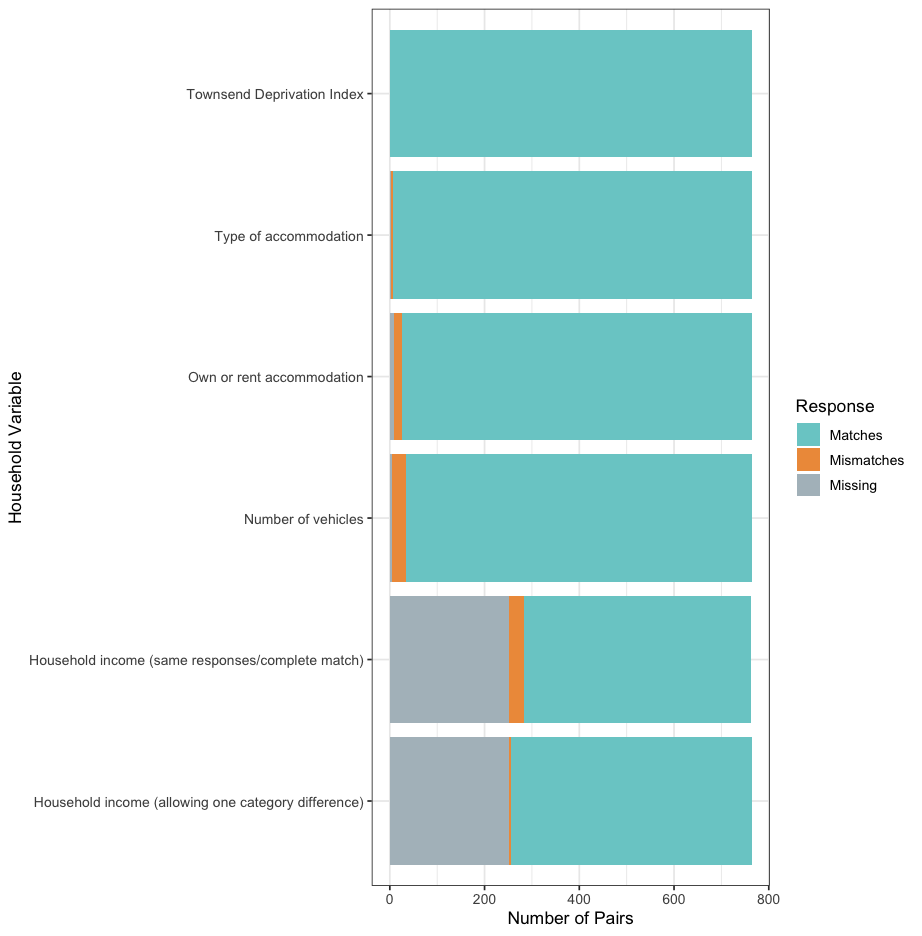

**Supplementary Figure 2.** *Distribution of the matches and mismatches for the five household variables across couples that were present both in the trios file and in the Tenesa et al. dataset (N=764).* Comparison was made with the returned dataset from Tenesa et al., 2015 after excluding pairs originally reported as couples but who appeared to be genetically related (N= 90,117). Couples in confirmed parent-offspring trios were identified based on genetic relatedness (N=1,027). The five household variables presented were used to further match spouses. We also provide a comparison of matches and mismatches for income when requiring identical responses and allowing a one category difference. Spouses had to report identical responses for it to be considered a “match”, with the exception of income which had an alternative criterion where one-category differences or missing data were permitted.

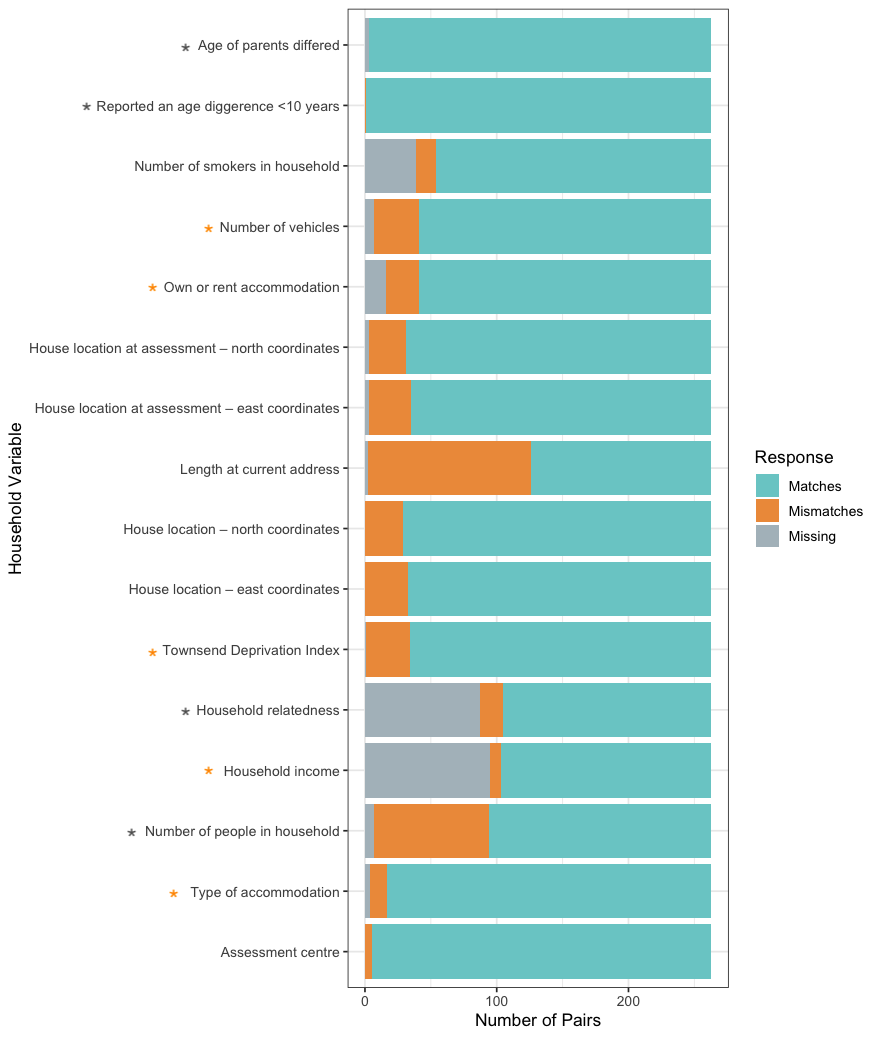

**Supplementary Figure 3.** *Distribution of the matches and mismatches for spouses that were not present in the returned dataset from Tenesa et al., 2015 but were present in the confirmed parent-offspring trios (N=263).* Comparison was made with the returned dataset from Tenesa et al., 2015 after excluding related individuals (N= 90,117). Couples in confirmed parent-offspring trios were identified based on genetic relatedness (N=1,027). Spouses had to report identical responses for it to be considered a “match”. Couples might not have been identified in Tenesa et al., 2015 due to discrepancies in *i)* their responses for number of individuals in household and house location, potentially suggesting they might have had a child but are no longer living together, and *ii)* missing information for household relatedness.

### **2.0 Sex-stratified GWAS in the UKBB**

Genome-wide association analyses (GWAS) were conducted using data from the UKBB, stratified by sex, for traits without publicly available sex-specific GWAS results. These traits included basal metabolic rate (BMR) and cardiovascular disease (CVD) risk traits (angina, myocardial infarction, and hypertension). Definition of cases and controls for binary outcomes are provided in **Supplementary Table 3**. Analyses included only European individuals. Individuals who withdrawn their consent from having their data stored by UKBB, and those failing standard quality checks (e.g., due to sex mismatch, high heterozygosity, missingness, sex chromosome aneuploidy) were excluded.

For binary traits (angina, myocardial infarction, and hypertension), GWAS were performed using REGENIE v4.0, which applies a two-step whole-genome regression framework to account for population structure and relatedness, modelling genetic effects via ridge regression followed by single-variant association testing. For BMR, GWAS were performed using BOLT-LMM v2.4.1, which uses a linear mixed model framework. For BMR, which was not normally distributed, an inverse normal transformation was applied prior to analysis using BOLT-LMM (**Supplementary Figure 7**). All GWAS models were adjusted for the first five principal components, birth year, and genotyping batch. Manhattan and QQ plots are provided below (**Supplementary Figure 5-8**).

The LD-score regression software (https://github.com/bulik/ldsc) was implemented to estimate SNP heritability (SNP h^2^) for CVD traits and BMR. In the supplementary figures below, we also report the SNP-based heritability and LDSC regression intercepts.

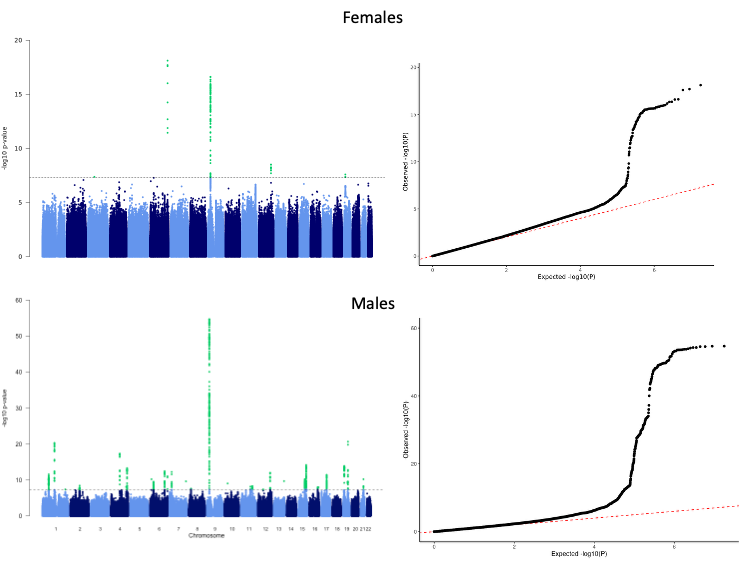

**Supplementary Figure 4.** *Manhattan and QQ plots for the sex-stratified GWAS of angina.* Results are shown for females (top) and males (bottom). Calculated lambda was 1.08 for females and 1.11 males. Female GWAS had 10,602 cases and 157,695 controls. Male GWAS had 18,572 cases and 114,964 controls. SNP-based heritability estimates on the observed scale were 0.042 (SE = 0.004) in females and 0.098 (SE = 0.006) in males. The LDSC intercepts were 1.036 (SE = 0.008) for females and 1.028 (SE = 0.009) for males. SE; Standard error.

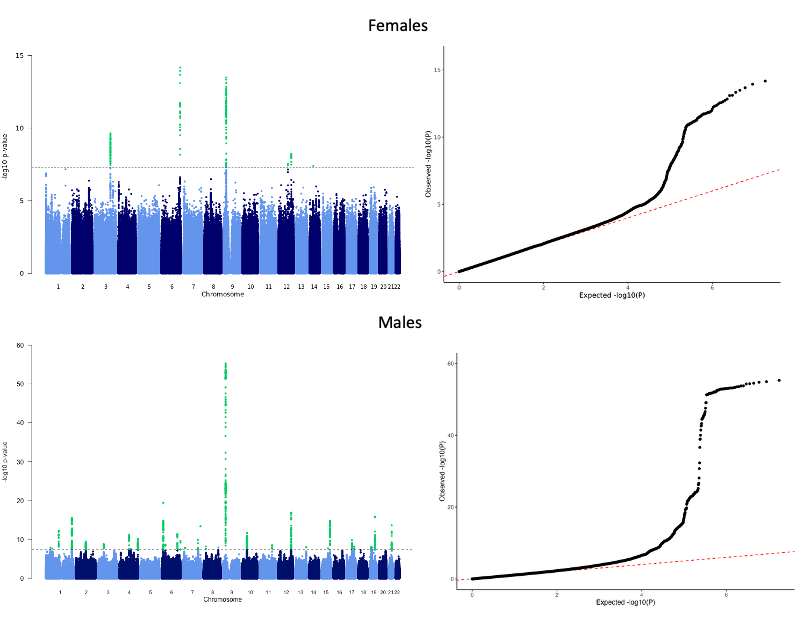

**Supplementary Figure 5.** *Manhattan and QQ plots for the sex-stratified GWAS of myocardial infarction.* Results are shown for females (top) and males (bottom) Calculated lambda was 1.05 for females and 1.10 males. Female GWAS had 4,761 cases and 159,332 controls. Male GWAS had 13,862 cases and 116,139 controls. SNP-based heritability estimates on the observed scale were 0.022 (SE = 0.003) in females and 0.091 (SE = 0.006) in males. The LDSC intercepts were 1.025 (SE = 0.007) for females and 1.034 (SE = 0.009) for males. SE; Standard error.

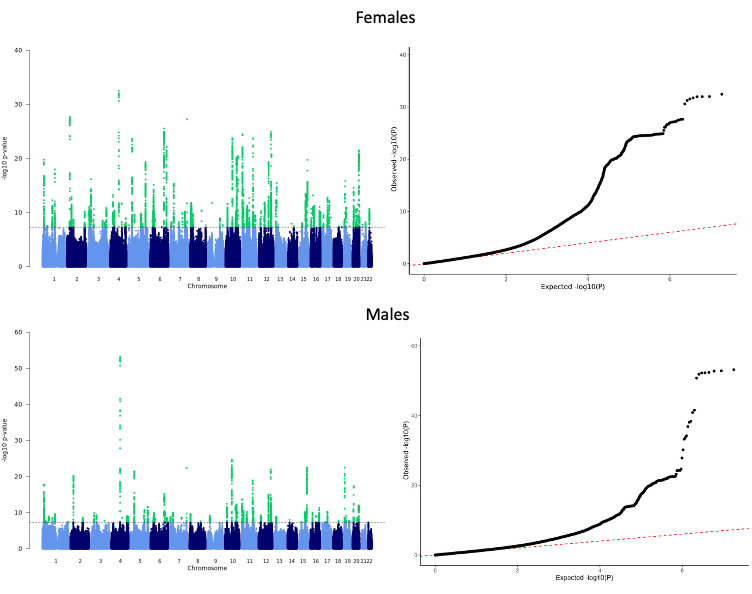

**Supplementary Figure 6.** *Manhattan and QQ plots for the sex-stratified GWAS of hypertension.* Results are shown for females (top) and males (bottom). Calculated lambda was 1.17 for females and 1.16 males. Female GWAS had 75,967 cases and 139,250 controls. Male GWAS had 82,642 cases and 97,697 controls. SNP-based heritability estimates on the observed scale were 0.134 (SE = 0.007) in females and 0.130 (SE = 0.006) in males. The LDSC intercepts were 1.05 (SE = 0.013) for females and 1.048 (SE = 0.011) for males. SE; Standard error.

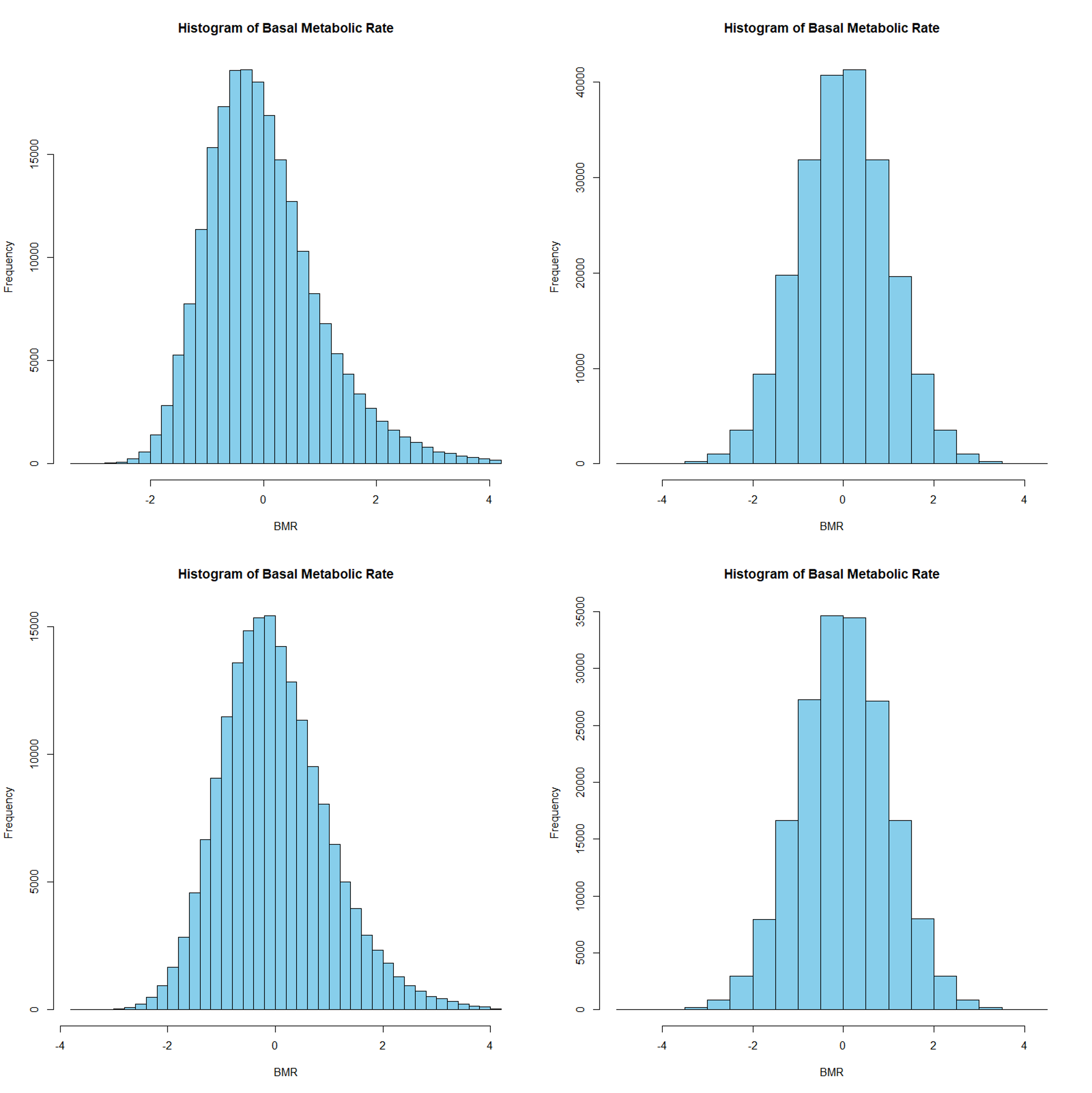

D)

C)

A)

B)

B)

B)

**Supplementary Figure 7.** *Distribution of basal metabolic rate prior and after inverse normal transformation.* Distribution is presented for females (top) and males (bottom), before (A and C) and after transformation (B and D).

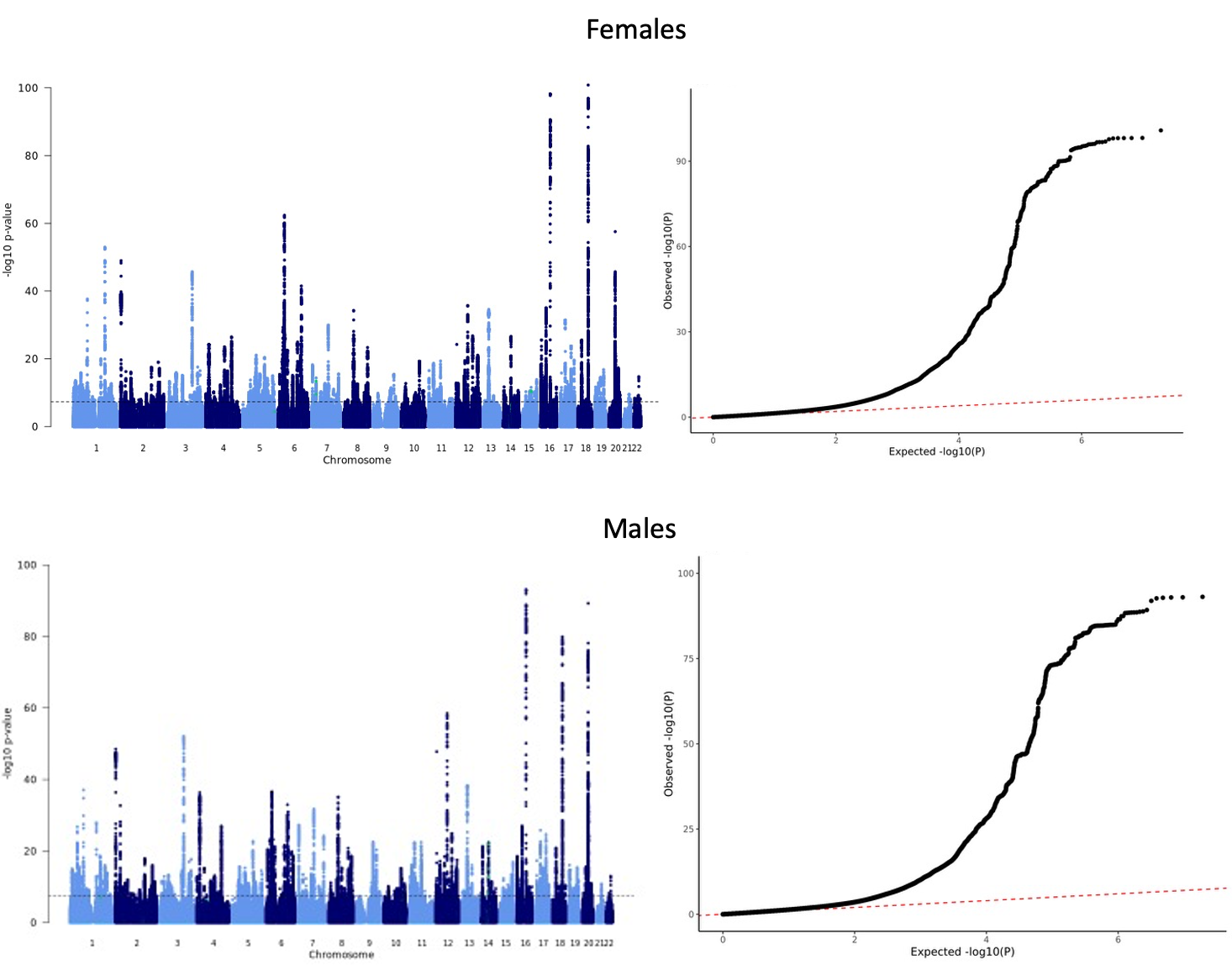

**Supplementary Figure 8.** *Manhattan and QQ plots for the sex-stratified GWAS of basal metabolic rate (BMR).* Results are shown for females (top) and males (bottom). Calculated lambda was 1.37 for females and 1.37 males. Female GWAS had 212,061. Male GWAS had 179,217. SNP-based heritability estimates on the observed scale were 0.300 (SE = 0.010) in females and 0.325 (SE = 0.013) in males. The LDSC intercepts were 1.07 (SE = 0.016) for females and 1.08 (SE = 0.012) for males. SE; Standard error.

### **3.0 Calculating SNP-NEB estimates without UKBB**

In the UKBB, we regressed NEB on the identified SNPs to obtain the UKBB-specific effect estimates and respective standard errors (SE). We assessed these associations in a sample similar to the one in Mathieson et al. 2023, excluding related and non-European individuals, those who did not pass quality control checks, and including individuals based on age (females should be at least 45 years of age and males at least 55). Linear regressions were used to assess SNP-exposure association in females (for female-specific loci), males (for male-specific loci) and in a sex-combined sample (for loci identified in the sex-combined meta-analysis). Models included ten principal components (PCs), birth year minus 1900 and its polynomials (square and cube) to capture nonlinear birth cohort effects and, in the sex-combined analyses, interaction terms between birth year (and its polynomials) with sex were included to adjust for potential gender differences in the cohort effects.

We then recalculated the meta-analysis SE (Equation I) and computed effect estimates accordingly (Equation II) (**Supplementary Table 7**).

(I) ${SE}_{noUKBB}=\sqrt{\frac{1}{\left( \frac{1}{{{SE}_{meta}}^{2}} \right)-\left( \frac{1}{{{SE}_{UKBB}}^{2}} \right)}}$

Where SE_noUKBB_ is the standard error removing the effect of the UKBB, SE_meta_ is the standard error from the meta-analysis, SE_UKBB_ is the standard error from the UK Biobank.

(II) $\beta_{noUKBB}=\left( \beta_{meta}\times\left( \frac{1}{{{SE}_{UKBB}}^{2}}+\frac{1}{{{SE}_{noUKBB}}^{2}} \right)-\frac{\beta_{UKBB}}{{{SE}_{UKBB}}^{2}} \right)\times{{SE}_{noUKBB}}^{2}$

Where β_noUKBB_ is the adjusted beta coefficient, β_meta_ is the beta coefficient from the meta-analysis, β_UKBB_ is the beta coefficient from the UKBB analysis, SE_noUKBB_ is the standard error removing the effect of the UKBB, SE_meta_ is the standard error from the meta-analysis, and SE_UKBB_ is the standard error from the UKBB.

### **4.0 Principal component analyses**

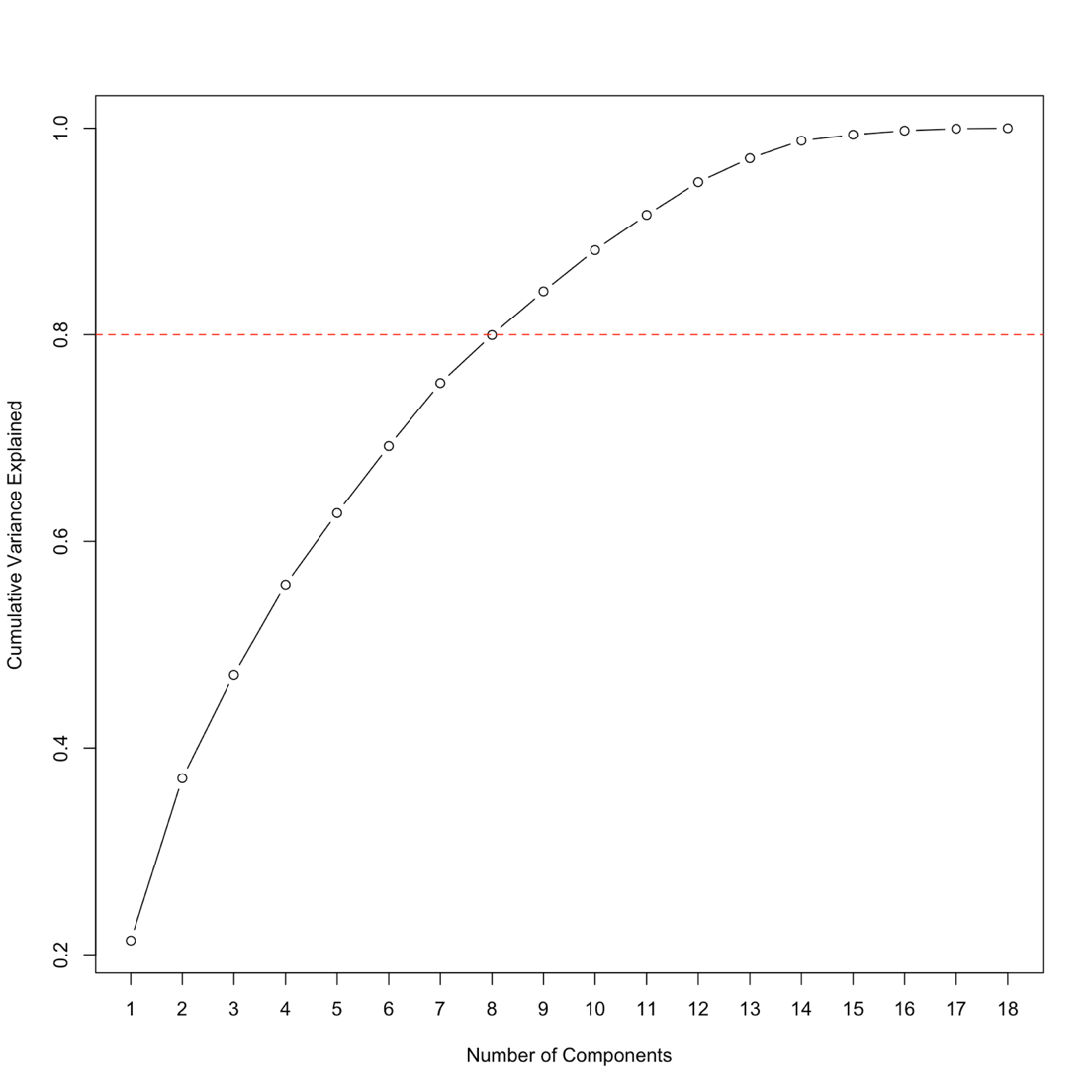

**Supplementary Figure 9.** *Scree plot showing the cumulative variance explained by principal components in the principal component analysis (PCA) in the UK Biobank female subset.* PCA was conducted across 95,197 females in UK Biobank with complete outcome data to determine the number of PCs that explained >80% of the variance shared between outcomes (represented by the red dashed line). A total of eight PCs accounted for 80% of the covariance between traits, suggesting an approximate multiple-testing corrected p-value threshold of P < 6.25$\times$10^-3^ for statistical significance in female-specific analyses involving all relevant outcomes (i.e. the traditional epidemiological analyses, individual-level MR and spousal MR analyses).

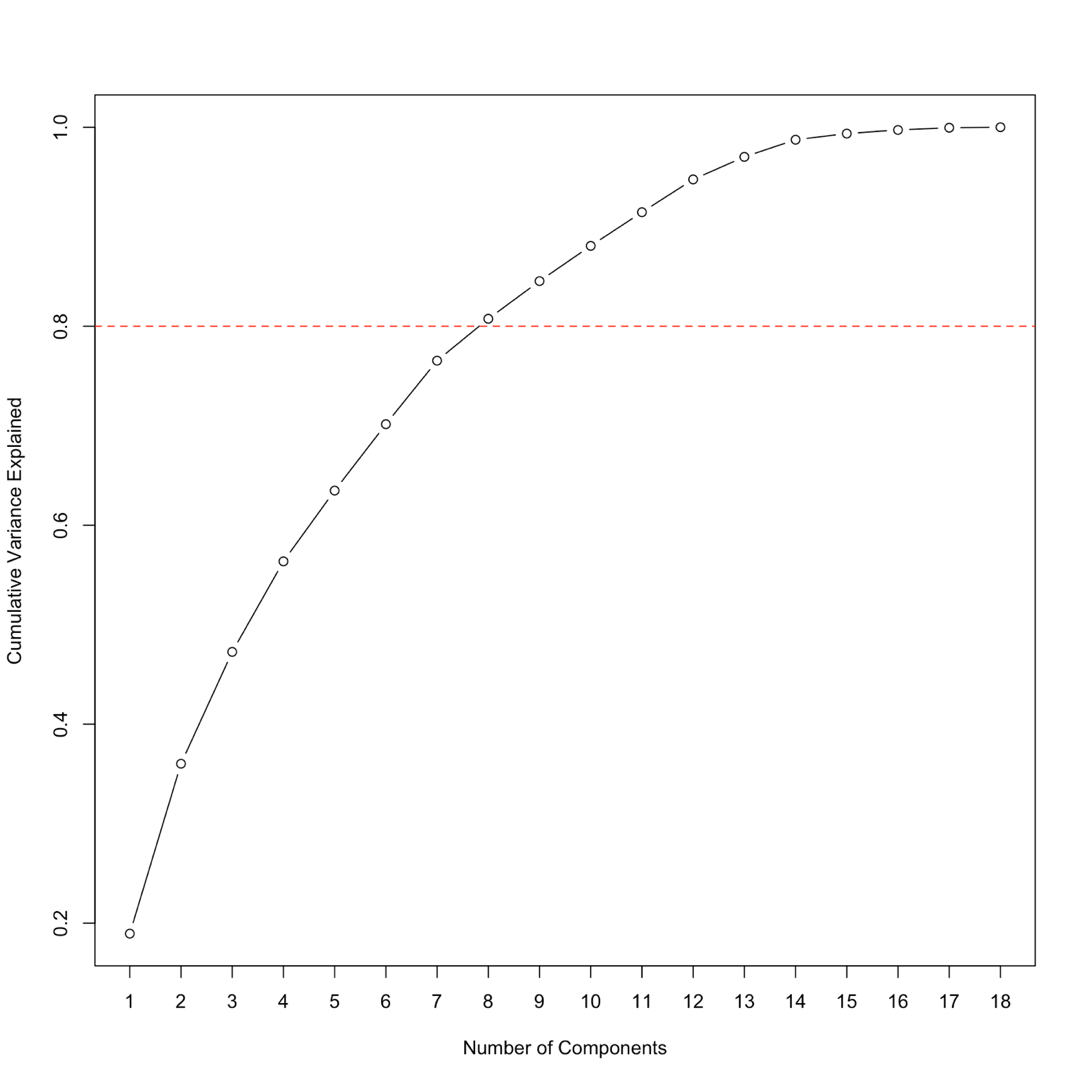

**Supplementary Figure 10.** *Scree plot showing the cumulative variance explained by principal components in the principal component analysis (PCA) in the UK Biobank male subset.* PCA was conducted across 78,711 males in UK Biobank with complete outcome data to determine the number of PCs that explained >80% of the variance shared between outcomes (represented by the red dashed line). A total of eight PCs accounted for more than 80% of the covariance between traits, suggesting an approximate multiple-testing corrected p-value threshold of P < 6.25$\times$10^-3^ for statistical significance in male-specific analyses involving all relevant outcomes (i.e. the traditional epidemiological analyses, individual-level MR and spousal MR analyses).

### **5.0 Results for the traditional observational epidemiological analyses**

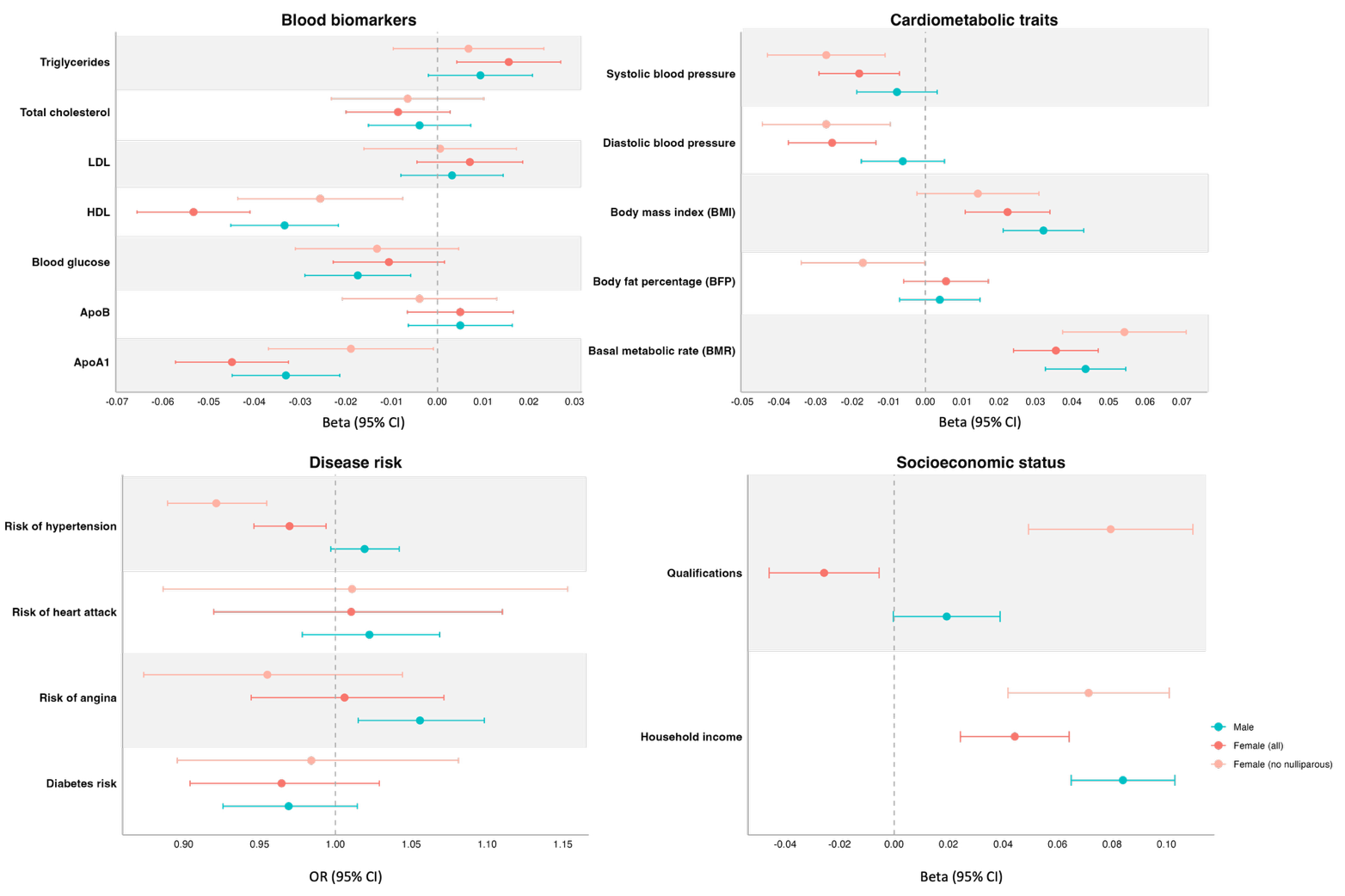

**Supplementary Figure 11.** *Forest plot showing the observational associations between NEB cardiometabolic traits in the UK Biobank spouse subset using the adjusted model.* Plot shows analyses for females and males in the spousal pair subset (N=53,237) as well as sensitivity analyses including only parous females in the spousal pair dataset (N= 47,791). The estimates (beta or odds ratio) and 95% confidence intervals (95% CI) are plotted in the x-axis for each trait. Adjusted analyses were corrected for year of birth, ten principal components, income and educational attainment. For income, covariates only included year of birth, ten principal components and educational attainment. For qualifications, covariates only included year of birth, ten principal components and income. NEB; Number of children ever born.

### **6.0 Sample characteristics**

We examined the characteristics of the spouse subset within UK Biobank to assess how this group may differ from the broader sample (i.e. individual participant sample) as spouses may represent a non-random subgroup, potentially selected on shared social, behavioural, and reproductive factors.

##### *6.1 Birth year*

Compared with the full UKBB cohort, the spouse subset was generally older, with fewer individuals born after 1954 and a concentration of birth years in the early 1940s to early 1950s (**Supplementary Figure 12**).

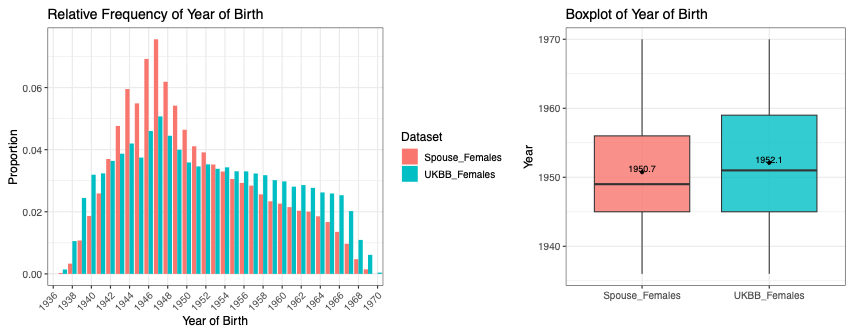

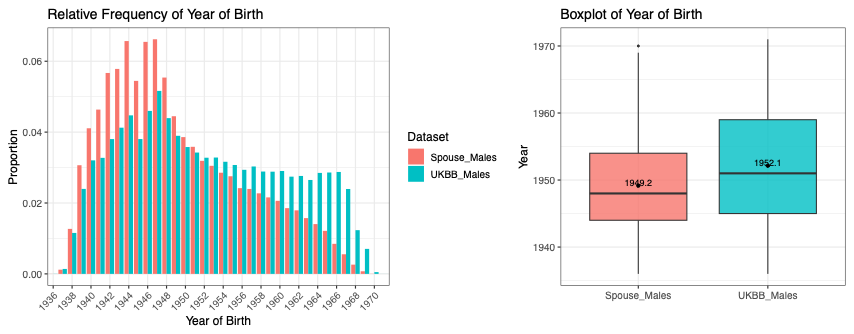

**Supplementary Figure 12.** *Distribution of birth year in the individual participant sample and the spousal subset.* The left panels show bar plots of the relative frequency distributions of birth year in females (top) and males (bottom) across the UK Biobank (UKBB) and spouse subsets, two independent datasets. The right panels present box plots displaying the distributions of these counts by dataset, with numbers indicating the mean values.

##### *6.2 Reproductive history*

Individuals in the spousal subset sample had more children than individuals in the individual participant sample (**Supplementary Figure 13**). In females, 57.7% had two children compared with 40.2% in UKBB overall, and fewer were nulliparous (10.2% vs. 21.2%). Similar patterns were observed in males, with 57.7% reporting two children versus 36.9% in UKBB males, and fewer reporting no children (10.2% vs. 24.6%).

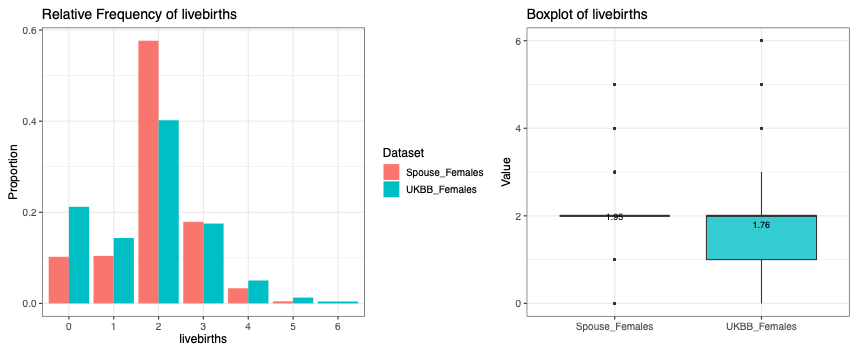

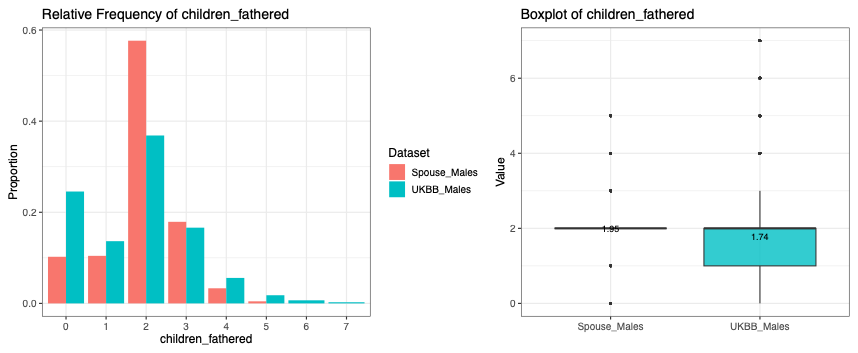

**Supplementary Figure 13.** *Distribution of number of children ever born (NEB) in the individual participant sample and the spousal subset.* The panels display the bar plots of the relative frequency distributions of live births (females at the top) and children fathered (males at the bottom) across the UK Biobank (UKBB) and spouse subsets, two independent datasets. Individuals in the spousal subset sample were matched on NEB.

##### *6.3 Socioeconomic characteristics*

Education levels differed slightly between the two samples (**Supplementary Figure 14**). Among females, a higher proportion in the individual participant sample held university degrees compared with females in the spouse subset (33.2% vs. 29.9%), whereas among males, university degree attainment was higher in the spouse subset than in the individual participant sample (36.5% vs. 34.5%).

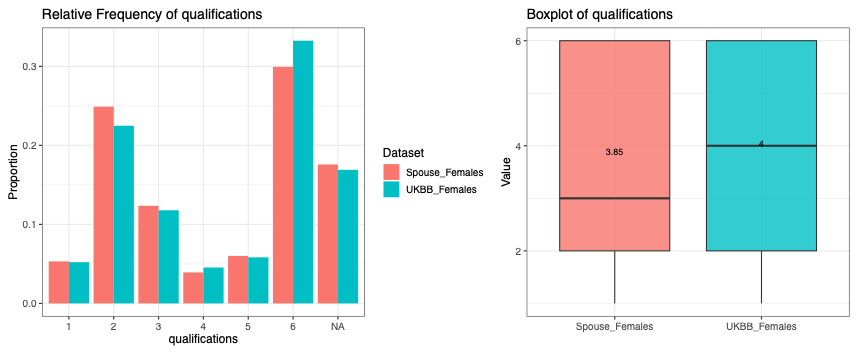

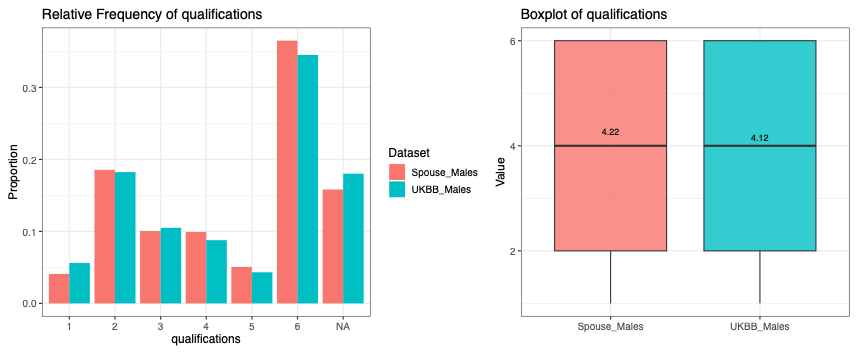

**Supplementary Figure 14.** *Distribution of educational qualifications in the individual participant sample and the spousal subset.* The panels display the bar plots of the relative frequency distributions of educational qualifications in females (top) and males (bottom) across the UK Biobank (UKBB) and spouse subsets, two independent datasets. For analysis, educational qualifications were coded so that higher values reflected higher levels of attainment: 6 - College or University degree, 5 - Other professional qualifications (e.g., nursing, teaching), 4 - A levels/AS levels or equivalent, 3 - O levels/GCSEs or equivalent, 2 - CSEs or equivalent, and 1 - NVQ or HND or HNC or equivalent.

Spouses also tended to report higher household income (**Supplementary Figure 15**). Only 11.4% of spouse females and 12.4% of spouse males fell in the lowest income category, compared to 22.5% and 19.8% in UKBB females and males, respectively. Spouses were more likely to fall into middle-income categories (particularly categories 3 and 4). Compared to males, females were more likely to have missing income data, often reporting ‘prefer not to say’ or ‘don’t know’.

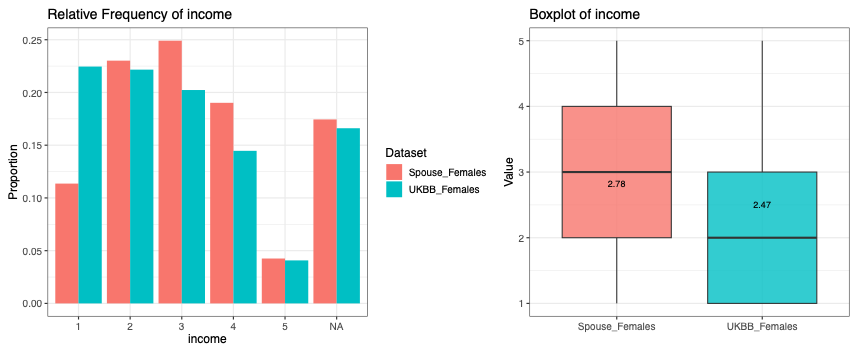

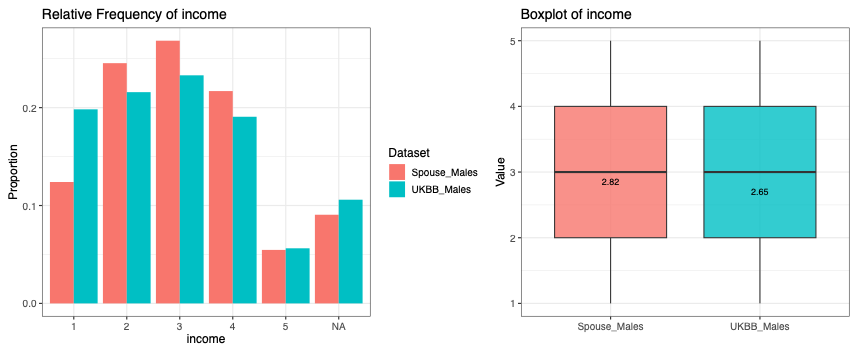

**Supplementary Figure 15.** *Distribution of number of household income in the individual participant sample and the spousal subset.* The panels display the bar plots of the relative frequency distributions of household income in females (top) and males (bottom) across the UK Biobank (UKBB) and spouse subsets, two independent datasets. Household income was coded as an ordinal variable ranging from 1 to 5, with higher values indicating higher income brackets: 1 - Less than £18,000; 2 - £18,000 to £30,999; 3 - £31,000 to £51,999; 4 - £52,000 to £100,000; and 5 - Greater than £100,000.

##### *6.4 Lifestyle factors*

Smoking prevalence differed between samples (**Supplementary Figure 16**). Females in the spousal group were less likely to be current smokers and more likely to have never smoked compared the females in the individual participant sample (4.5% vs. 9.6% and 65.0% vs. 58.0%, respectively). Similar patterns were observed among males (current smoking: 7.4% vs. 13.5%; never smoked: 50.1% vs. 48.5%).

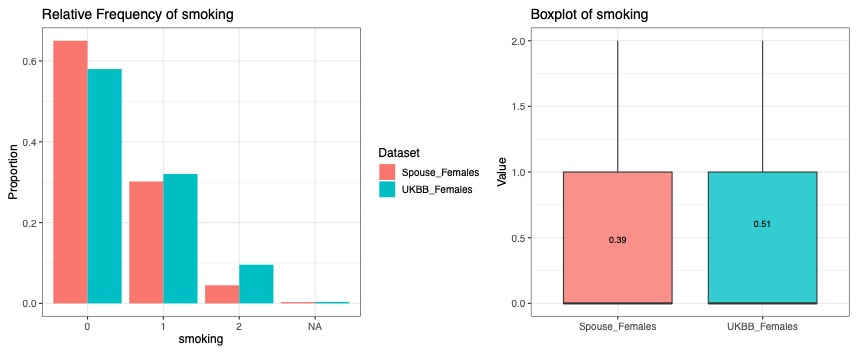

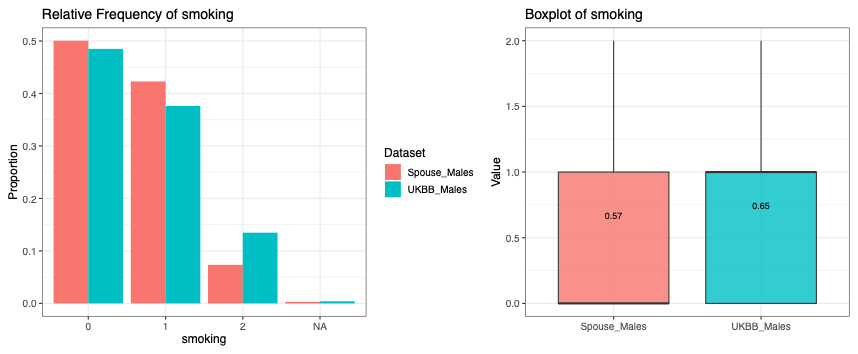

**Supplementary Figure 16.** *Distribution of smoking status in the individual participant sample and the spousal subset.* The left panels show bar plots of the relative frequency distributions of smoking status in females (top) and males (bottom) across the UK Biobank (UKBB) and spouse subsets, two independent datasets. Smoking status was coded as 0 - Never smoked, 1 – Previous smoker and 2 – Current smoker.

Daily or near-daily drinking (category 1) was slightly more common in spouse males (26.8% vs. 24.1%) and spouse females (16.5% vs. 15.7%) compared with the broader UKBB sample (**Supplementary Figure 17**). Individuals in the spouse subset were also more likely to report moderate drinking (3-4 or 1-2 times per week, categories 2-3) compared to the individual participant sample (**Supplementary Figure 17**).

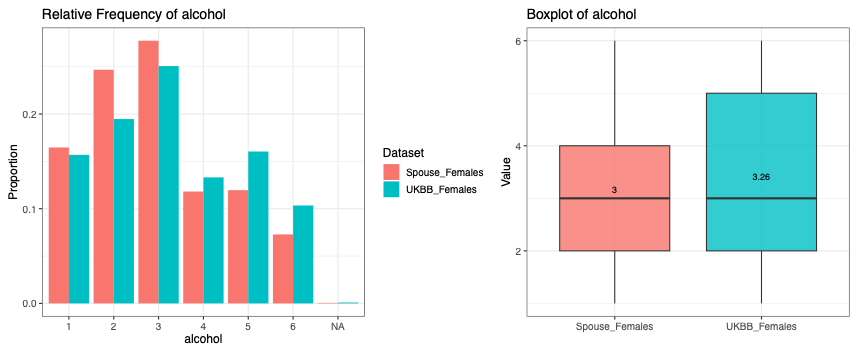

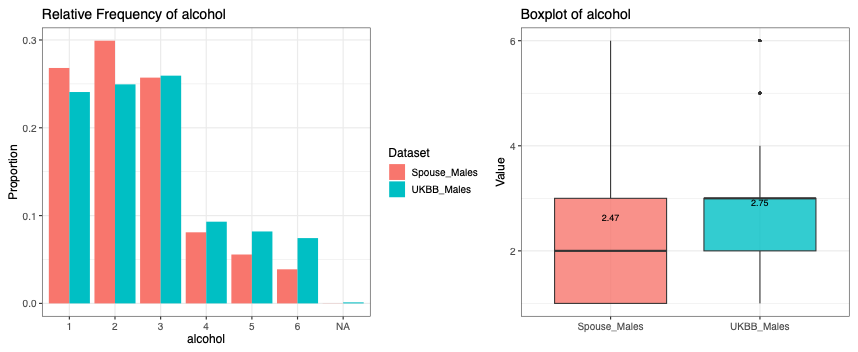

**Supplementary Figure 17.** *Distribution of alcohol consumption frequency in the individual participant sample and the spousal subset.* The left panels show bar plots of the relative frequency distributions of alcohol consumption frequency in females (top) and males (bottom) across the UK Biobank (UKBB) and spouse subsets, two independent datasets. Alcohol consumption frequency was coded as: 1 - Daily or almost daily, 2 - Three or four times a week, 3 - Once or twice a week, 4 - One to three times a month, 5 - Special occasions only, 6 - Never.

### **7.0 Traditional Two-sample MR**

#### 7.1 Female-specific analyses

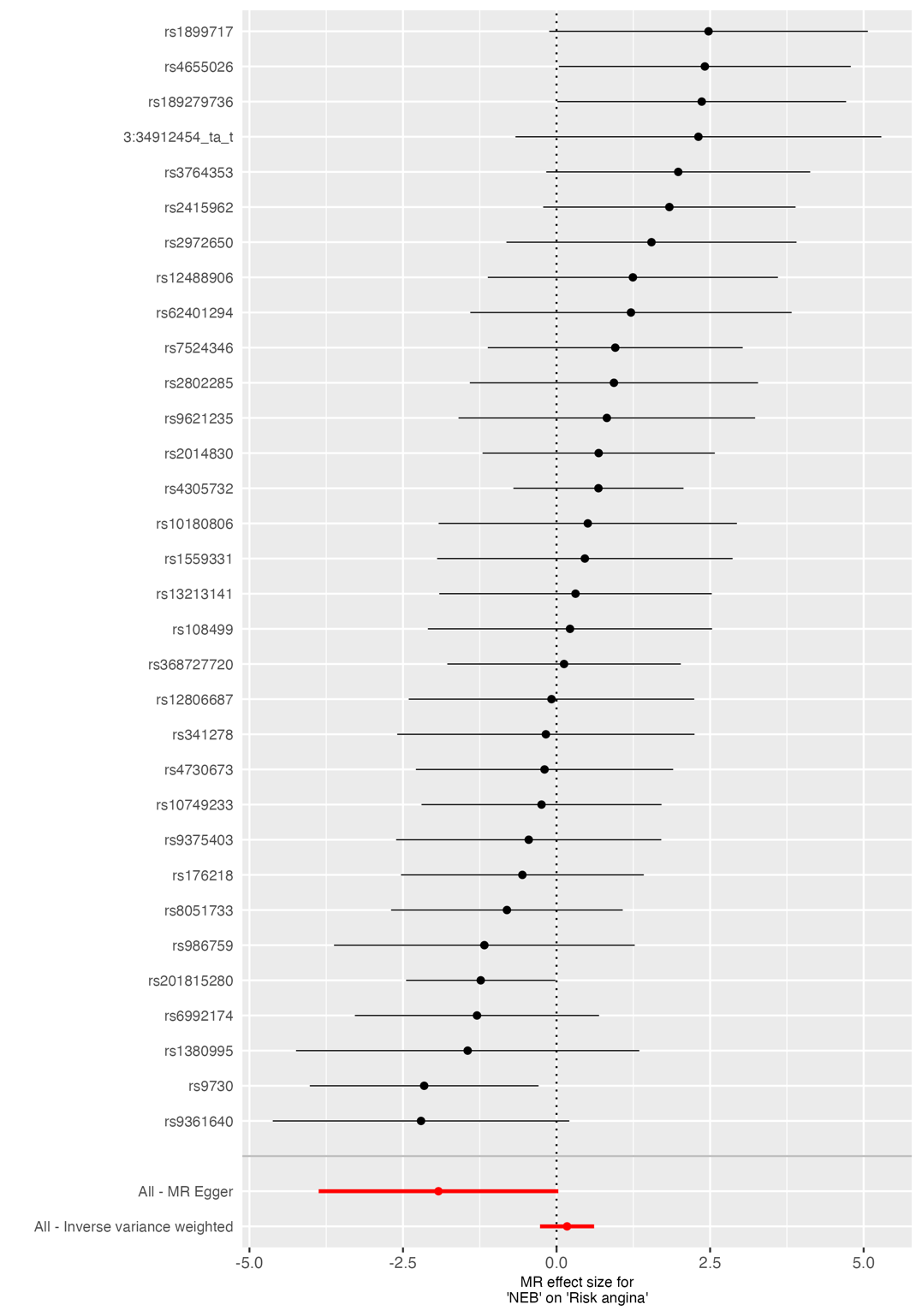

**Supplementary Figure 18.** *Forest plot of SNP-specific causal estimates for the effect of NEB on angina.* Forest plot displaying individual Wald ratio estimates and 95% confidence intervals for each SNP used as an instrumental variable for parity in the two-sample MR analysis. Summary causal estimates derived from both inverse-variance weighted (IVW) and MR Egger methods are shown at the bottom of the plot. Variation in SNP-specific effects reflects heterogeneity across instruments. NEB; Number of children ever born.

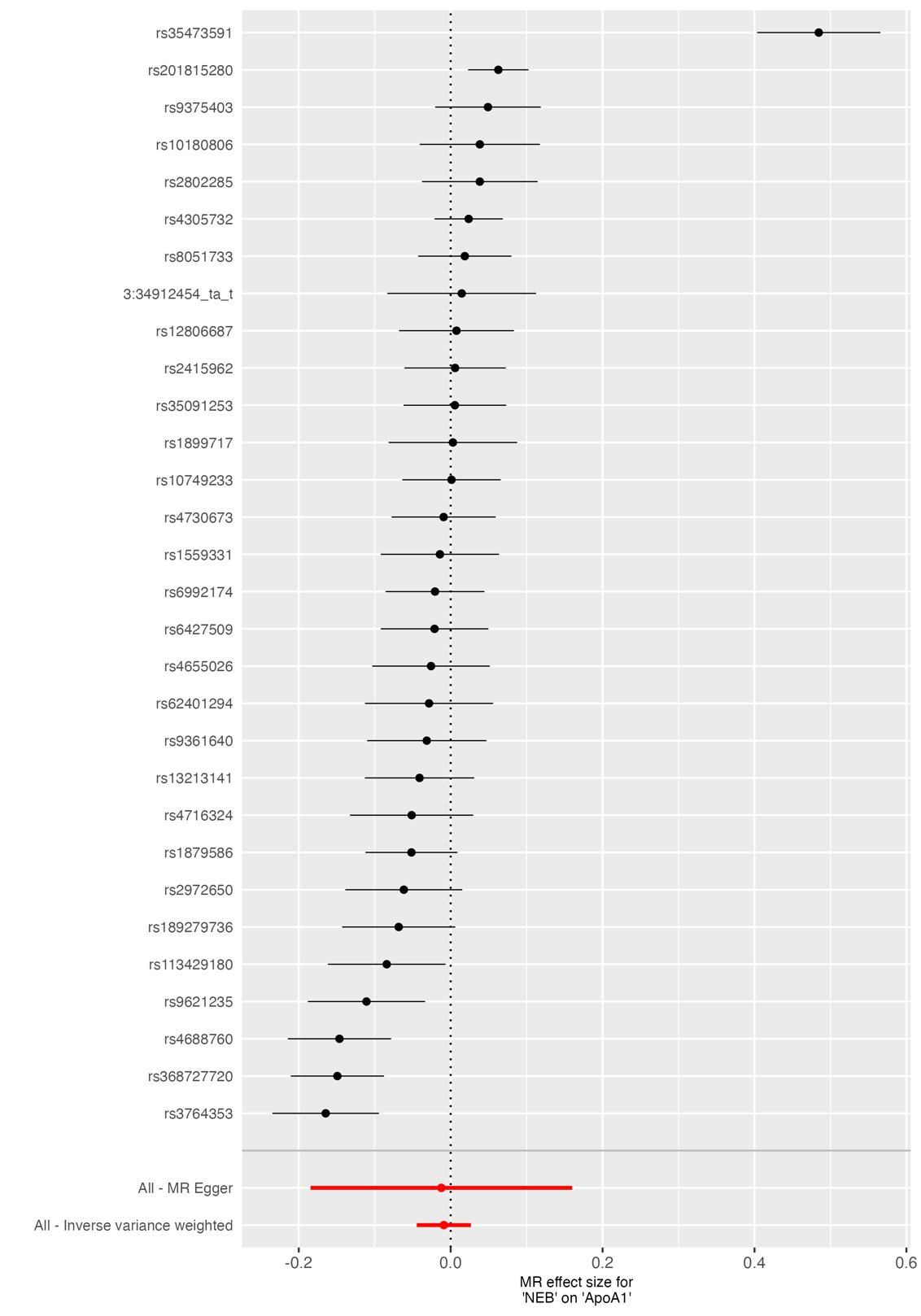

**Supplementary Figure 19.** *Forest plot of SNP-specific causal estimates for the effect of NEB on ApoA1.* Forest plot displaying individual Wald ratio estimates and 95% confidence intervals for each SNP used as an instrumental variable for parity in the two-sample MR analysis. Summary causal estimates derived from both inverse-variance weighted (IVW) and MR Egger methods are shown at the bottom of the plot. Variation in SNP-specific effects reflects heterogeneity across instruments. NEB; Number of children ever born.

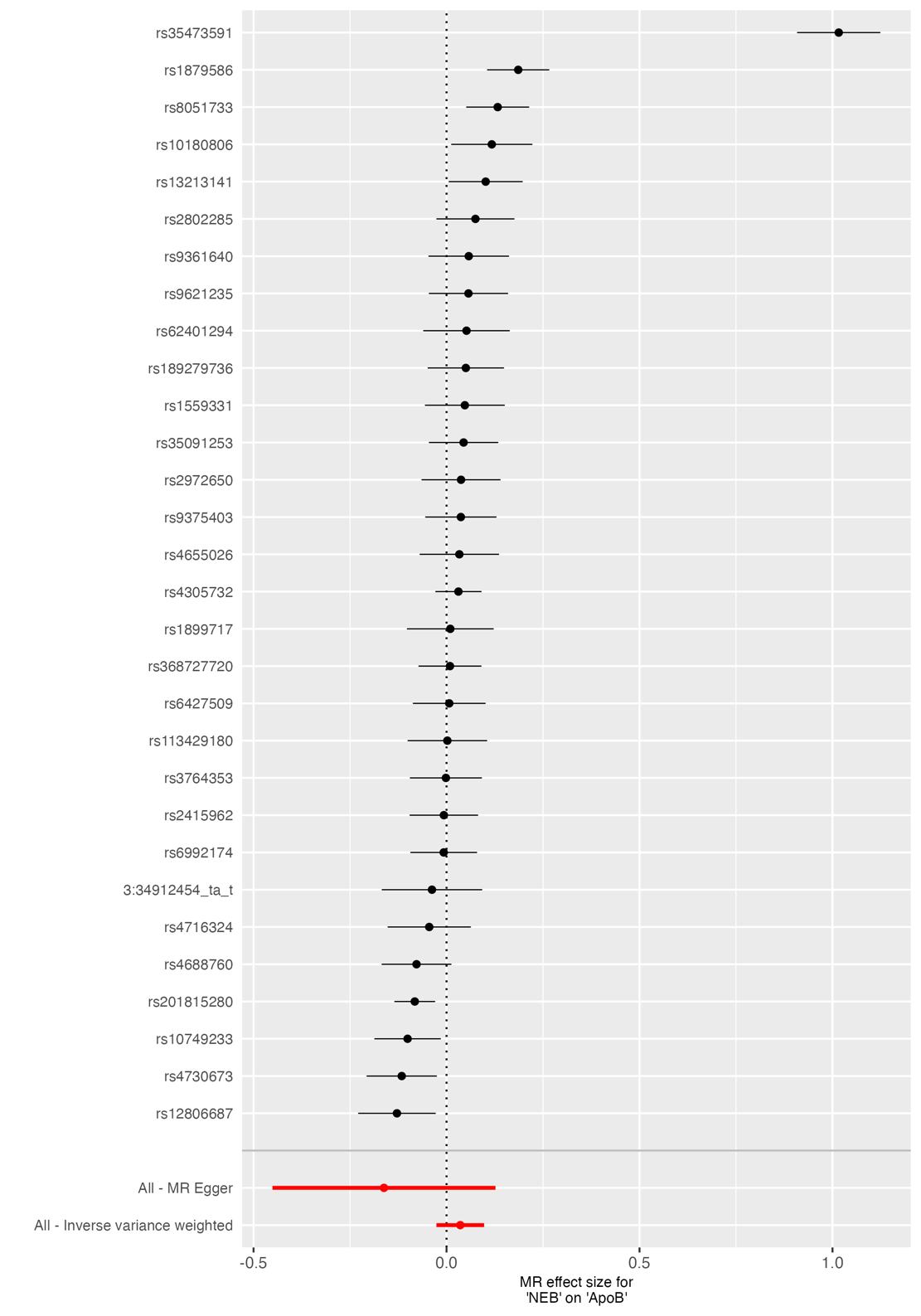

**Supplementary Figure 20.** *Forest plot of SNP-specific causal estimates for the effect of NEB on ApoB.* Forest plot displaying individual Wald ratio estimates and 95% confidence intervals for each SNP used as an instrumental variable for parity in the two-sample MR analysis. Summary causal estimates derived from both inverse-variance weighted (IVW) and MR Egger methods are shown at the bottom of the plot. Variation in SNP-specific effects reflects heterogeneity across instruments. NEB; Number of children ever born.

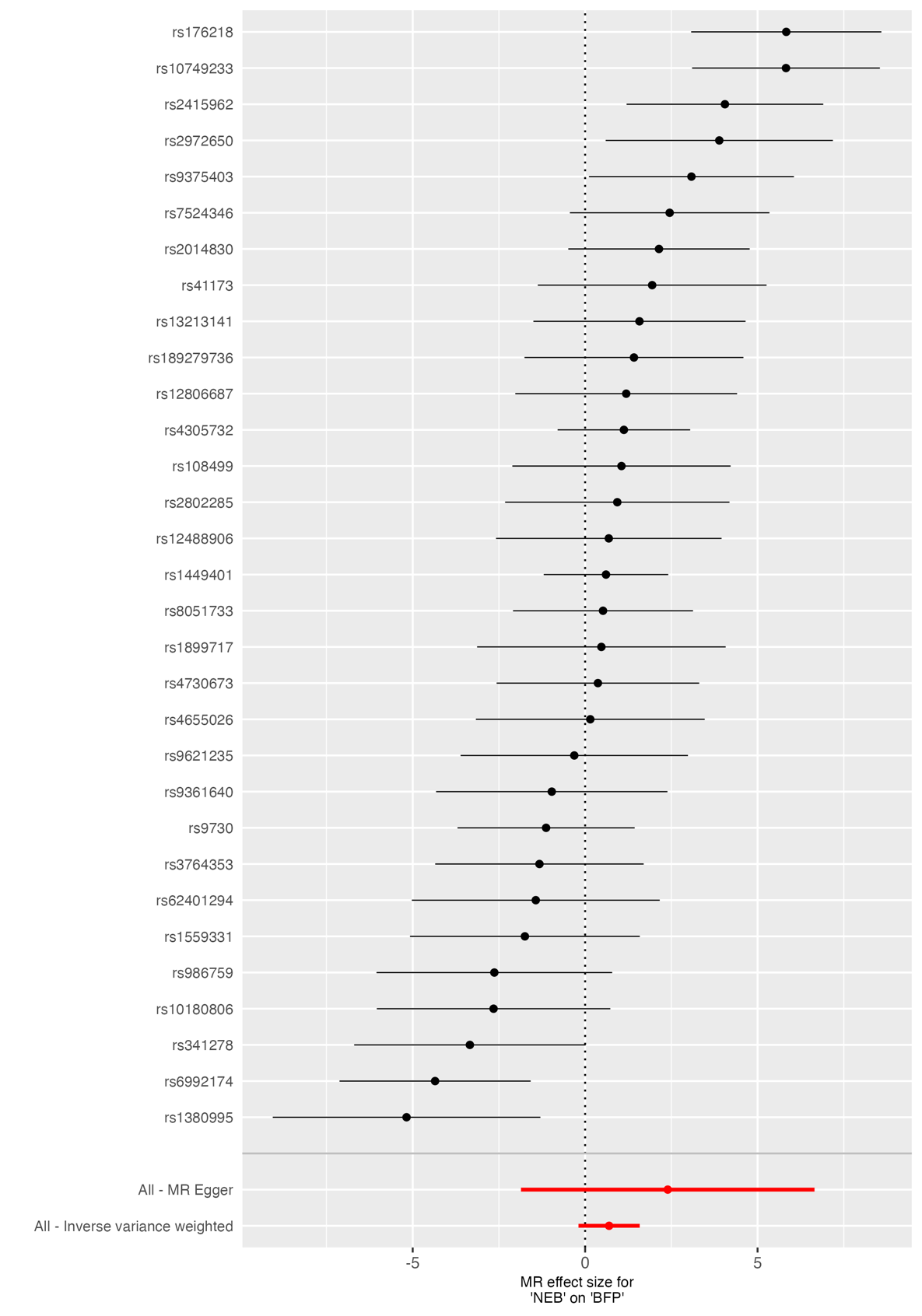

**Supplementary Figure 21.** *Forest plot of SNP-specific causal estimates for the effect of NEB on body fat percentage (BFP).* Forest plot displaying individual Wald ratio estimates and 95% confidence intervals for each SNP used as an instrumental variable for parity in the two-sample MR analysis. Summary causal estimates derived from both inverse-variance weighted (IVW) and MR Egger methods are shown at the bottom of the plot. Variation in SNP-specific effects reflects heterogeneity across instruments. NEB; Number of children ever born.

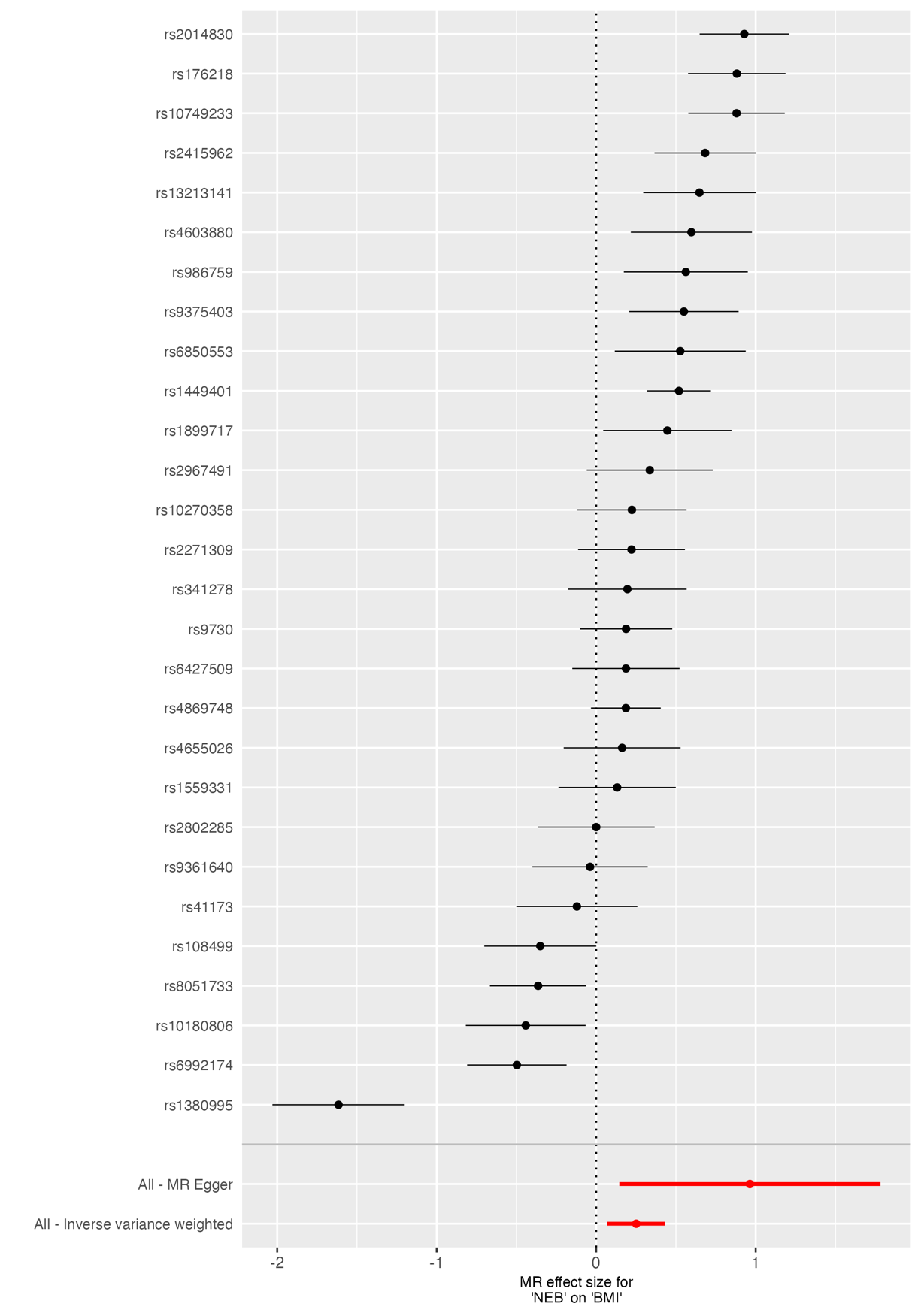

**Supplementary Figure 22.** *Forest plot of SNP-specific causal estimates for the effect of NEB on body mass index (BMI).* Forest plot displaying individual Wald ratio estimates and 95% confidence intervals for each SNP used as an instrumental variable for parity in the two-sample MR analysis. Summary causal estimates derived from both inverse-variance weighted (IVW) and MR Egger methods are shown at the bottom of the plot. Variation in SNP-specific effects reflects heterogeneity across instruments. NEB; Number of children ever born.

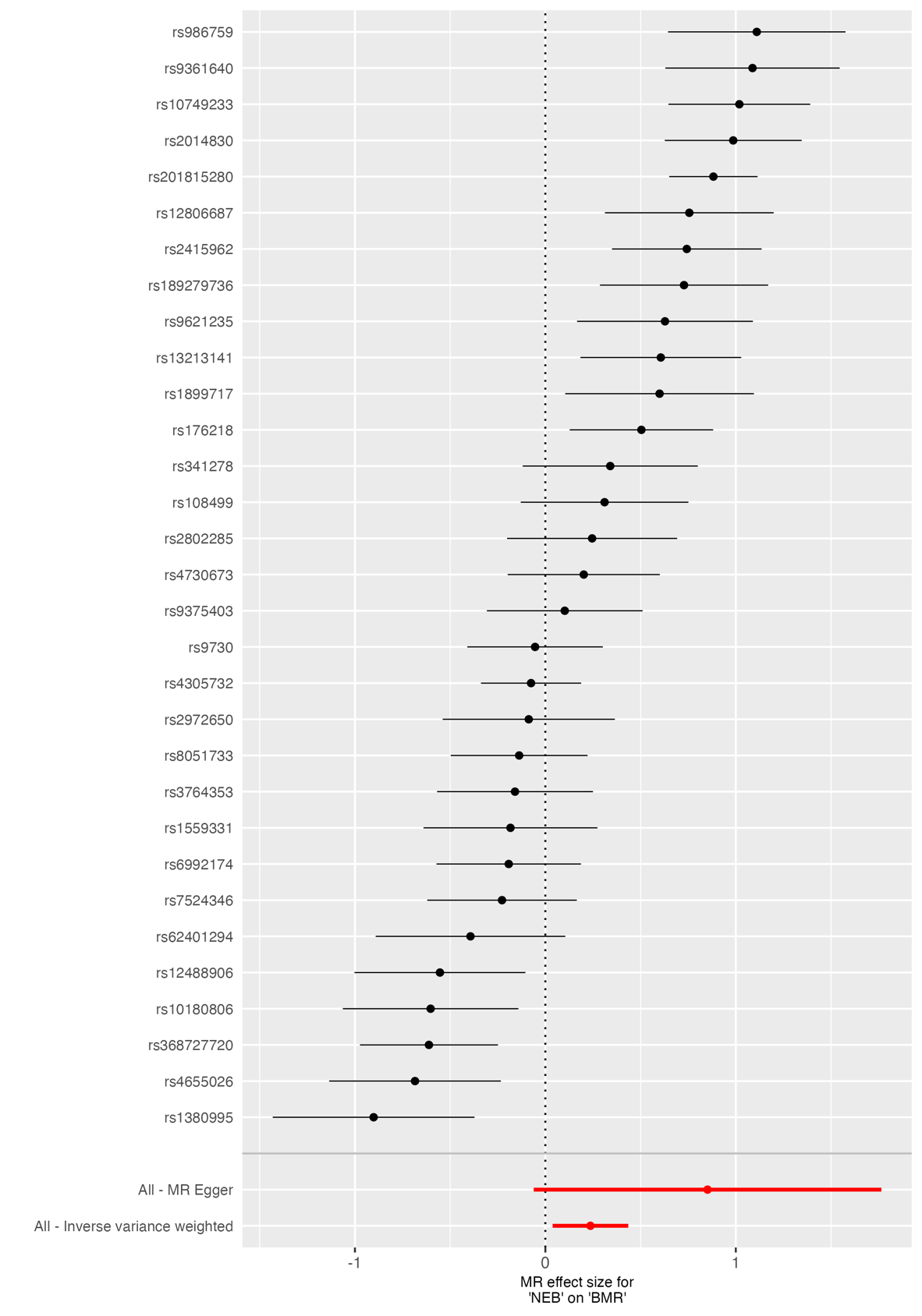

**Supplementary Figure 23.** *Forest plot of SNP-specific causal estimates for the effect of NEB on basal metabolic rate (BMR).* Forest plot displaying individual Wald ratio estimates and 95% confidence intervals for each SNP used as an instrumental variable for parity in the two-sample MR analysis. Summary causal estimates derived from both inverse-variance weighted (IVW) and MR Egger methods are shown at the bottom of the plot. Variation in SNP-specific effects reflects heterogeneity across instruments. NEB; Number of children ever born.

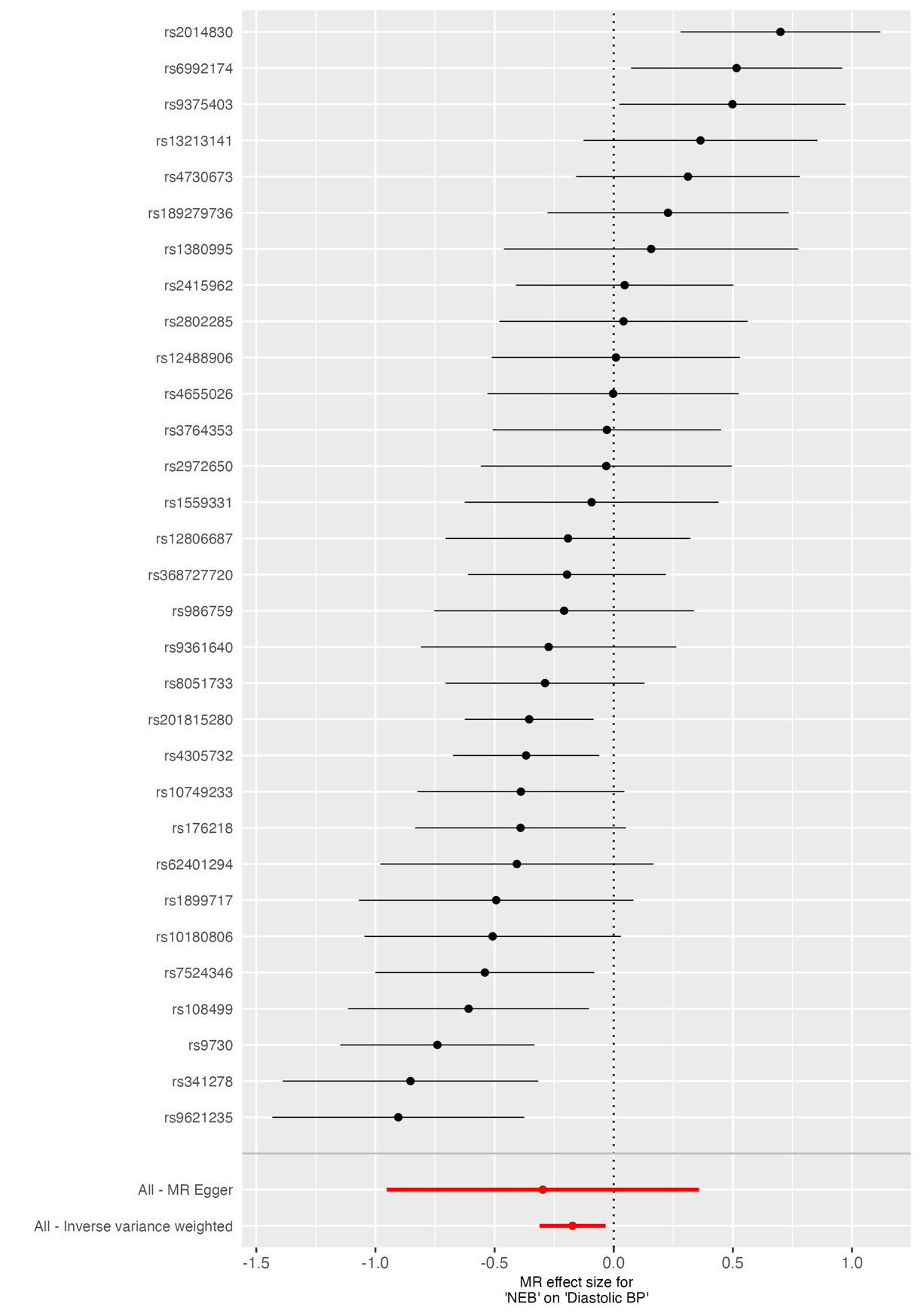

**Supplementary Figure 24.** *Forest plot of SNP-specific causal estimates for the effect of NEB on diastolic blood pressure (BP).* Forest plot displaying individual Wald ratio estimates and 95% confidence intervals for each SNP used as an instrumental variable for parity in the two-sample MR analysis. Summary causal estimates derived from both inverse-variance weighted (IVW) and MR Egger methods are shown at the bottom of the plot. Variation in SNP-specific effects reflects heterogeneity across instruments. NEB; Number of children ever born.

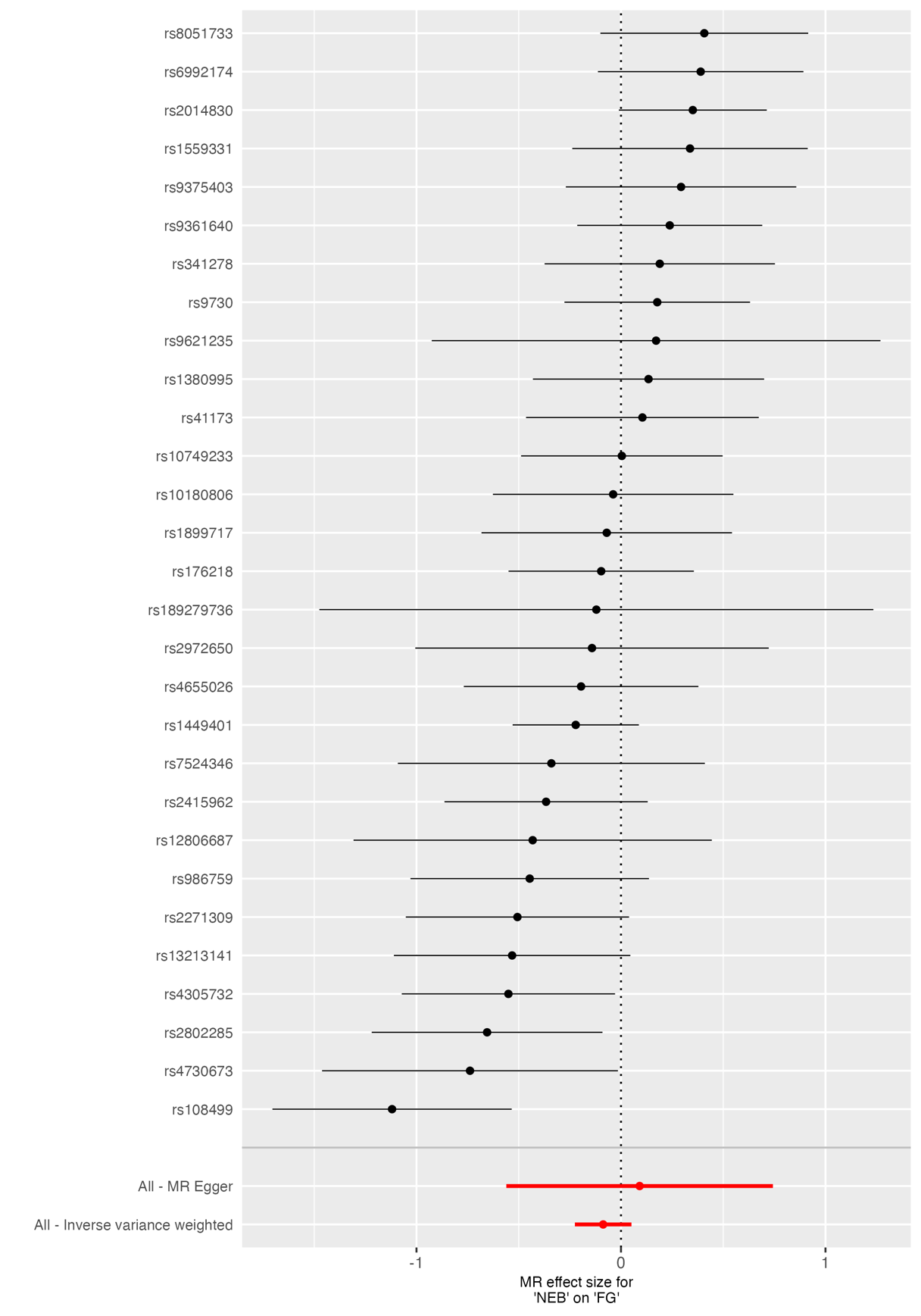

**Supplementary Figure 25.** *Forest plot of SNP-specific causal estimates for the effect of NEB on fasting glucose (FG).* Forest plot displaying individual Wald ratio estimates and 95% confidence intervals for each SNP used as an instrumental variable for parity in the two-sample MR analysis. Summary causal estimates derived from both inverse-variance weighted (IVW) and MR Egger methods are shown at the bottom of the plot. Variation in SNP-specific effects reflects heterogeneity across instruments. NEB; Number of children ever born.

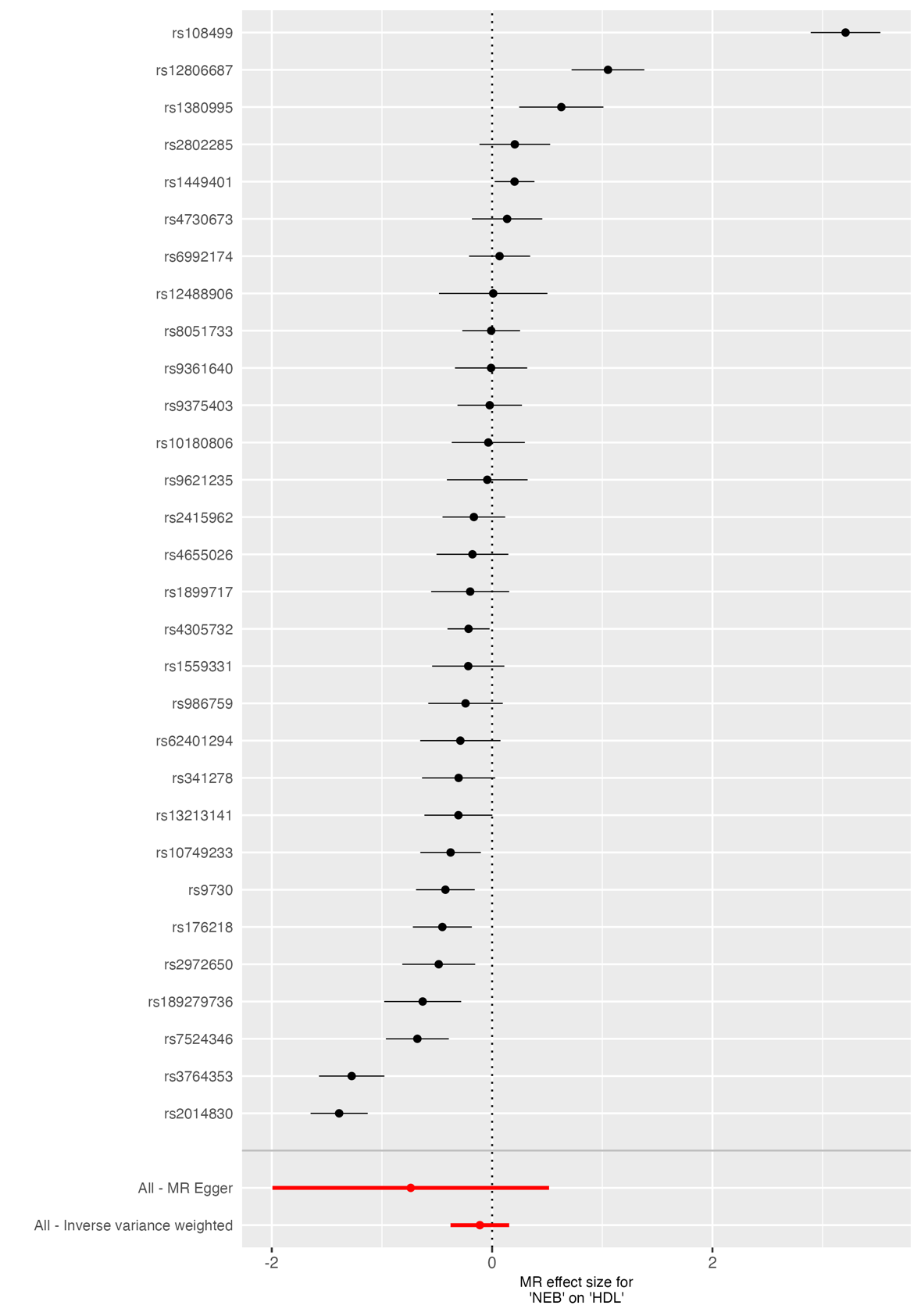

**Supplementary Figure 26.** *Forest plot of SNP-specific causal estimates for the effect of NEB on high-density lipoprotein (HDL).* Forest plot displaying individual Wald ratio estimates and 95% confidence intervals for each SNP used as an instrumental variable for parity in the two-sample MR analysis. Summary causal estimates derived from both inverse-variance weighted (IVW) and MR Egger methods are shown at the bottom of the plot. Variation in SNP-specific effects reflects heterogeneity across instruments. NEB; Number of children ever born.

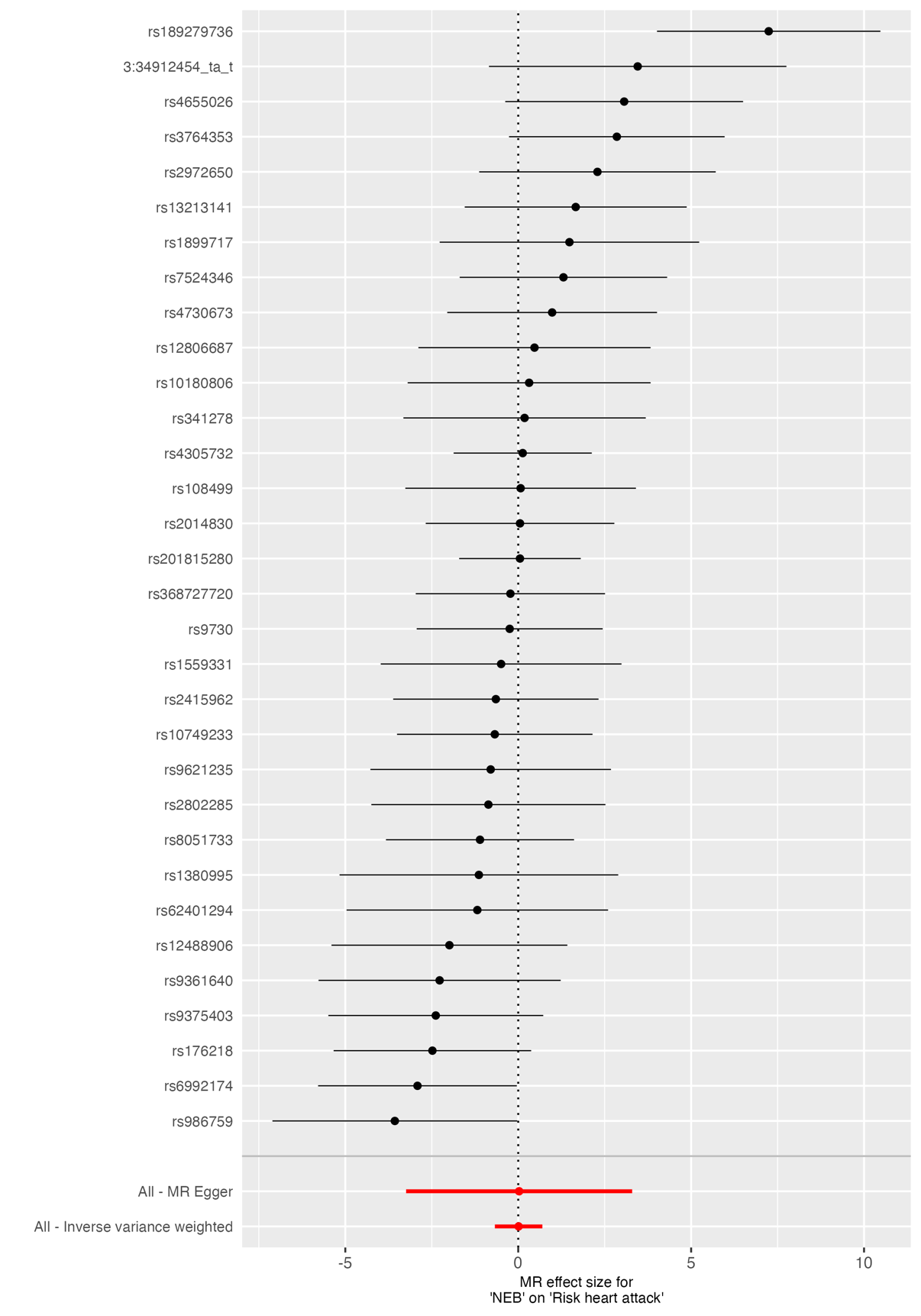

**Supplementary Figure 27.** *Forest plot of SNP-specific causal estimates for the effect of NEB on risk of myocardial infarction.* Forest plot displaying individual Wald ratio estimates and 95% confidence intervals for each SNP used as an instrumental variable for parity in the two-sample MR analysis. Summary causal estimates derived from both inverse-variance weighted (IVW) and MR Egger methods are shown at the bottom of the plot. Variation in SNP-specific effects reflects heterogeneity across instruments. NEB; Number of children ever born.

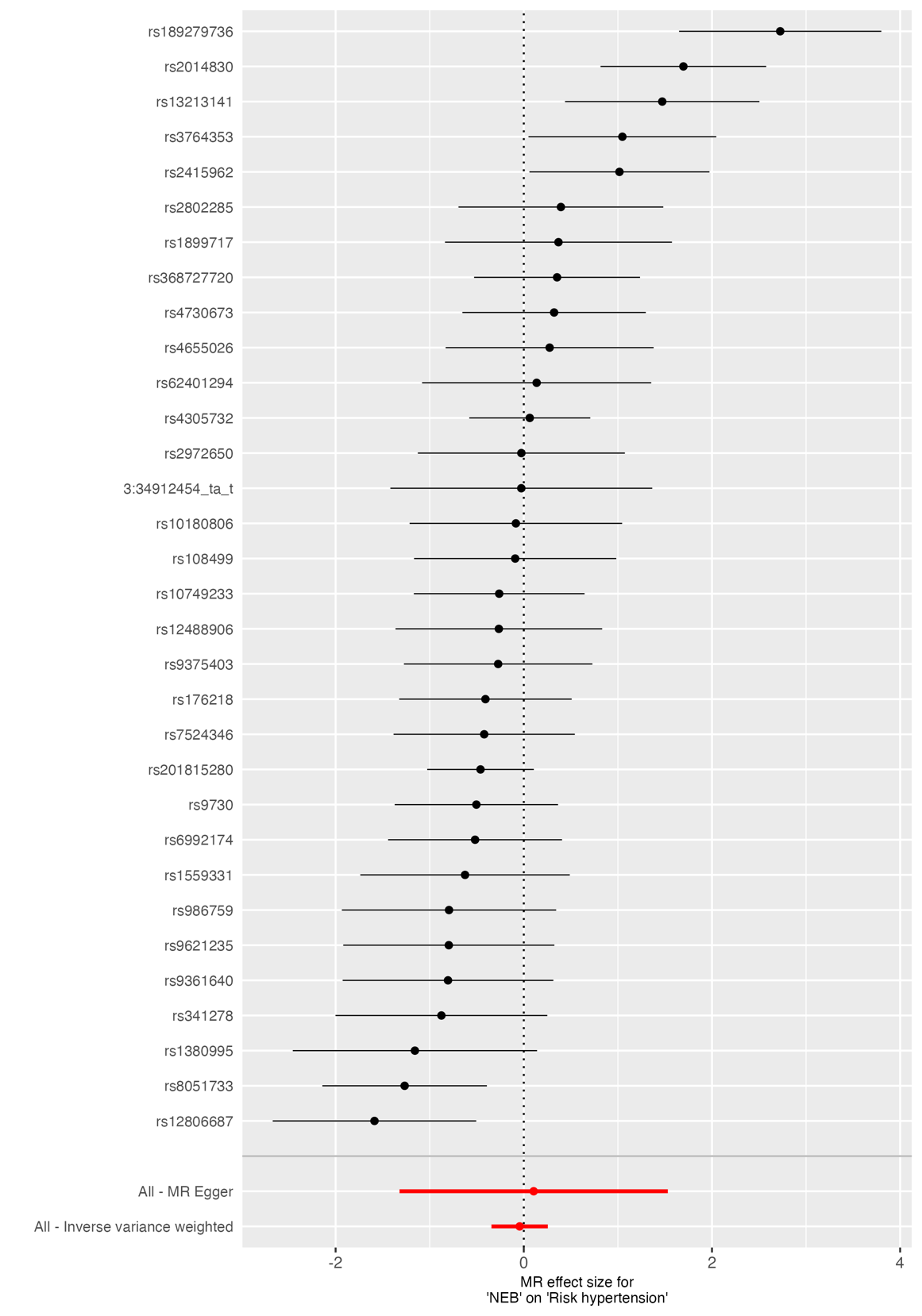

**Supplementary Figure 28.** *Forest plot of SNP-specific causal estimates for the effect of NEB on risk of hypertension.* Forest plot displaying individual Wald ratio estimates and 95% confidence intervals for each SNP used as an instrumental variable for parity in the two-sample MR analysis. Summary causal estimates derived from both inverse-variance weighted (IVW) and MR Egger methods are shown at the bottom of the plot. Variation in SNP-specific effects reflects heterogeneity across instruments. NEB; Number of children ever born.

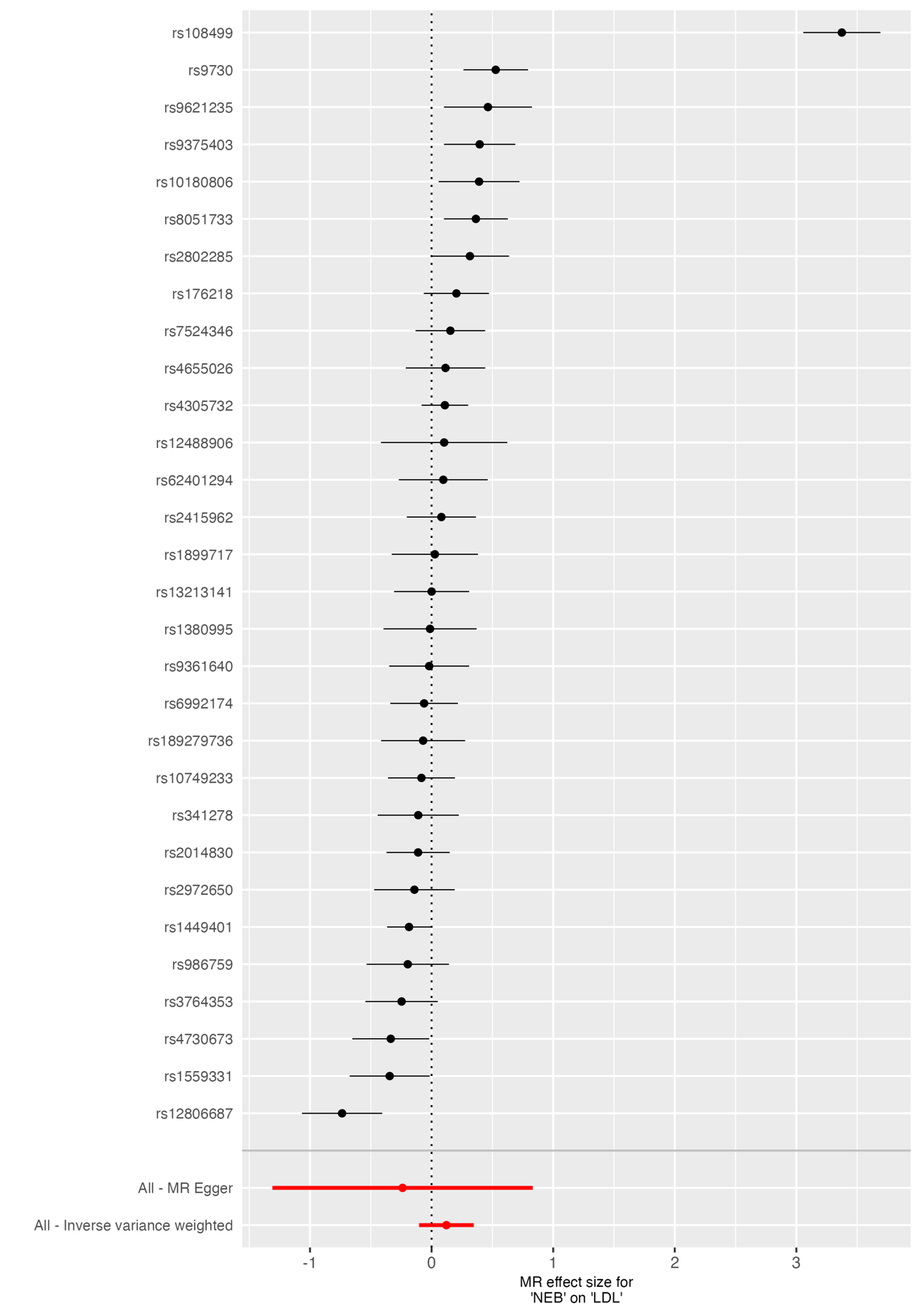

**Supplementary Figure 29.** *Forest plot of SNP-specific causal estimates for the effect of NEB on low-density lipoprotein (LDL) cholesterol.* Forest plot displaying individual Wald ratio estimates and 95% confidence intervals for each SNP used as an instrumental variable for parity in the two-sample MR analysis. Summary causal estimates derived from both inverse-variance weighted (IVW) and MR Egger methods are shown at the bottom of the plot. Variation in SNP-specific effects reflects heterogeneity across instruments. NEB; Number of children ever born.

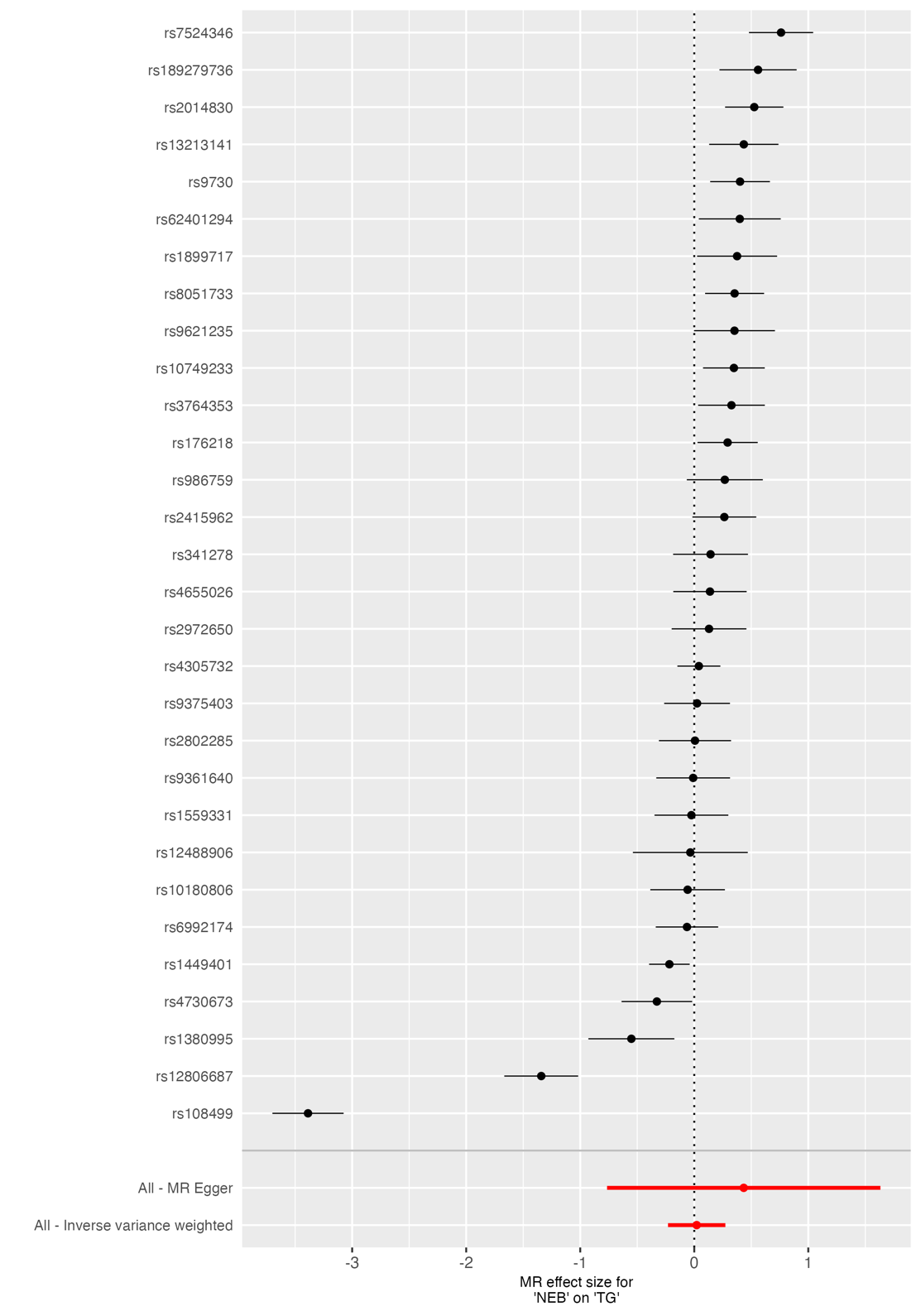

**Supplementary Figure 30.** *Forest plot of SNP-specific causal estimates for the effect of NEB on triglyceride (TG) level.* Forest plot displaying individual Wald ratio estimates and 95% confidence intervals for each SNP used as an instrumental variable for parity in the two-sample MR analysis. Summary causal estimates derived from both inverse-variance weighted (IVW) and MR Egger methods are shown at the bottom of the plot. Variation in SNP-specific effects reflects heterogeneity across instruments. TG in outcome GWAS was log-transformed. NEB; Number of children ever born.

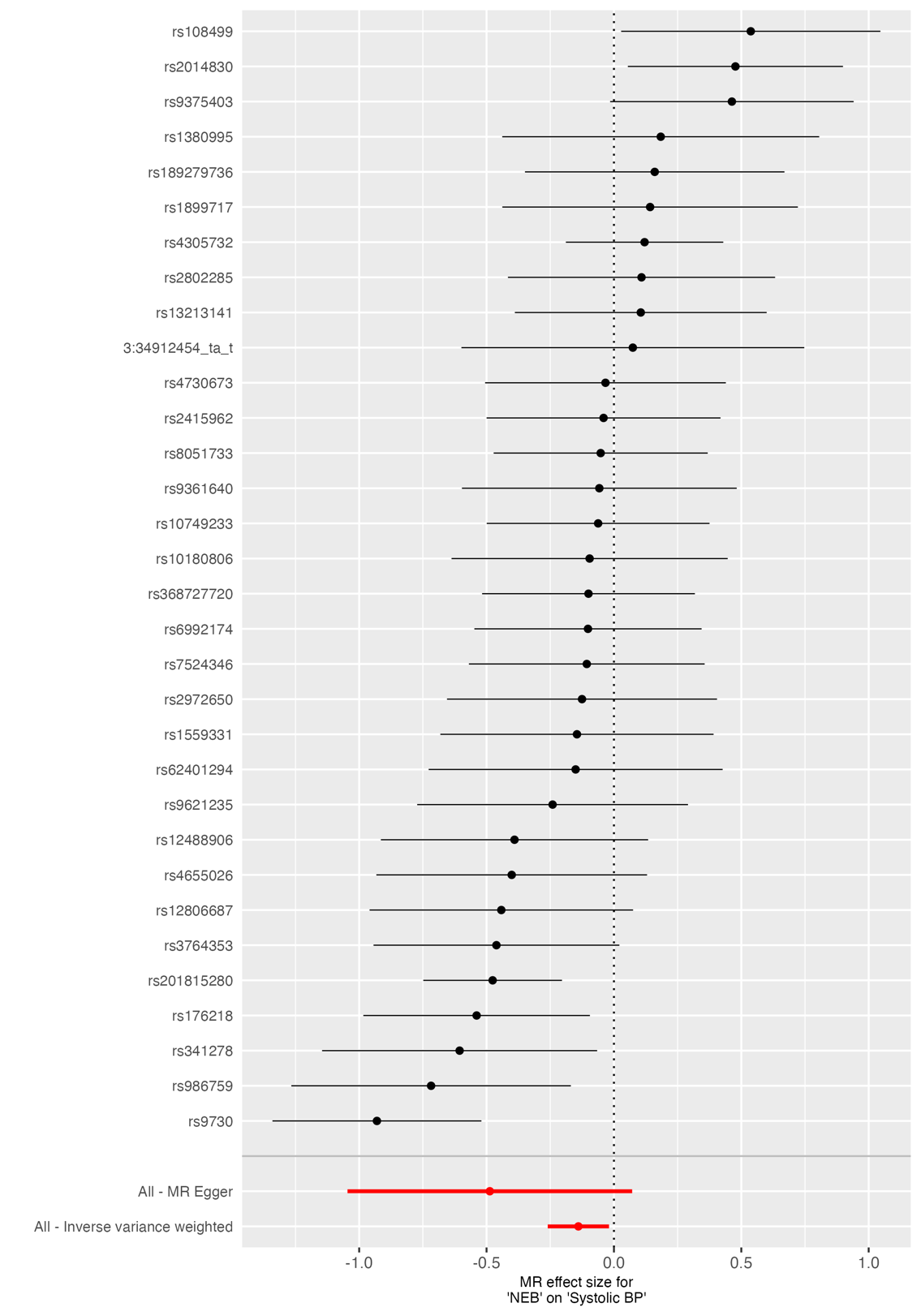

**Supplementary Figure 31.** *Forest plot of SNP-specific causal estimates for the effect of NEB on systolic blood pressure (BP).* Forest plot displaying individual Wald ratio estimates and 95% confidence intervals for each SNP used as an instrumental variable for parity in the two-sample MR analysis. Summary causal estimates derived from both inverse-variance weighted (IVW) and MR Egger methods are shown at the bottom of the plot. Variation in SNP-specific effects reflects heterogeneity across instruments. TG in outcome GWAS was log-transformed. NEB; Number of children ever born.

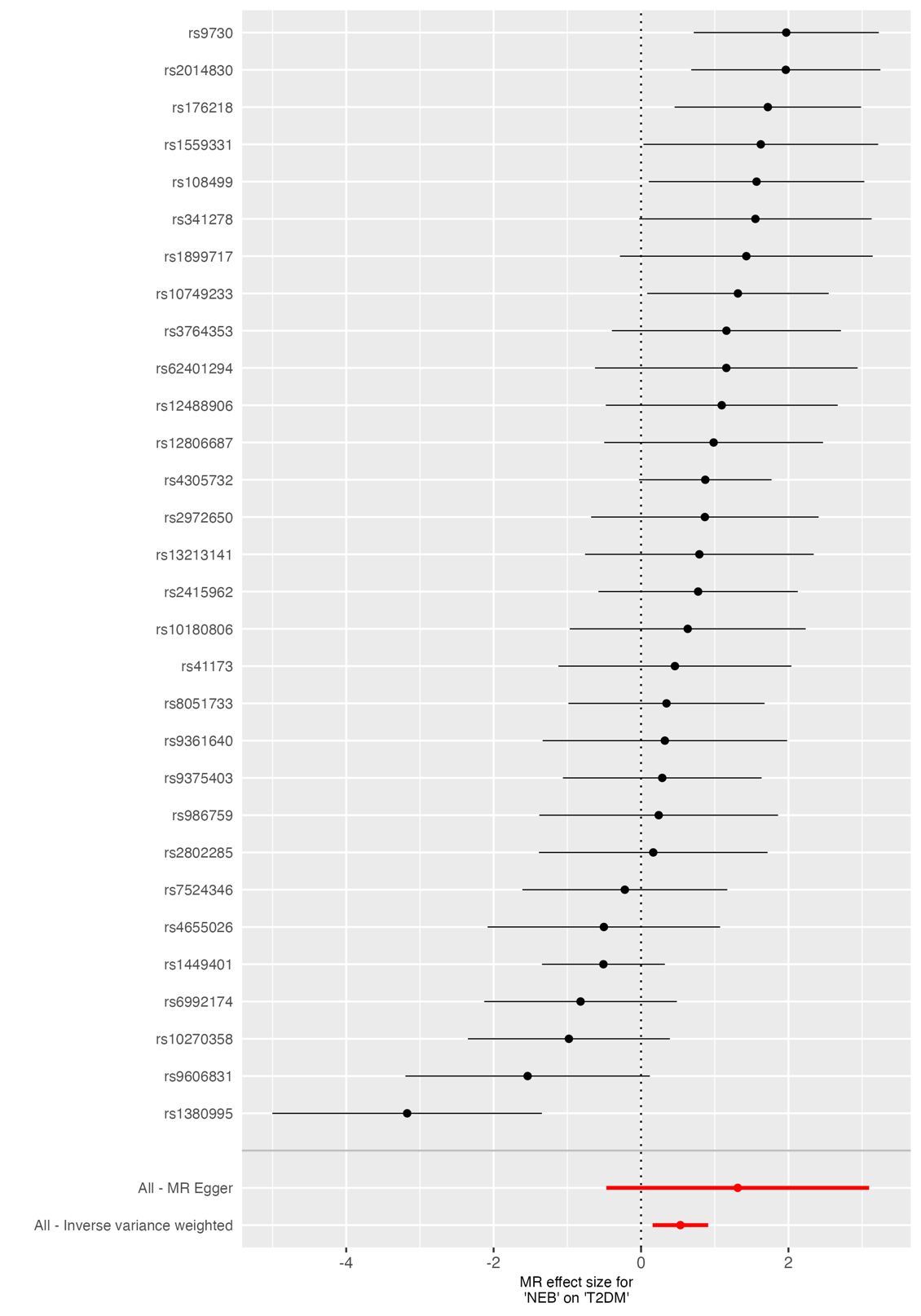

**Supplementary Figure 32.** *Forest plot of SNP-specific causal estimates for the effect of NEB on type-II diabetes (T2DM).* Forest plot displaying individual Wald ratio estimates and 95% confidence intervals for each SNP used as an instrumental variable for parity in the two-sample MR analysis. Summary causal estimates derived from both inverse-variance weighted (IVW) and MR Egger methods are shown at the bottom of the plot. Variation in SNP-specific effects reflects heterogeneity across instruments. TG in outcome GWAS was log-transformed. NEB; Number of children ever born.

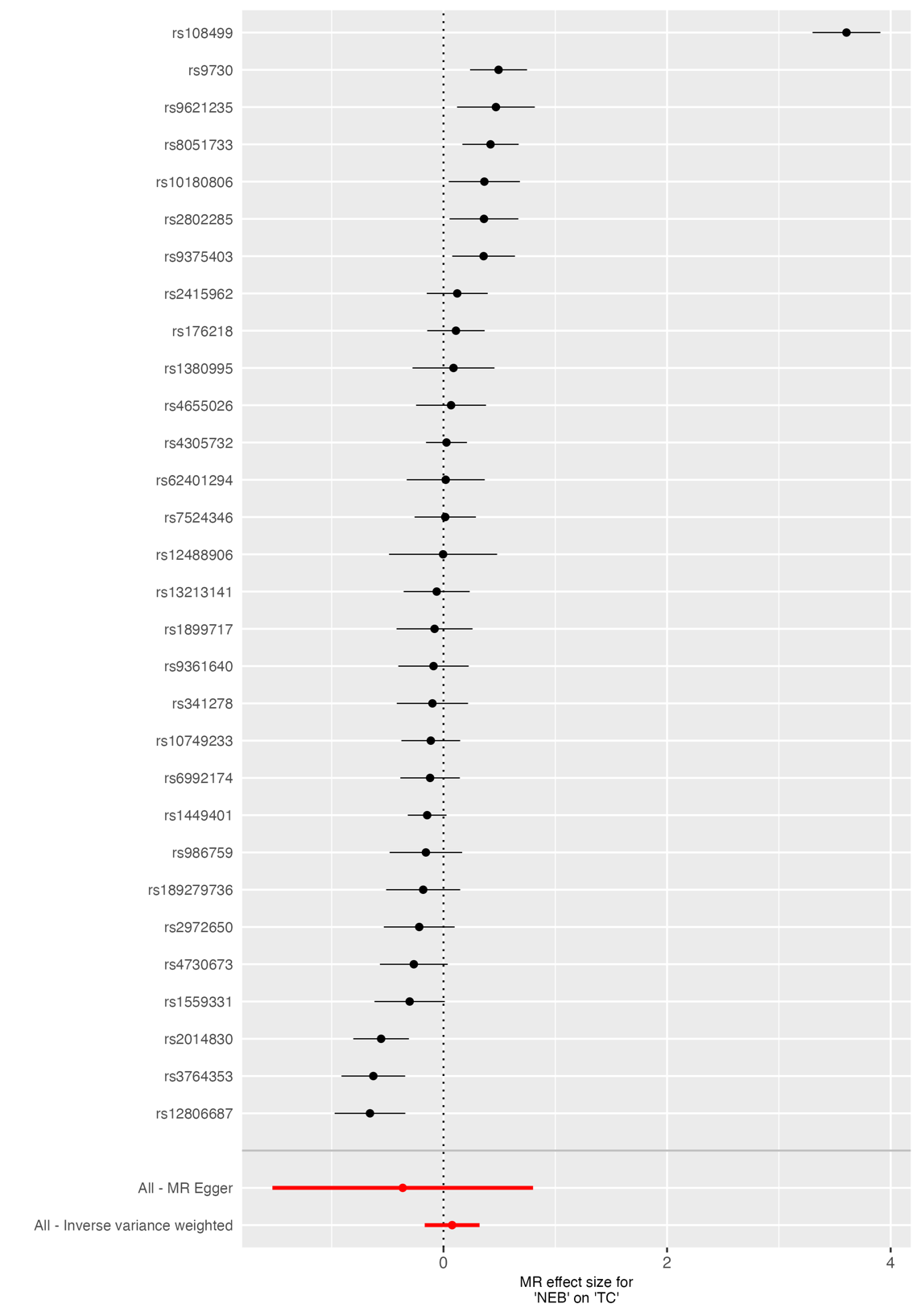

**Supplementary Figure 33.** *Forest plot of SNP-specific causal estimates for the effect of NEB on total cholesterol (TC).* Forest plot displaying individual Wald ratio estimates and 95% confidence intervals for each SNP used as an instrumental variable for parity in the two-sample MR analysis. Summary causal estimates derived from both inverse-variance weighted (IVW) and MR Egger methods are shown at the bottom of the plot. Variation in SNP-specific effects reflects heterogeneity across instruments. TG in outcome GWAS was log-transformed. NEB; Number of children ever born.

#### 7.2 Male-specific analyses

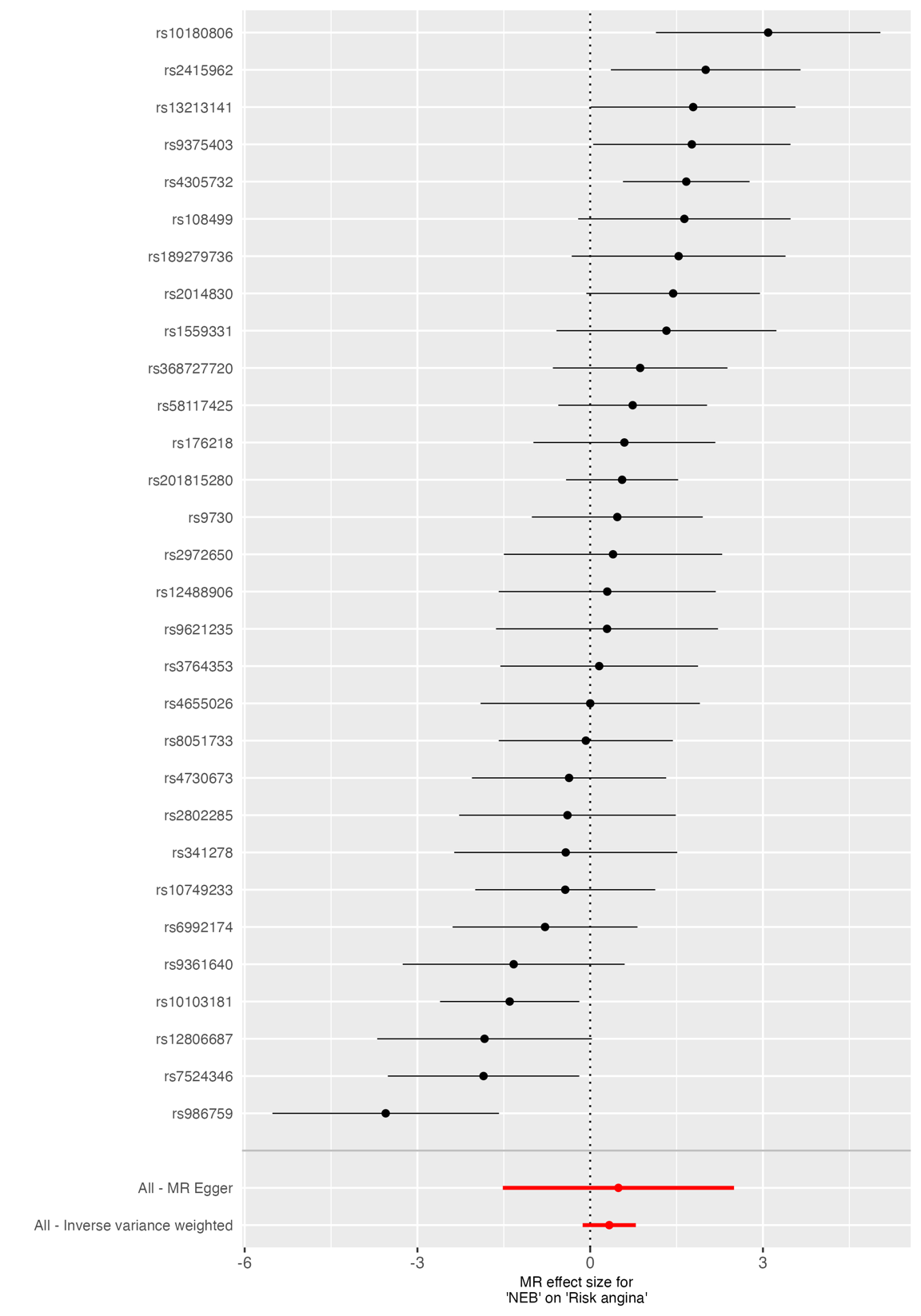

**Supplementary Figure 34.** *Forest plot of SNP-specific causal estimates for the effect of NEB on angina.* Forest plot displaying individual Wald ratio estimates and 95% confidence intervals for each SNP used as an instrumental variable for parity in the two-sample MR analysis. Summary causal estimates derived from both inverse-variance weighted (IVW) and MR Egger methods are shown at the bottom of the plot. Variation in SNP-specific effects reflects heterogeneity across instruments. NEB; Number of children ever born.

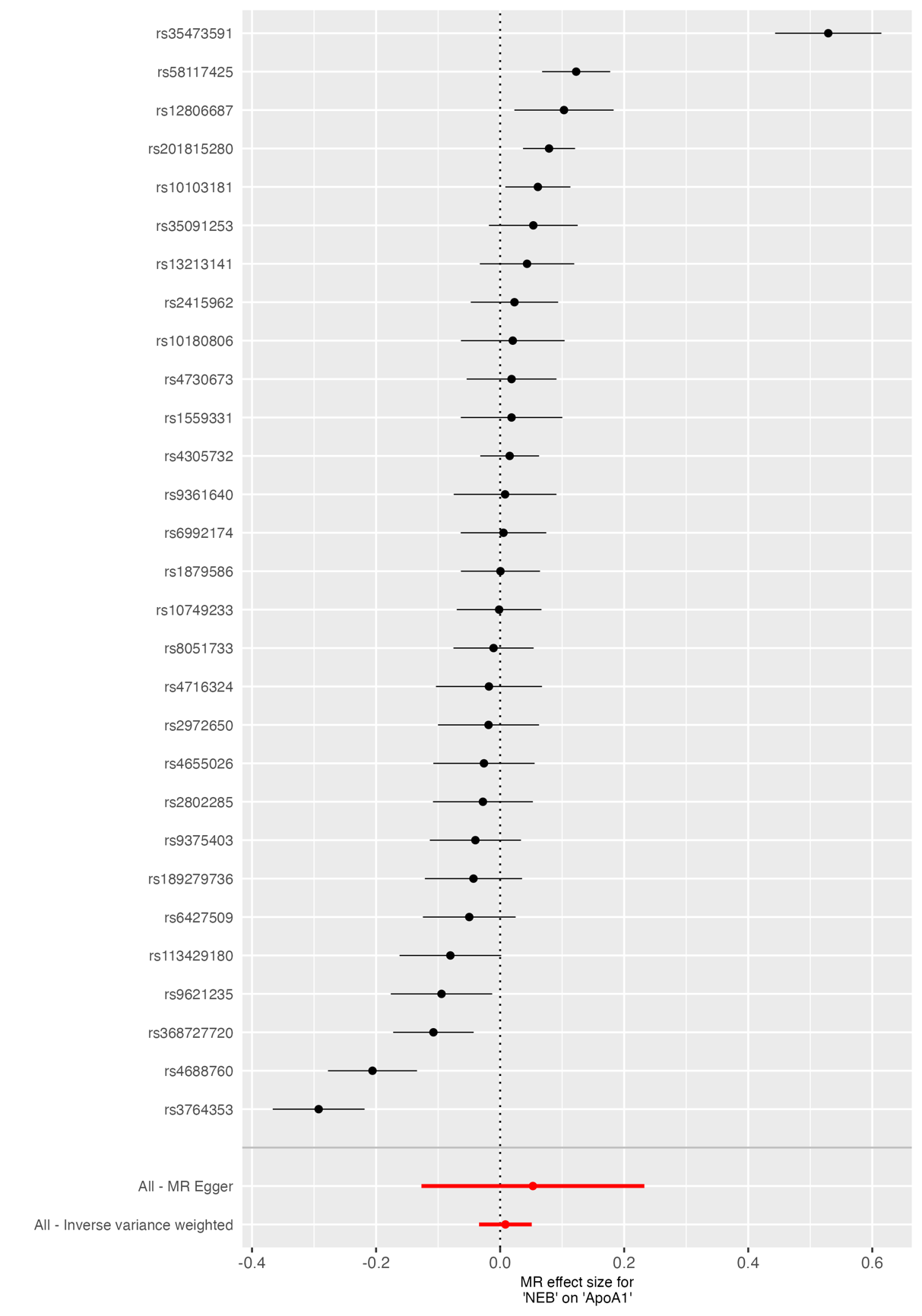

**Supplementary Figure 35.** *Forest plot of SNP-specific causal estimates for the effect of NEB on ApoA1.* Forest plot displaying individual Wald ratio estimates and 95% confidence intervals for each SNP used as an instrumental variable for parity in the two-sample MR analysis. Summary causal estimates derived from both inverse-variance weighted (IVW) and MR Egger methods are shown at the bottom of the plot. Variation in SNP-specific effects reflects heterogeneity across instruments. NEB; Number of children ever born.

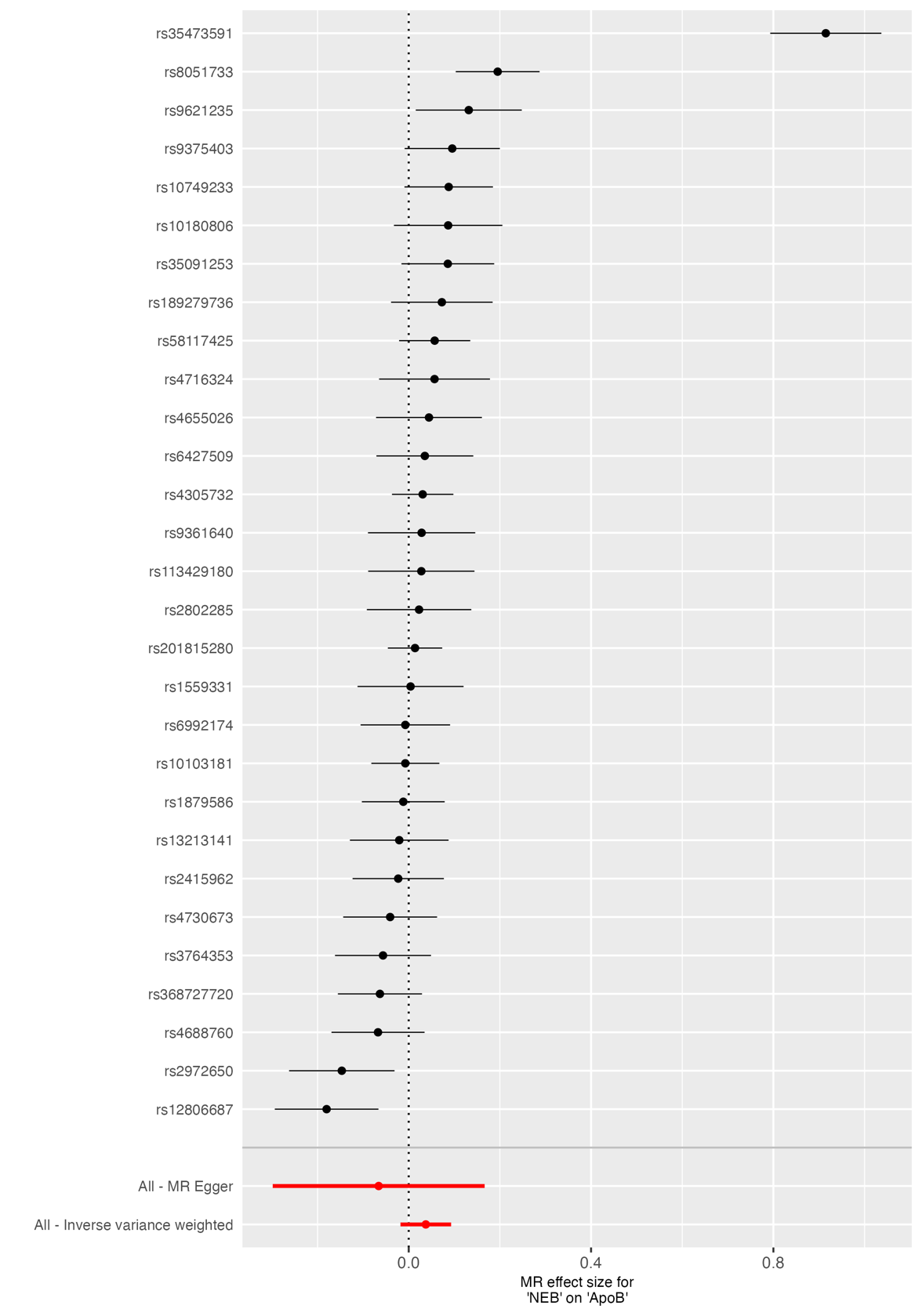

**Supplementary Figure 36.** *Forest plot of SNP-specific causal estimates for the effect of NEB on ApoB.* Forest plot displaying individual Wald ratio estimates and 95% confidence intervals for each SNP used as an instrumental variable for parity in the two-sample MR analysis. Summary causal estimates derived from both inverse-variance weighted (IVW) and MR Egger methods are shown at the bottom of the plot. Variation in SNP-specific effects reflects heterogeneity across instruments. NEB; Number of children ever born.

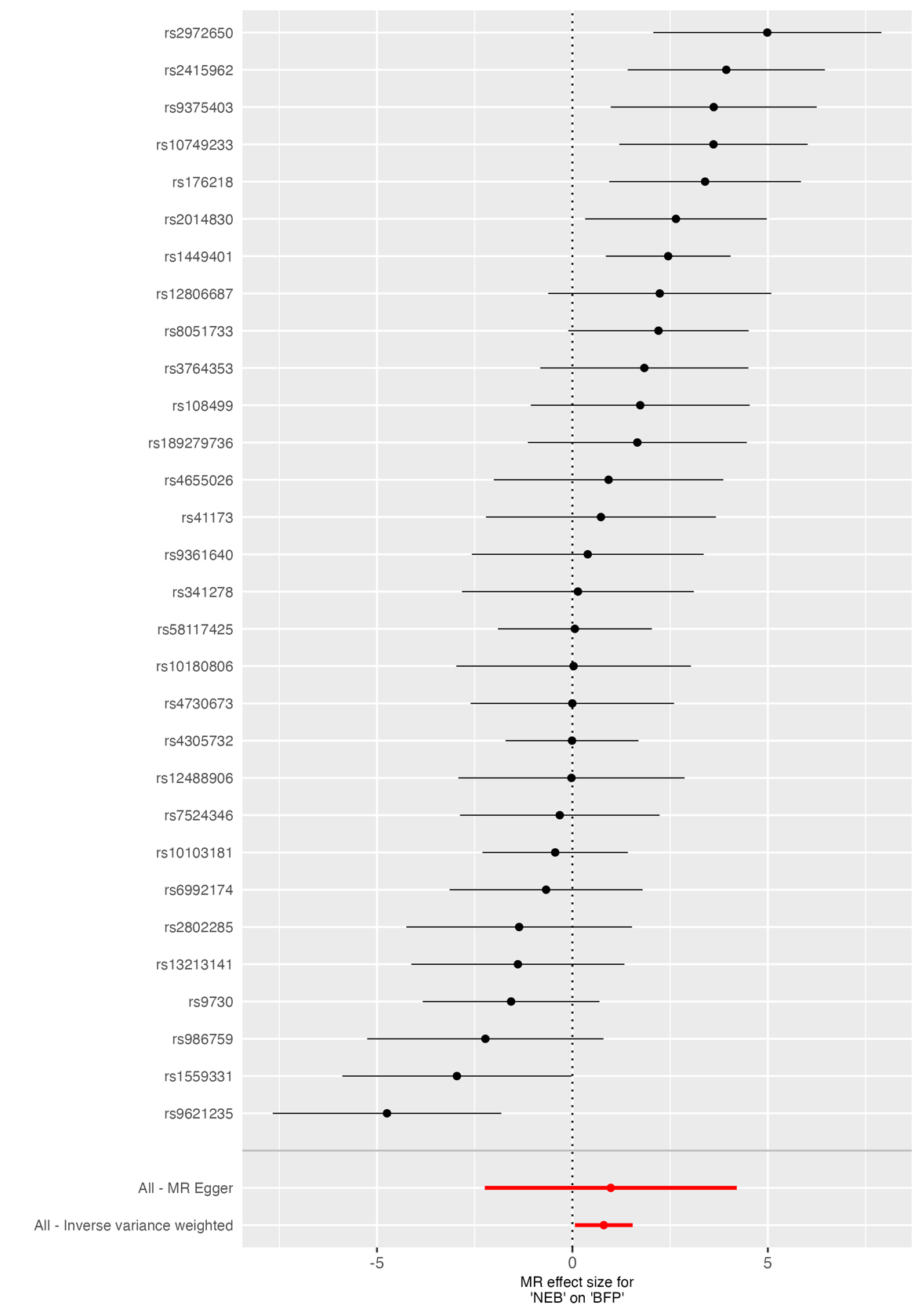

**Supplementary Figure 37.** *Forest plot of SNP-specific causal estimates for the effect of NEB on body fat percentage (BFP).* Forest plot displaying individual Wald ratio estimates and 95% confidence intervals for each SNP used as an instrumental variable for parity in the two-sample MR analysis. Summary causal estimates derived from both inverse-variance weighted (IVW) and MR Egger methods are shown at the bottom of the plot. Variation in SNP-specific effects reflects heterogeneity across instruments. NEB; Number of children ever born.

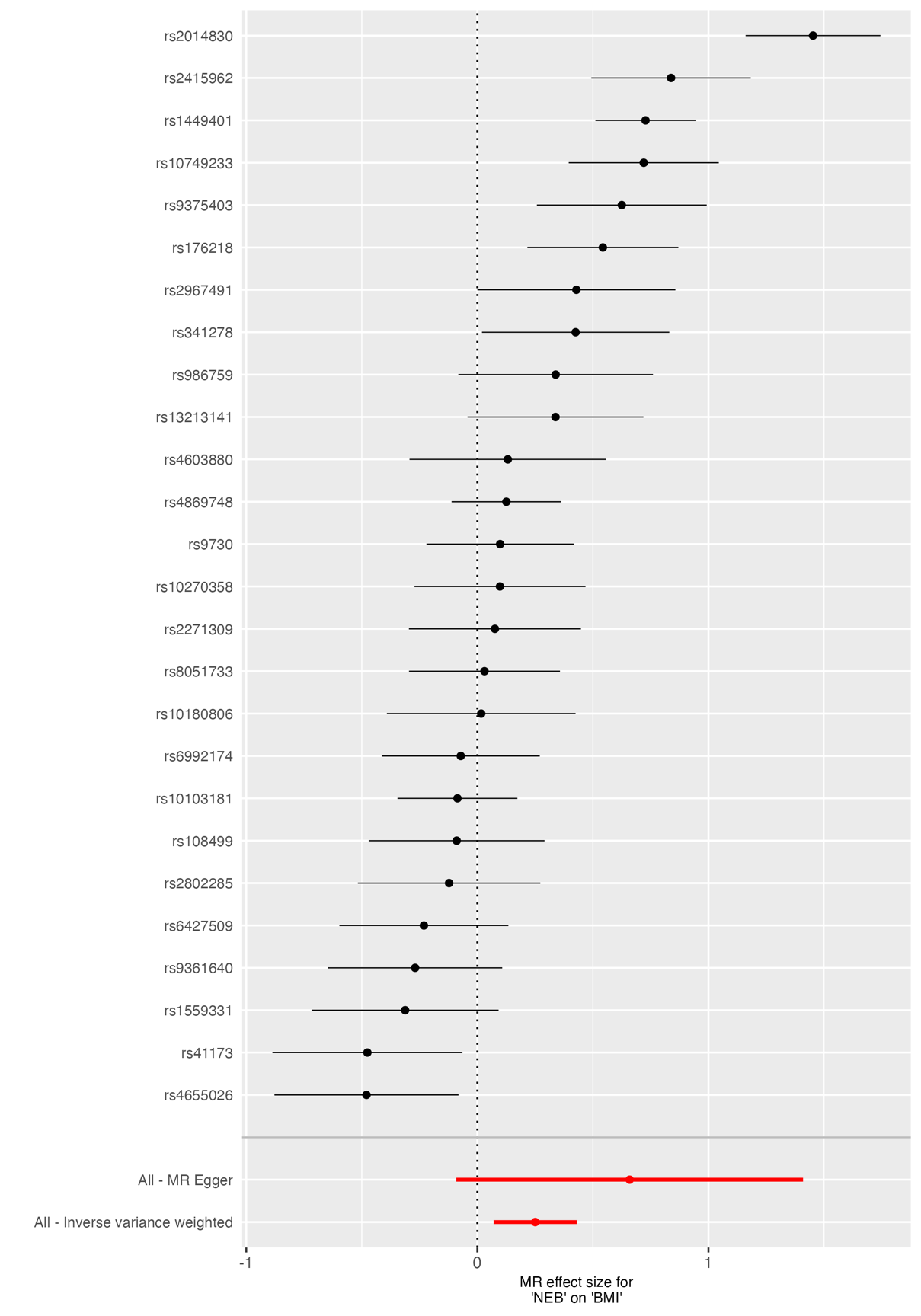

**Supplementary Figure 38.** *Forest plot of SNP-specific causal estimates for the effect of NEB on body mass index (BMI).* Forest plot displaying individual Wald ratio estimates and 95% confidence intervals for each SNP used as an instrumental variable for parity in the two-sample MR analysis. Summary causal estimates derived from both inverse-variance weighted (IVW) and MR Egger methods are shown at the bottom of the plot. Variation in SNP-specific effects reflects heterogeneity across instruments. NEB; Number of children ever born.

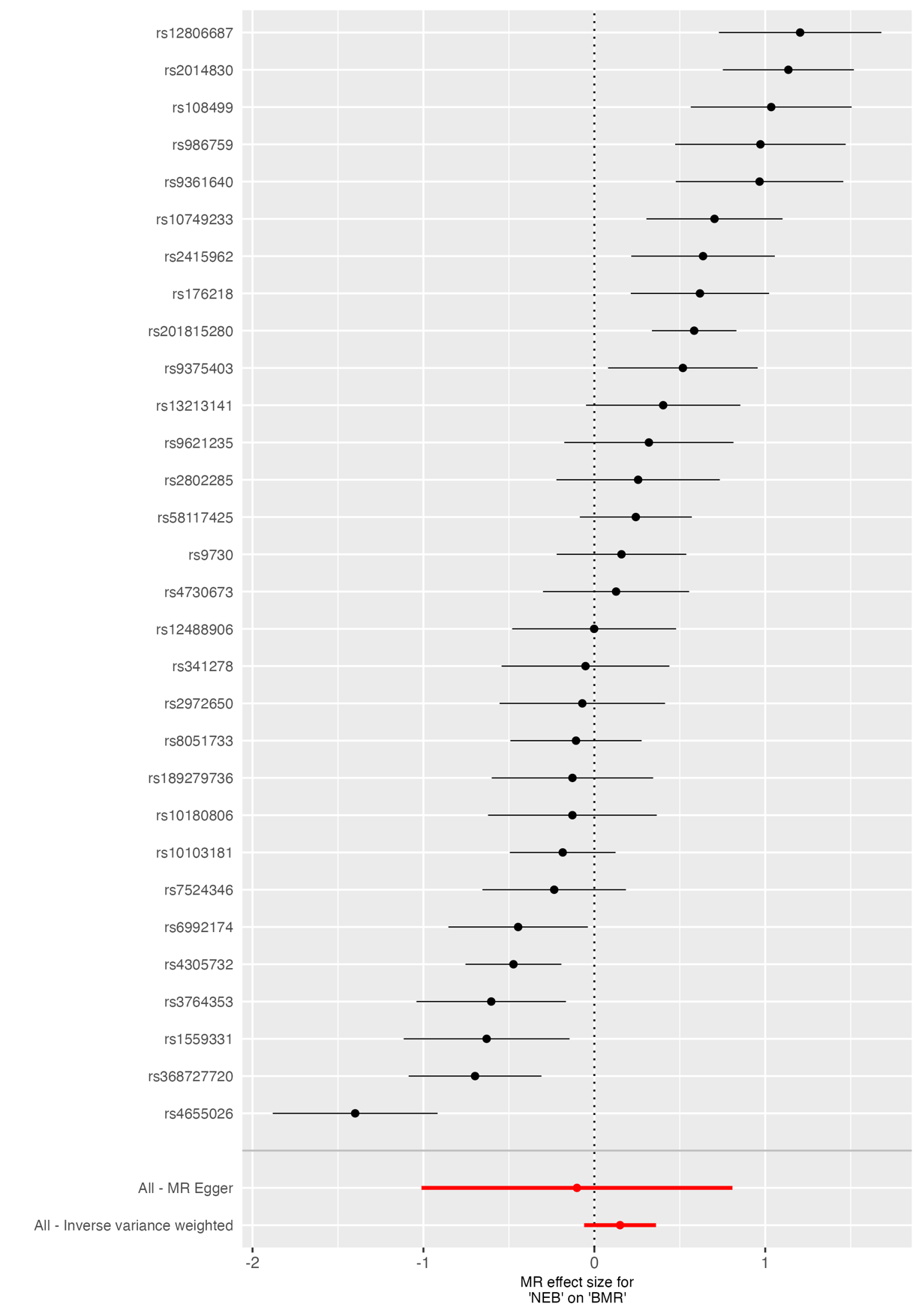

**Supplementary Figure 39.** *Forest plot of SNP-specific causal estimates for the effect of NEB on basal metabolic rate (BMR).* Forest plot displaying individual Wald ratio estimates and 95% confidence intervals for each SNP used as an instrumental variable for parity in the two-sample MR analysis. Summary causal estimates derived from both inverse-variance weighted (IVW) and MR Egger methods are shown at the bottom of the plot. Variation in SNP-specific effects reflects heterogeneity across instruments. NEB; Number of children ever born.

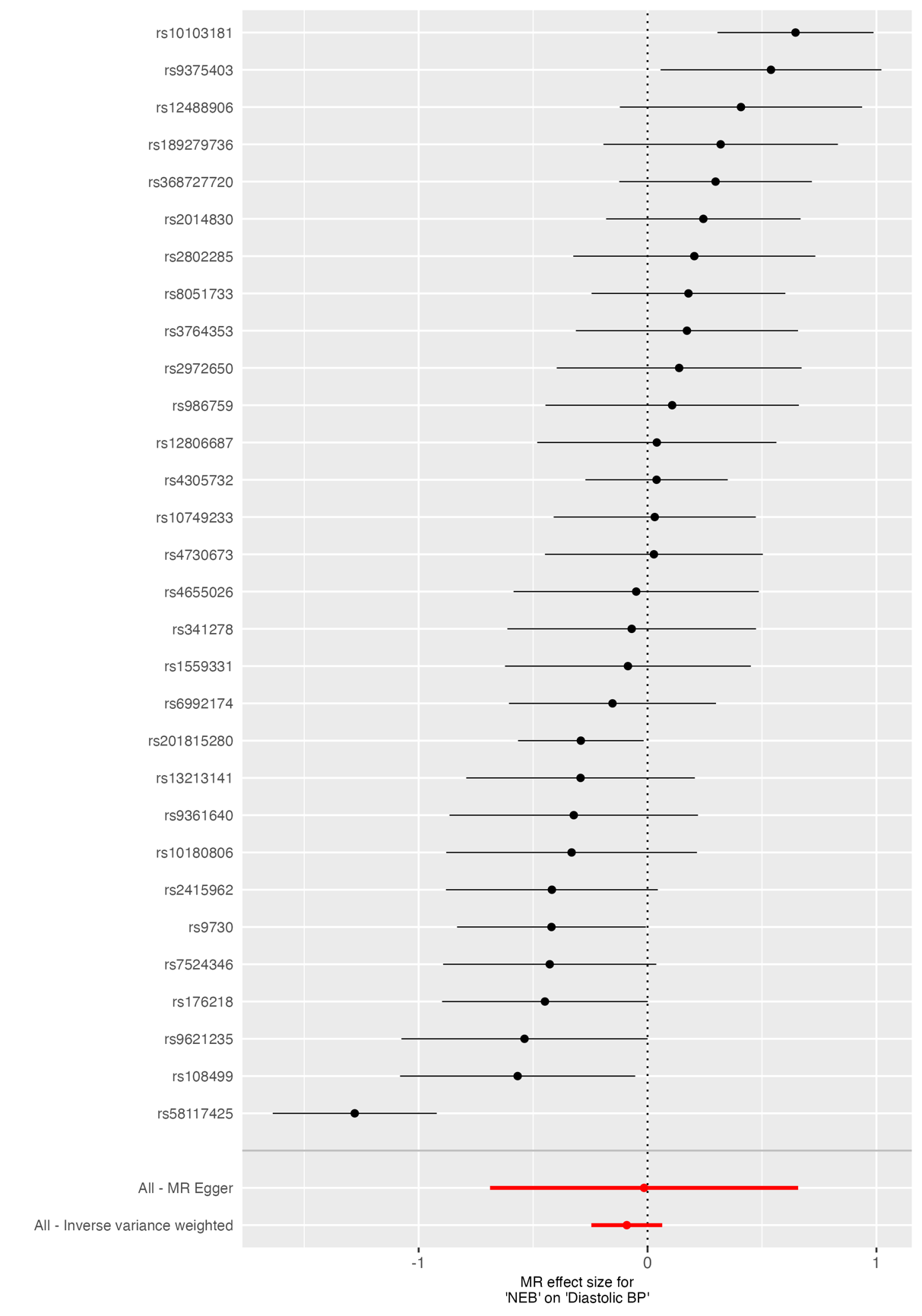

**Supplementary Figure 40.** *Forest plot of SNP-specific causal estimates for the effect of NEB on diastolic blood pressure (BP).* Forest plot displaying individual Wald ratio estimates and 95% confidence intervals for each SNP used as an instrumental variable for parity in the two-sample MR analysis. Summary causal estimates derived from both inverse-variance weighted (IVW) and MR Egger methods are shown at the bottom of the plot. Variation in SNP-specific effects reflects heterogeneity across instruments. NEB; Number of children ever born.

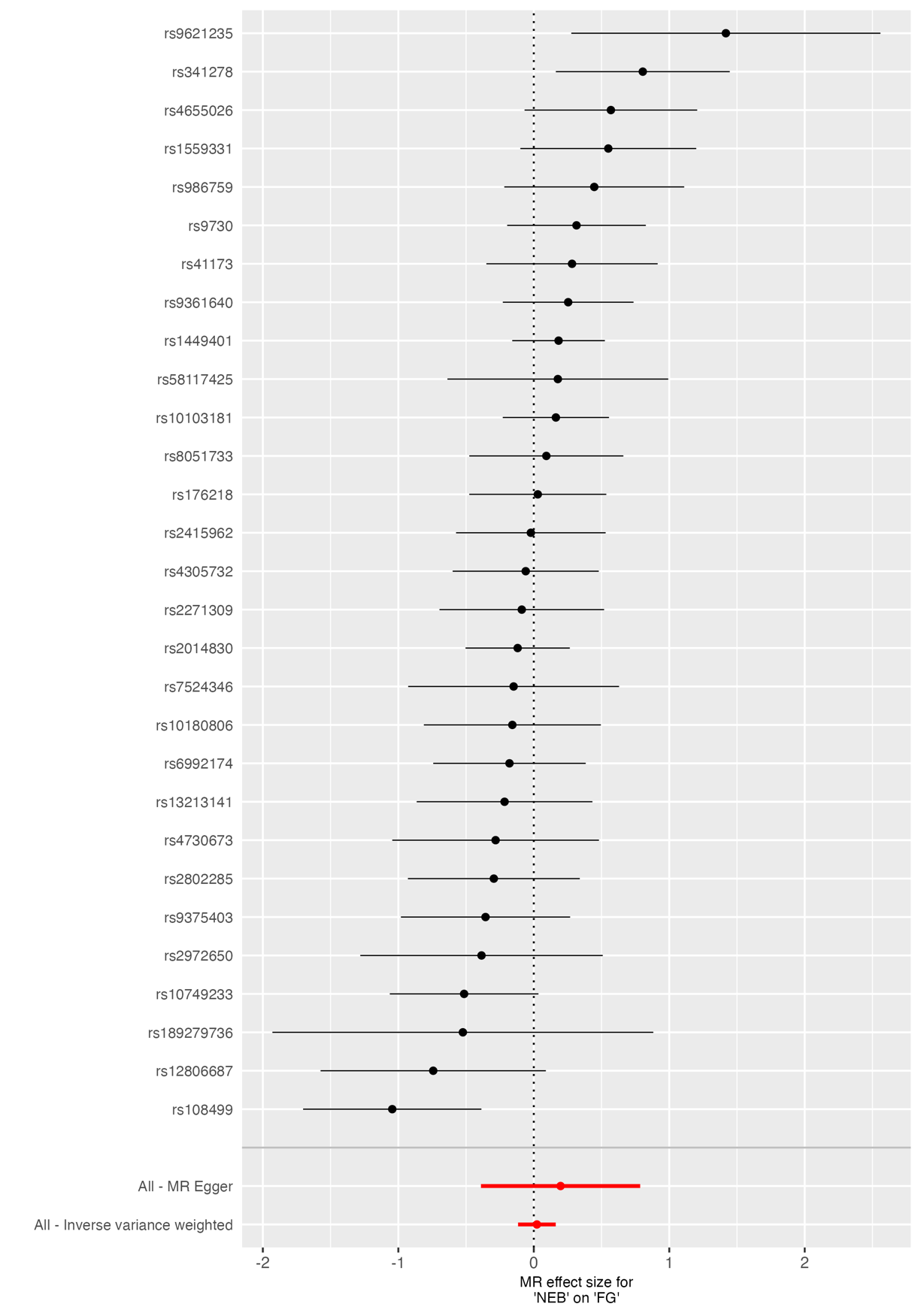

**Supplementary Figure 41.** *Forest plot of SNP-specific causal estimates for the effect of NEB on fasting glucose (FG).* Forest plot displaying individual Wald ratio estimates and 95% confidence intervals for each SNP used as an instrumental variable for parity in the two-sample MR analysis. Summary causal estimates derived from both inverse-variance weighted (IVW) and MR Egger methods are shown at the bottom of the plot. Variation in SNP-specific effects reflects heterogeneity across instruments. NEB; Number of children ever born.

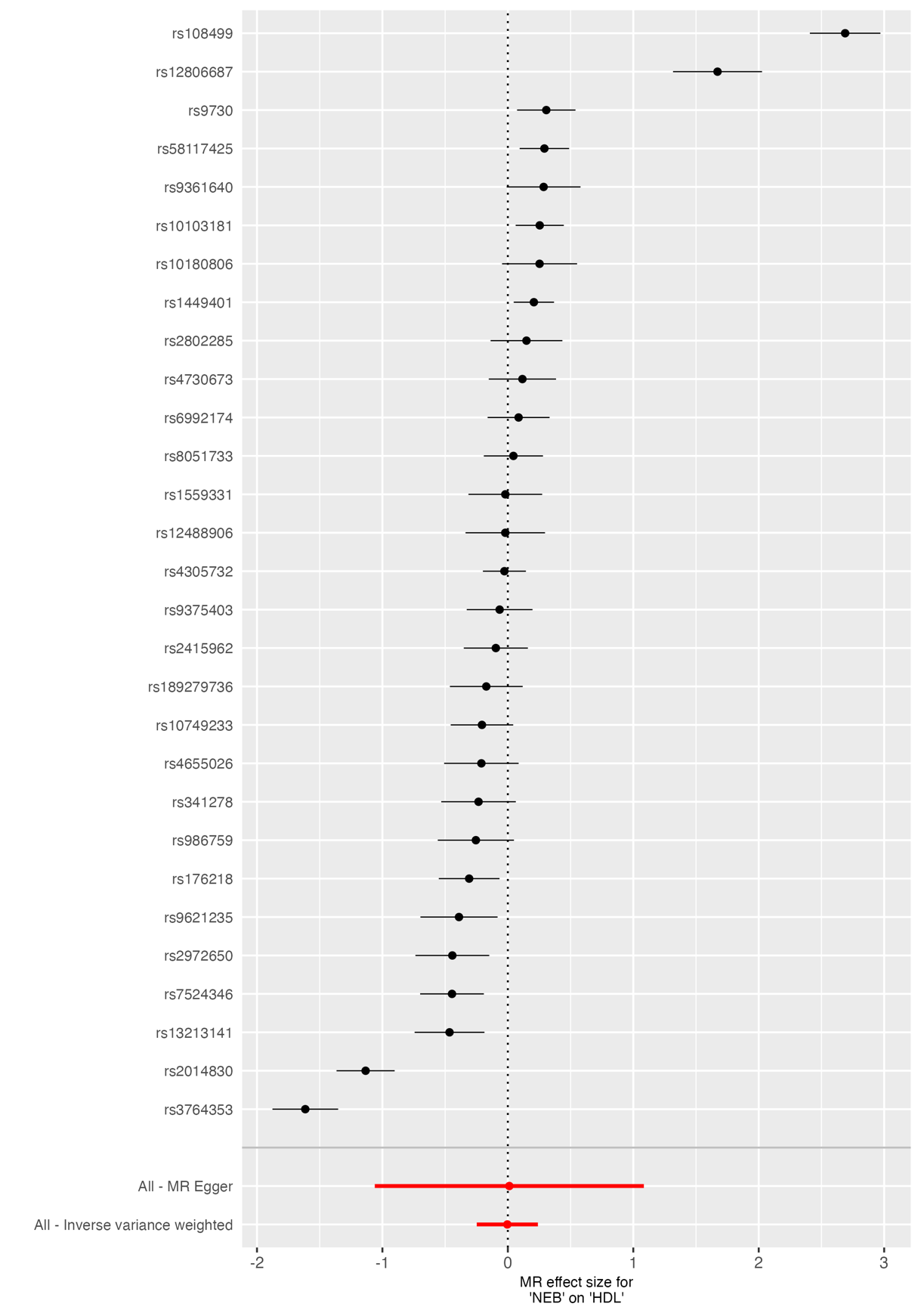

**Supplementary Figure 42.** *Forest plot of SNP-specific causal estimates for the effect of NEB on high-density lipoprotein (HDL).* Forest plot displaying individual Wald ratio estimates and 95% confidence intervals for each SNP used as an instrumental variable for parity in the two-sample MR analysis. Summary causal estimates derived from both inverse-variance weighted (IVW) and MR Egger methods are shown at the bottom of the plot. Variation in SNP-specific effects reflects heterogeneity across instruments. NEB; Number of children ever born.

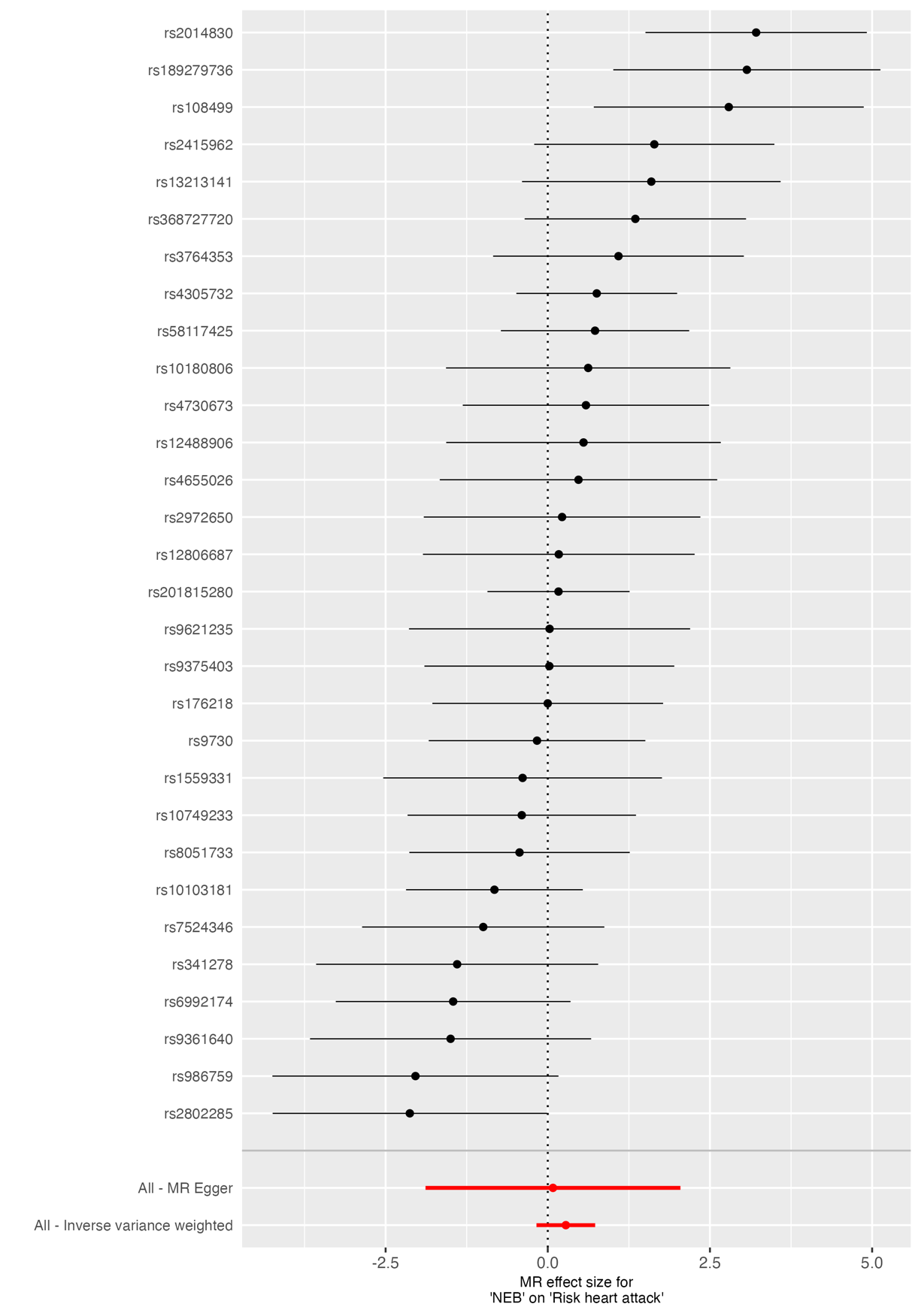

**Supplementary Figure 43.** *Forest plot of SNP-specific causal estimates for the effect of NEB on risk of myocardial infarction.* Forest plot displaying individual Wald ratio estimates and 95% confidence intervals for each SNP used as an instrumental variable for parity in the two-sample MR analysis. Summary causal estimates derived from both inverse-variance weighted (IVW) and MR Egger methods are shown at the bottom of the plot. Variation in SNP-specific effects reflects heterogeneity across instruments. NEB; Number of children ever born.

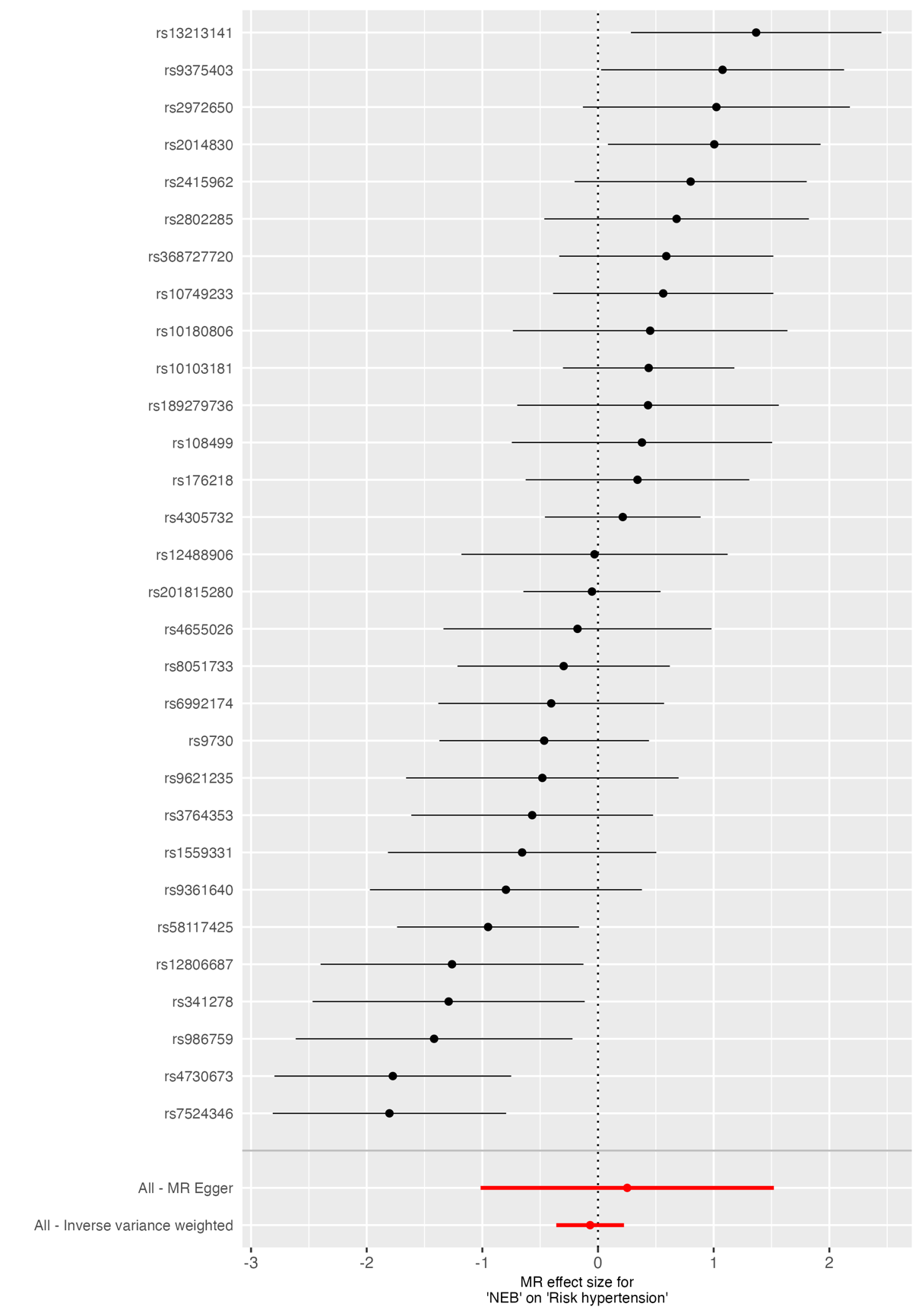

**Supplementary Figure 44.** *Forest plot of SNP-specific causal estimates for the effect of NEB on risk of hypertension.* Forest plot displaying individual Wald ratio estimates and 95% confidence intervals for each SNP used as an instrumental variable for parity in the two-sample MR analysis. Summary causal estimates derived from both inverse-variance weighted (IVW) and MR Egger methods are shown at the bottom of the plot. Variation in SNP-specific effects reflects heterogeneity across instruments. NEB; Number of children ever born.

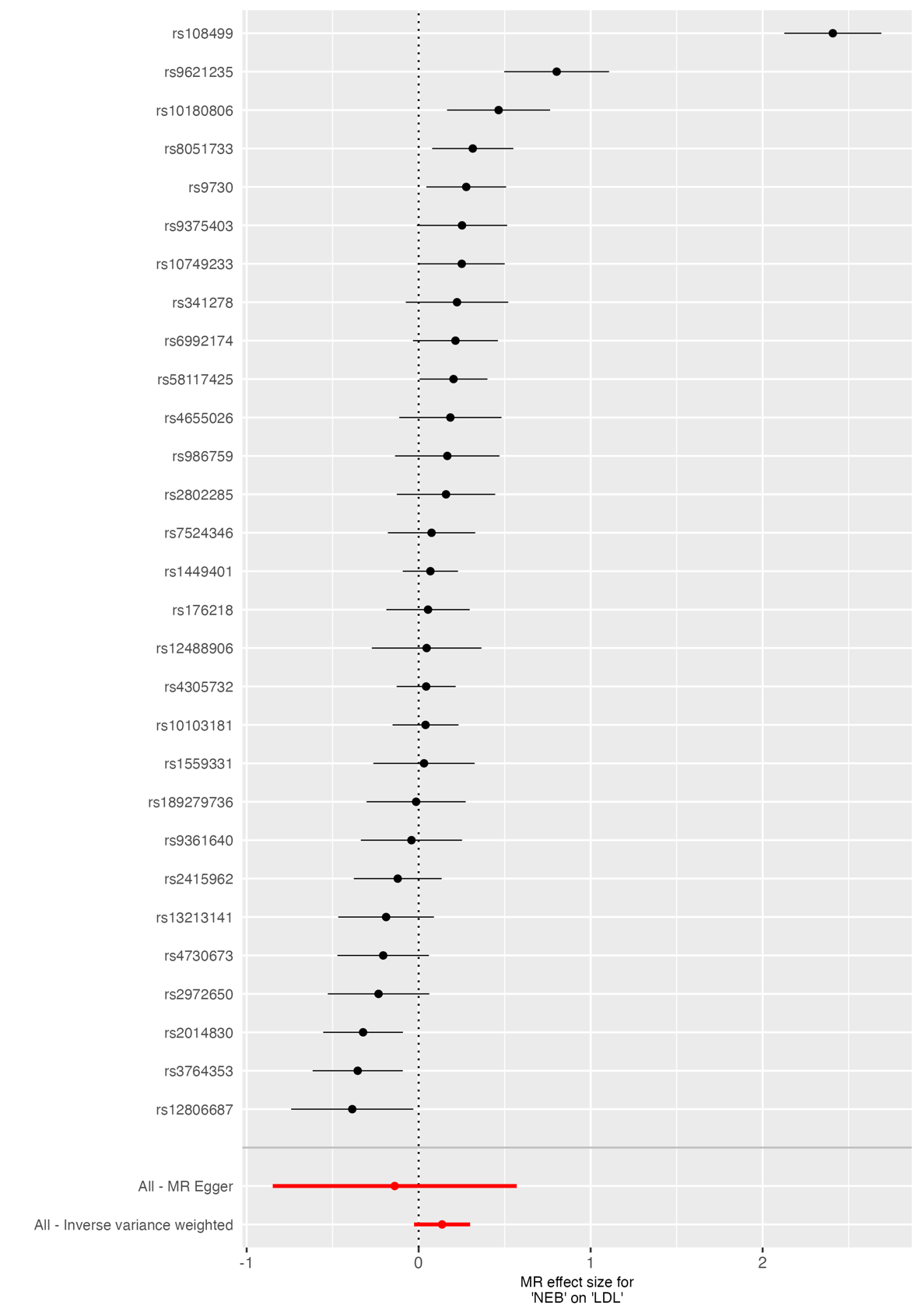

**Supplementary Figure 45.** *Forest plot of SNP-specific causal estimates for the effect of NEB on low-density lipoprotein (LDL) cholesterol.* Forest plot displaying individual Wald ratio estimates and 95% confidence intervals for each SNP used as an instrumental variable for parity in the two-sample MR analysis. Summary causal estimates derived from both inverse-variance weighted (IVW) and MR Egger methods are shown at the bottom of the plot. Variation in SNP-specific effects reflects heterogeneity across instruments. NEB; Number of children ever born.

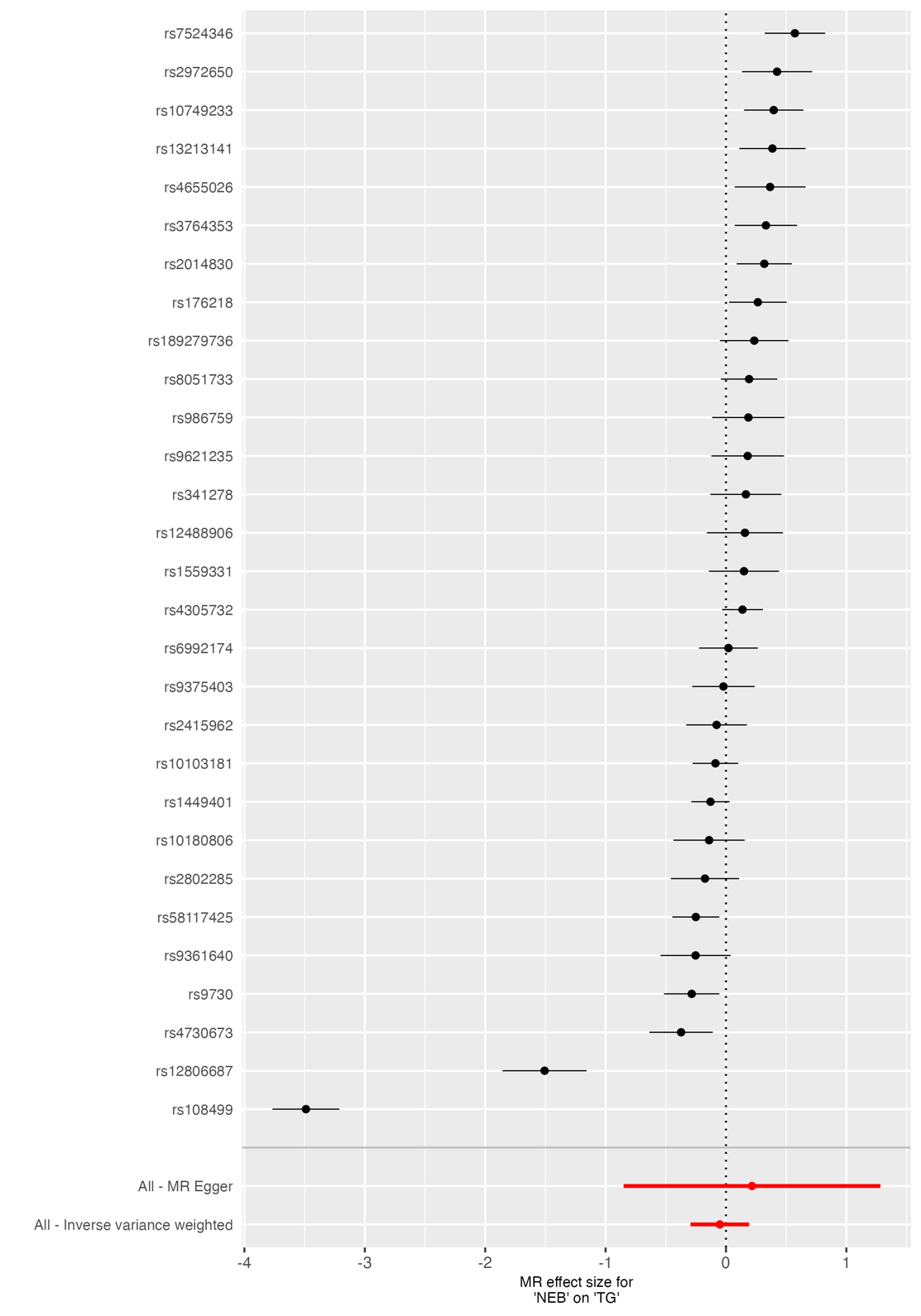

**Supplementary Figure 46.** *Forest plot of SNP-specific causal estimates for the effect of NEB on triglyceride (TG) level.* Forest plot displaying individual Wald ratio estimates and 95% confidence intervals for each SNP used as an instrumental variable for parity in the two-sample MR analysis. Summary causal estimates derived from both inverse-variance weighted (IVW) and MR Egger methods are shown at the bottom of the plot. Variation in SNP-specific effects reflects heterogeneity across instruments. TG in outcome GWAS was log-transformed. NEB; Number of children ever born.

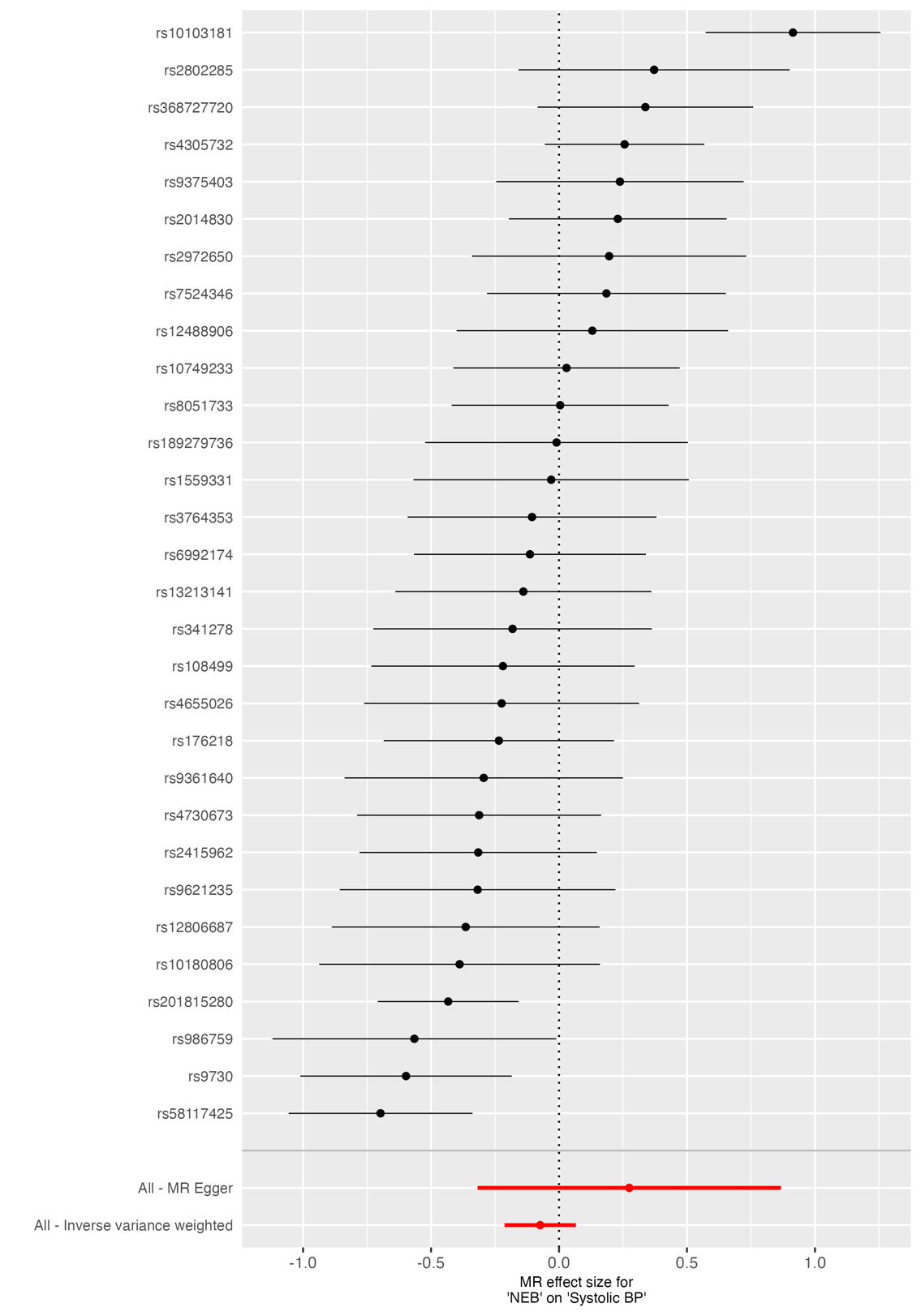

**Supplementary Figure 47.** *Forest plot of SNP-specific causal estimates for the effect of NEB on systolic blood pressure (BP).* Forest plot displaying individual Wald ratio estimates and 95% confidence intervals for each SNP used as an instrumental variable for parity in the two-sample MR analysis. Summary causal estimates derived from both inverse-variance weighted (IVW) and MR Egger methods are shown at the bottom of the plot. Variation in SNP-specific effects reflects heterogeneity across instruments. TG in outcome GWAS was log-transformed. NEB; Number of children ever born.

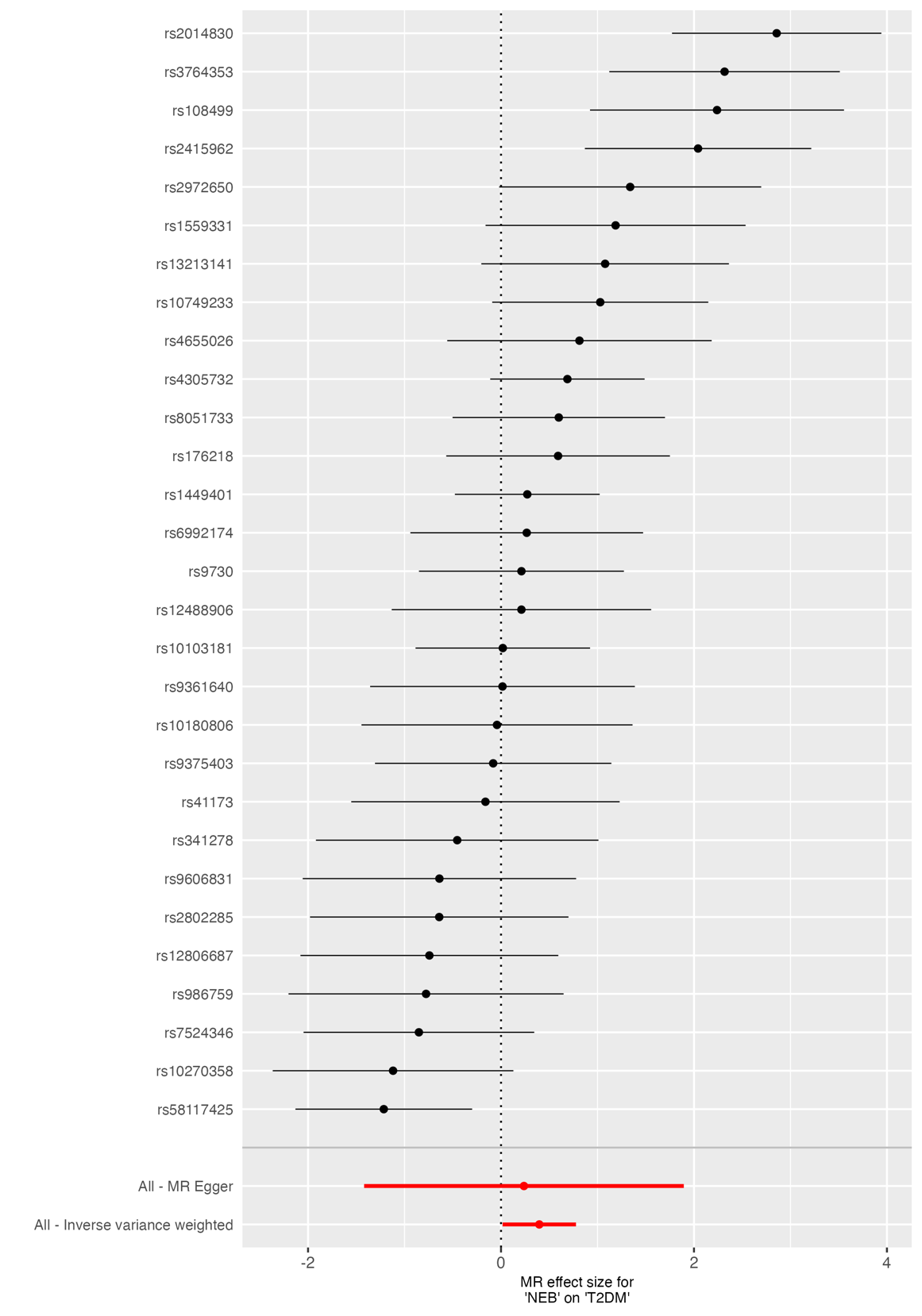

**Supplementary Figure 48.** *Forest plot of SNP-specific causal estimates for the effect of NEB on type-II diabetes (T2DM).* Forest plot displaying individual Wald ratio estimates and 95% confidence intervals for each SNP used as an instrumental variable for parity in the two-sample MR analysis. Summary causal estimates derived from both inverse-variance weighted (IVW) and MR Egger methods are shown at the bottom of the plot. Variation in SNP-specific effects reflects heterogeneity across instruments. TG in outcome GWAS was log-transformed. NEB; Number of children ever born.

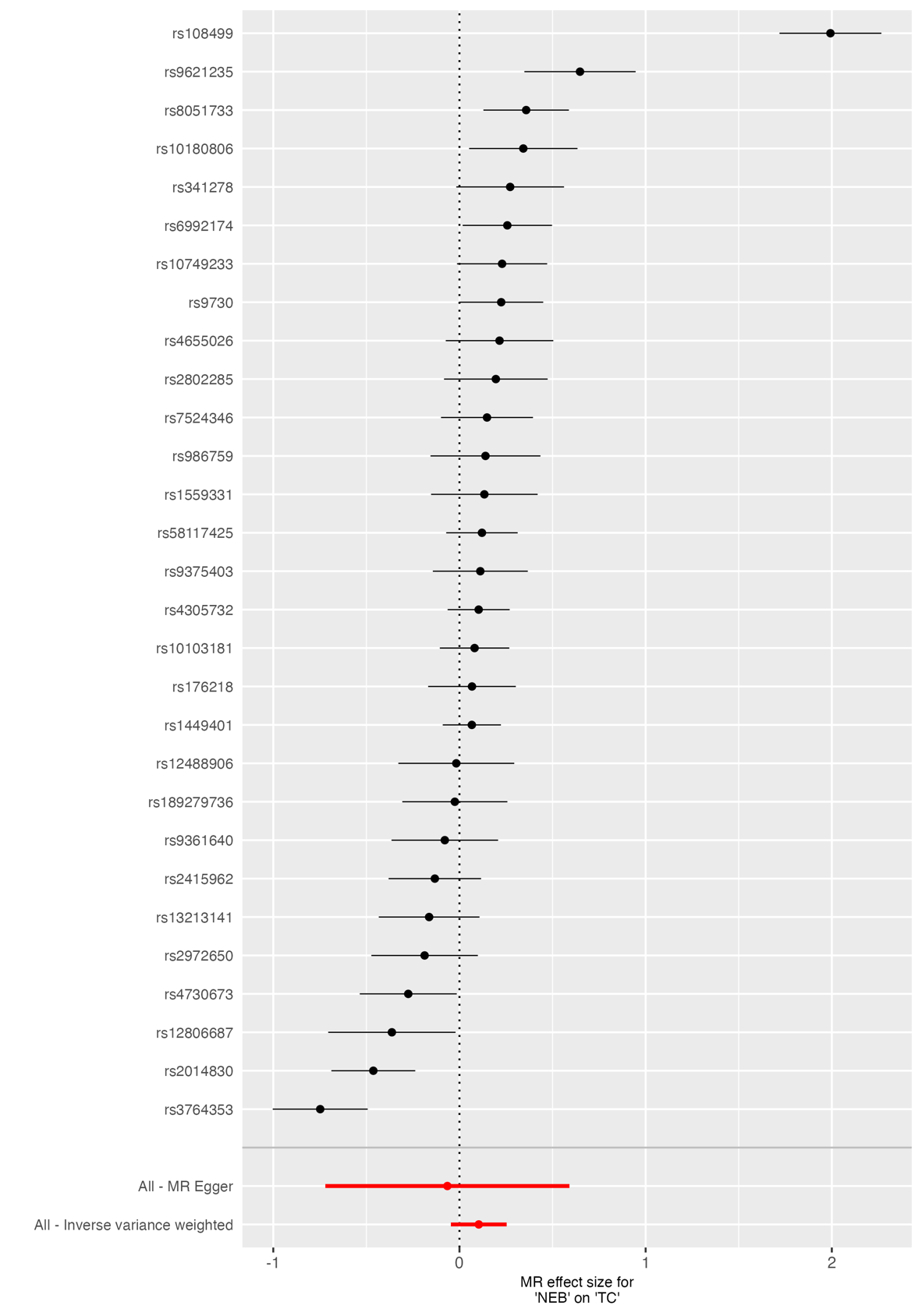

**Supplementary Figure 49.** *Forest plot of SNP-specific causal estimates for the effect of NEB on total cholesterol (TC).* Forest plot displaying individual Wald ratio estimates and 95% confidence intervals for each SNP used as an instrumental variable for parity in the two-sample MR analysis. Summary causal estimates derived from both inverse-variance weighted (IVW) and MR Egger methods are shown at the bottom of the plot. Variation in SNP-specific effects reflects heterogeneity across instruments. TG in outcome GWAS was log-transformed. NEB; Number of children ever born.

### **8.0 Power calculations**

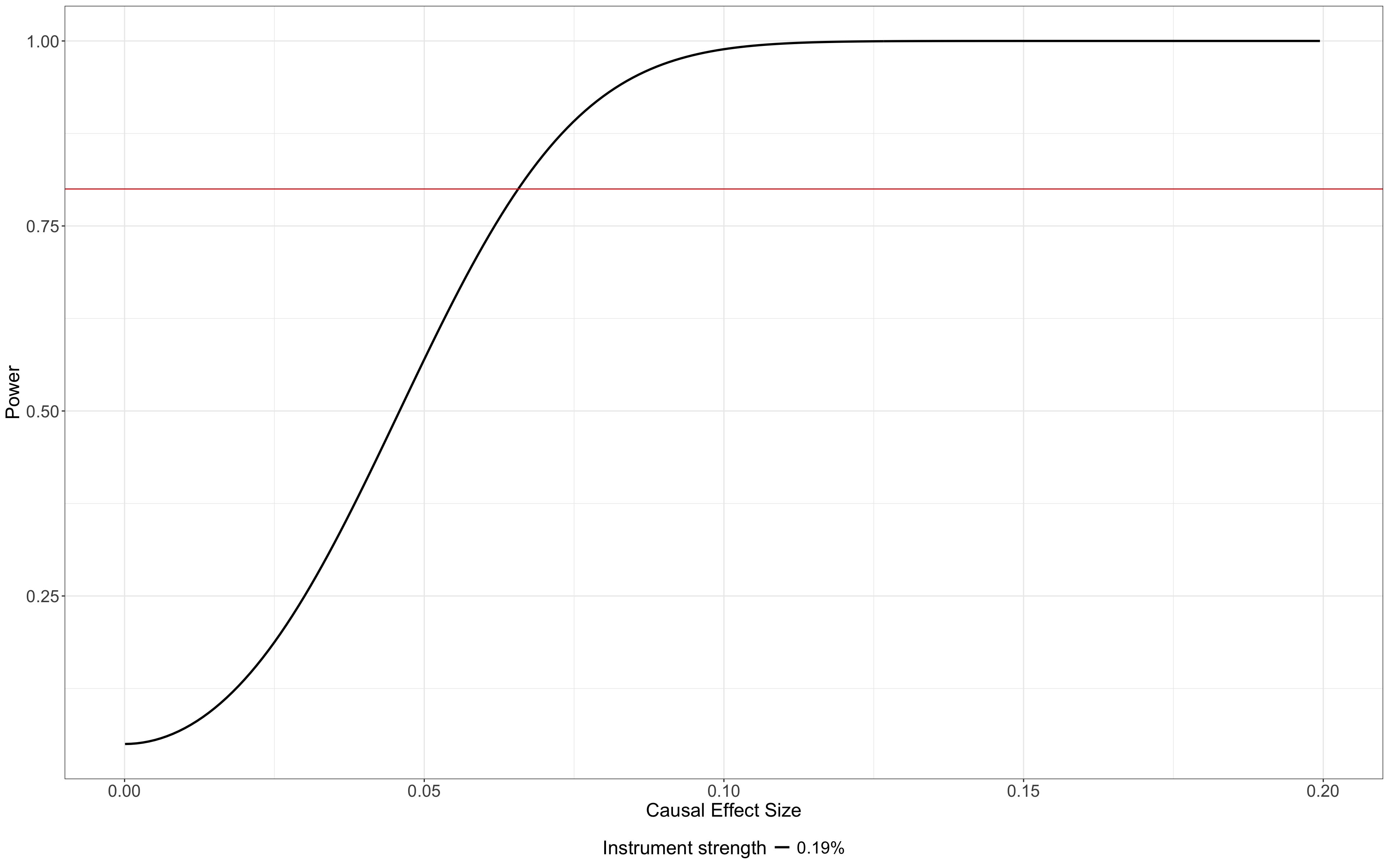

**Supplementary Figure 50.** *Power calculations for the traditional two-sample Mendelian randomisation analyses of a continous trait.* Power was estimated using the online MR power calculator (https://shiny.cnsgenomics.com/mRnd/), assuming a two-sided α = 0.05. The solid red line indicates 80% power, and the solid black line shows R^2^_(XZ)_ = 0.19% (variance explained reported in **Supplementary Table 9**). Calculations used a sample size of N= 400,000, variance of NEB (var(x))= 2.4, variance of the outcome (var(y))= 1, and an observational effect estimate (bOLS) = 0.0195.

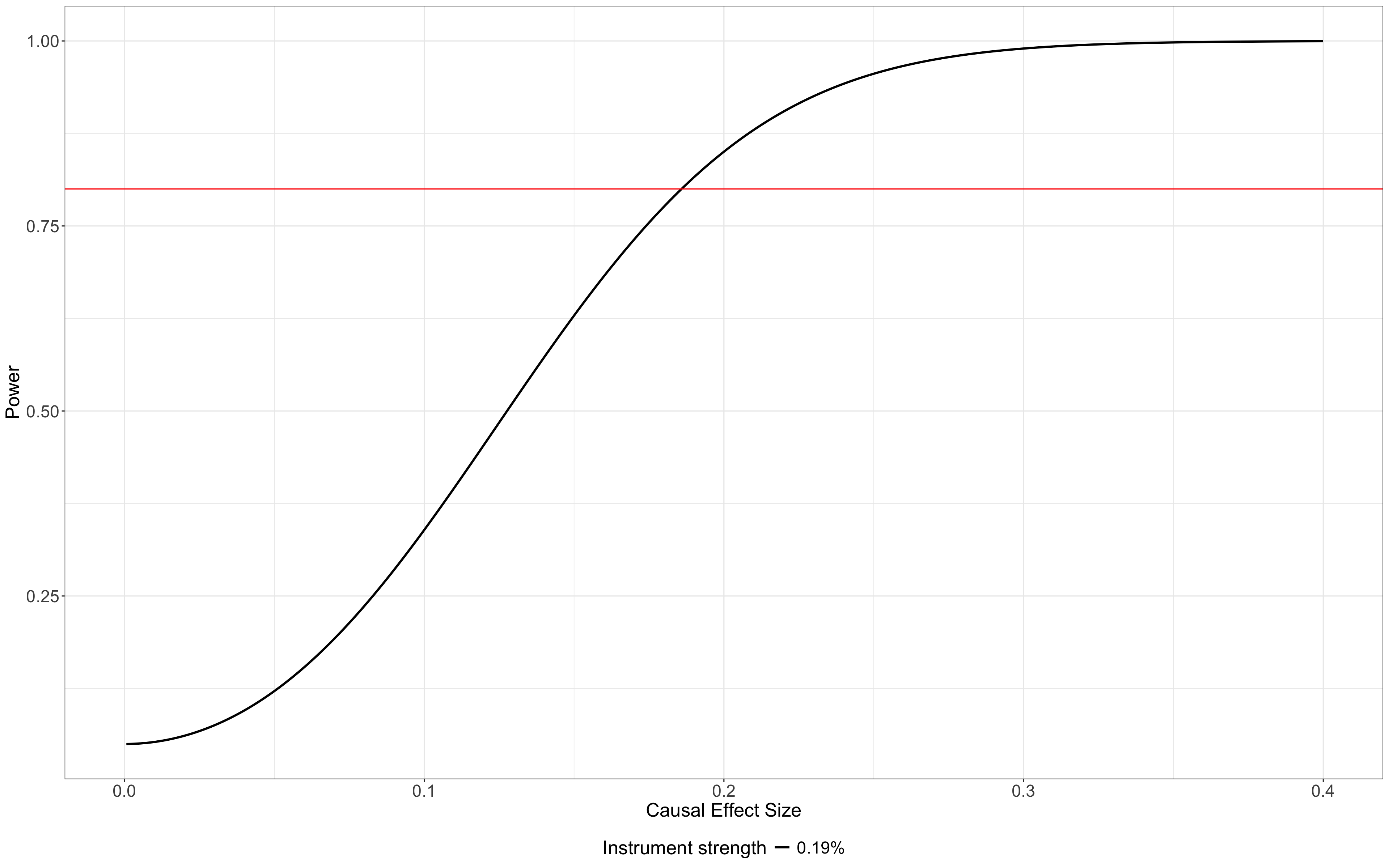

**Supplementary Figure 51.** *Power calculations for the spousal two-sample Mendelian randomisation analyses of a continous trait.* Power was estimated using the online MR power calculator (https://shiny.cnsgenomics.com/mRnd/), assuming a two-sided α = 0.05. The solid red line indicates 80% power, and the solid black line shows R^2^_(XZ)_ = 0.19% (variance explained reported in **Supplementary Table 9**). Calculations used a sample size of N= 53,000, variance of NEB (var(x))= 2.4, variance of the outcome (var(y))= 1, and an observational effect estimate (bOLS) = 0.0195.

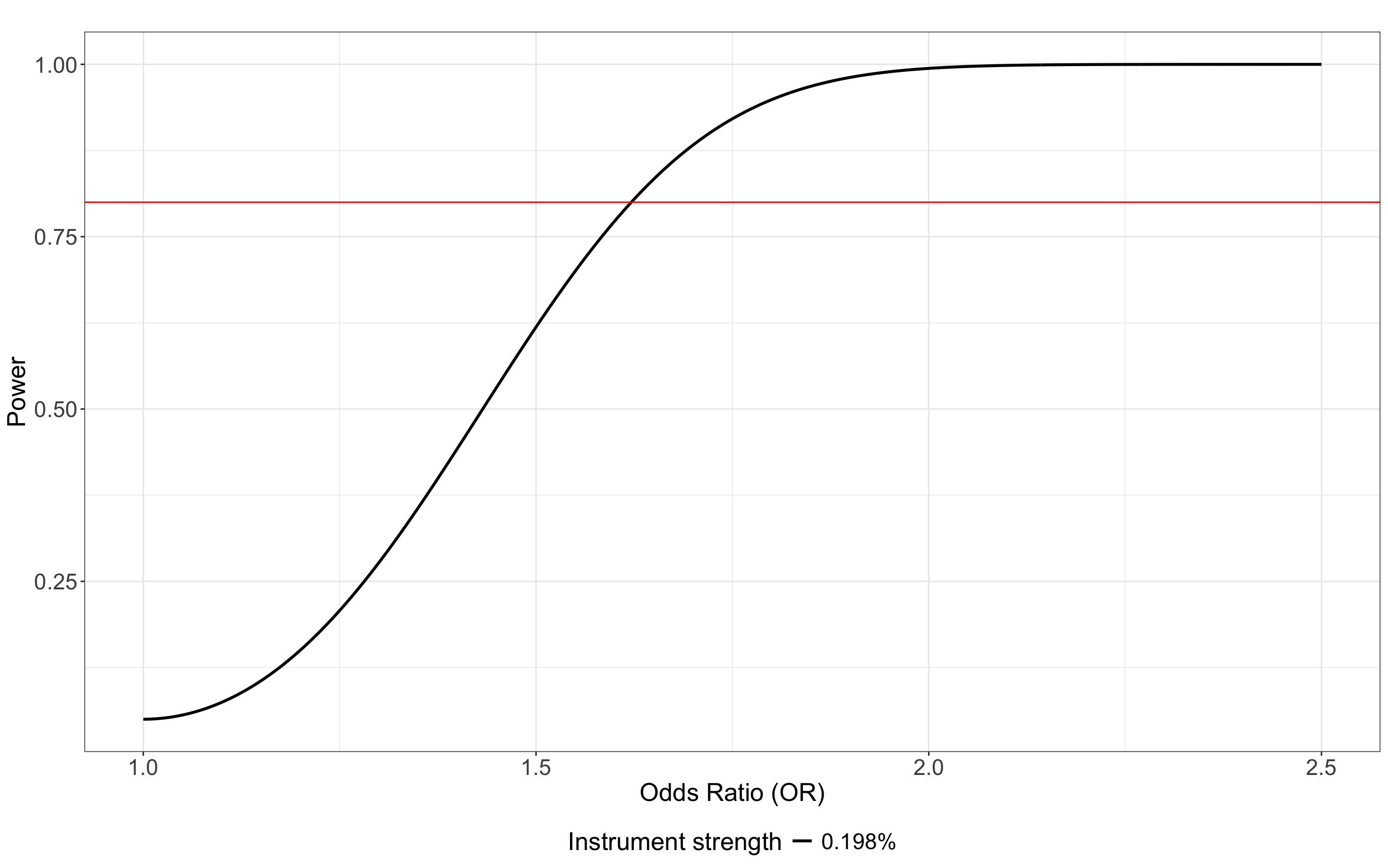

**Supplementary Figure 52.** *Power calculations for the traditional two-sample Mendelian randomisation analyses of a binary trait.* Power was estimated using the online MR power calculator (https://shiny.cnsgenomics.com/mRnd/), assuming a two-sided α = 0.05. The solid red line indicates 80% power, and the solid black line shows R^2^_(XZ)_ = 0.19% (variance explained reported in **Supplementary Table 9**). Calculations used a sample size of N = 200,000 and a trait prevalence of K = 6%.

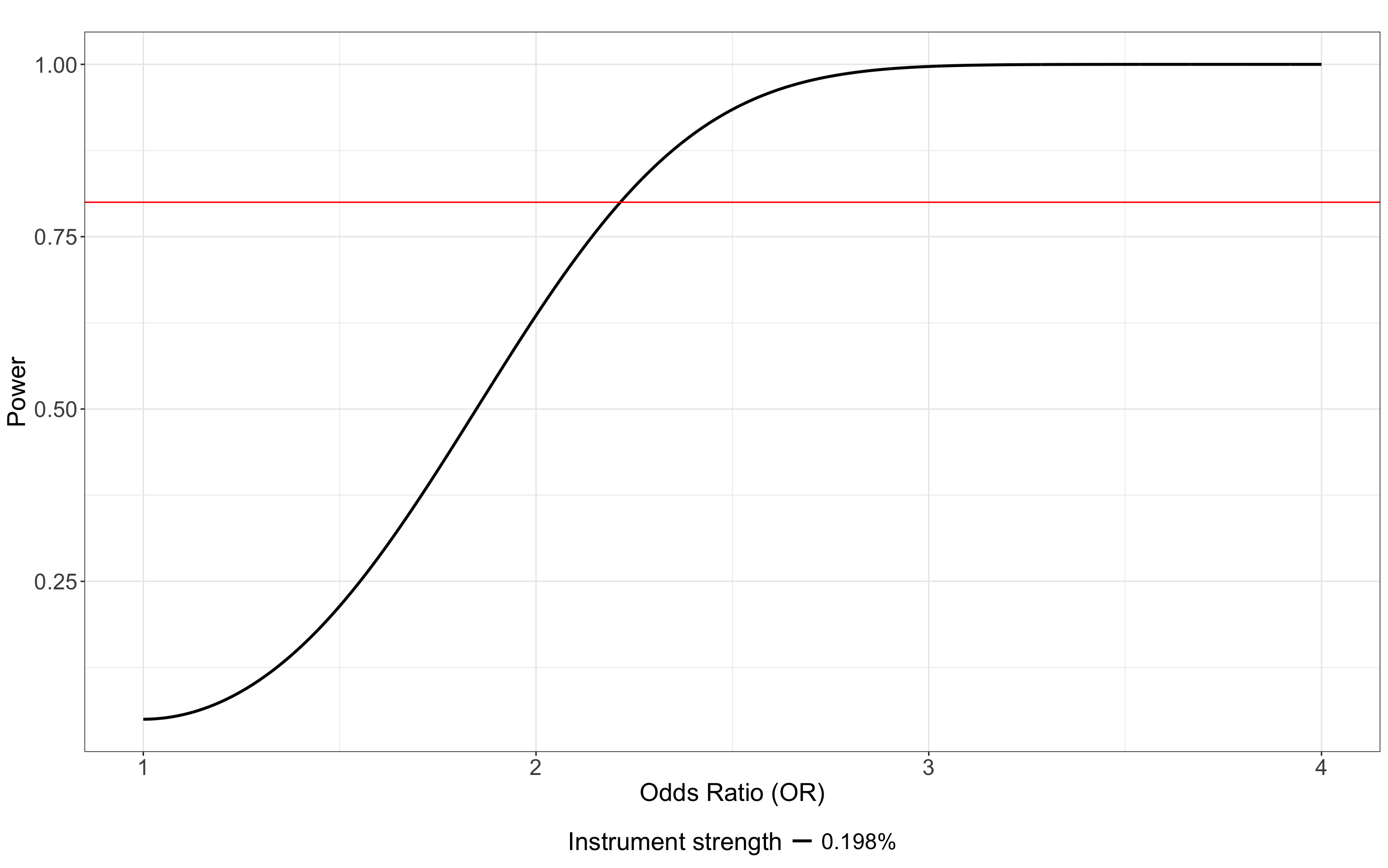

**Supplementary Figure 53.** *Power calculations for the spousal Mendelian randomisation analyses of a binary trait.* Power was estimated using the online MR power calculator (https://shiny.cnsgenomics.com/mRnd/), assuming a two-sided α = 0.05. The solid red line indicates 80% power, and the solid black line shows R^2^_(XZ)_ = 0.19% (variance explained reported in **Supplementary Table 9**). Calculations used a sample size of N = 53,000 and a trait prevalence of K = 6%.
